# Parent-mediated interventions versus usual care in children with autism: A systematic review with meta-analysis and Trial Sequential Analysis

**DOI:** 10.64898/2026.08.06.26357818

**Authors:** Charlotte Engberg Conrad, Sonja Martha Teresa Ziegler, Niels Bilenberg, Jens Christiansen, Kirstine Agnete Davidsen, Birgitte Fagerlund, Emil Færk, Helle Jakobsen, Rikke Hermann Jakobsen, Pia Jeppesen, Caroline Kamp, Tina Røndrup Kilburn, Per Hove Thomsen, Manon Varenne, Martin Vestergaard, Janus Christian Jakobsen, Marlene Briciet Lauritsen

## Abstract

**Objectives:** To evaluate the positive and adverse effects of parent-mediated interventions (PMIs) versus care as usual for children with autism.

**Setting:** Systematic review and meta-analysis and Trial Sequential Analyses (TSA), following the Preferred Reporting Items for Systematic Reviews and Meta-Analyses guidelines.

**Methods:** We searched for randomised clinical trials of PMIs for children with autism in the databases CENTRAL, EMBASE, LILACS, PsycINFO, MEDLINE, and SCI-EXPANDED (up to 13 August, 2025), complemented with manual searches. 12,359 articles were screened. Data were synthesised using meta-analyses and Trial Sequential Analyses (TSA), and risks of bias and certainty of the evidence were evaluated.

**Primary and secondary outcome measures:** The primary outcome was autism characteristics. Secondary outcomes were adverse effects, child adaptive functioning, child language, child and parent quality of life, and parental stress. Ten exploratory outcomes were included.

**Results:** 32 trials (N=1,625) comparing PMIs to usual care, waiting list, or no intervention were included. All trials had a high risk of bias. The multiplicity-adjusted threshold for statistical significance was *p* = 0.013 due to the number of outcomes. Meta-analyses and TSAs showed it could be rejected that PMIs reduced autism characteristics (MD = −0.88; 95% confidence interval −2.92 to 1.15; *p* = 0.05, 4 trials, *N*=353, low certainty), child adaptive functioning (7 trials, *N*=408), child language (4 trials, *N*=308), or parental stress (7 trials, *N*=385). Due to insufficient data, the remaining secondary meta-analyses could not be conducted. Meta-analyses of exploratory outcomes showed beneficial effects concerning child behaviour problems and parent sensitivity/synchronicity.

**Conclusions:** This meta-analysis found no benefits of PMIs on child autism characteristics, child adaptive functioning, child language, or parental stress. Benefits were found in reduction of child behaviour problems and improved parent sensitivity/synchronicity. The evidence remains uncertain, and more trials including outcomes of adverse effects and quality of life are needed.

**Trial registration:** CRD42022385188 [PROSPERO](https://www.crd.york.ac.uk/PROSPERO/view/CRD42022385188)

---

Autism spectrum disorder (ASD) is one of the most common neurodevelopmental conditions in high-income countries, with an overall prevalence of 2.8% in the US (1) and a global prevalence of approximately 1% (2) and is characterised by difficulties in social communication, repetitive behaviours, and sensory sensitivity (3, 4). Due to these core manifestations, early support for children is key and often includes interventions that specifically aim to improve communication and social interaction skills (4, 5).

Interventions for autistic children are founded on different theoretical and methodological backgrounds, e.g., behavioural and developmental frameworks or a combination of the two, often referred to as Naturalistic Developmental Behavioural Intervention (NDBI) (6–8). In parent-mediated interventions (PMIs) parents or primary carers (hereafter referred to as parents) are considered the primary (4) agents of change in the child’s development (9). Parents receive education, training, and coaching from skilled professionals, enabling them to maximise the child’s learning and development opportunities across contexts. Parents are able to provide a greater intensity of the intervention strategies, due to the time spent with their child (10). PMIs have increased in use as they are considered cost-effective and offer the opportunity to apply the intervention strategies in daily life in a sensitive manner (10, 11).

Previously conducted systematic reviews of PMIs for childhood autism vary significantly in methodology and quality (8). Based on AMSTAR 2 criteria (12), Conrad et al. (2025) concluded that only one (13) out of nine reviews conducted between 2013 and 2021 was of high quality, while one (14) was of low quality, and seven were of critically low quality (15–21). This highlights the need for an updated, high-quality, evidence-based synthesis of the effects of PMIs. Furthermore, no previous meta-analyses have investigated the adverse effects of PMIs. We therefore conducted a systematic review and meta-analysis to evaluate the positive and adverse effects of PMIs versus care as usual, waiting list, or no intervention on the outcomes for autistic children and their parents.

## Methods

The objective of this systematic review and meta-analysis was to synthesise the current evidence of both positive and adverse effects of PMIs versus usual care for children with autism and their parents through improved methodology and inclusion of newly published studies. Our methodology is described in detail in our protocol submitted prior to conducting the meta-analysis (8).

We carried out this systematic review following the recommendations of the Preferred Reporting Items for Systematic Reviews and Meta-Analyses (PRISMA) guidelines (22) (Appendix S1 for PRISMA Checklist).

We searched for randomised clinical trials of PMIs for children with autism in the databases CENTRAL, EMBASE, LILACS, PsycINFO, MEDLINE, and SCI-EXPANDED. We included all randomised clinical trials comparing PMIs versus usual care, waiting list, or no intervention for either children with a confirmed diagnosis of ASD or for younger children with characteristics strongly suggesting autism (excluding children with only a familial likelihood of autism) where parents implemented at least 80% of the interventions. Two authors (CEC, SMTZ) independently searched for trials identified prior to August 2025 (Appendix S2 Search History). We included randomised clinical trials regardless of trial design, setting, publication status, year, language (i.e., English and Spanish), and reporting of outcomes. The first author worked in pairs with another author (SMTZ, MBL, CBK or MV), and for three of the studies the authors MBL and SMTZ worked in a pair to extract and independently assess the risks of bias in included trials. A third author was consulted to resolve discrepancies within the pair (CBK or CEC). We contacted trial authors by email if data were unclear or missing.

### Outcomes

#### Primary outcome

- *Autism Characteristics* as measured by the clinician-rated Autism Diagnostic Observation Schedule (ADOS) (23) using the ADOS total score with continuous outcome scores. The ADOS measures the child’s social communicative behaviour and repetitive and restricted behaviour during a standardized intervention.

#### Secondary outcomes

For all secondary outcomes one specific outcome measure was chosen, believed to be widely used in research.

- *Adverse effects* as measured by the Negative Effects Questionnaire (NEQ) (24)
- *Child adaptive functioning* as measured by any version of the parent-rated Vineland Adaptive Behavior Scale (VABS) (25)
- *Child’s quality of life* as measured by the parent-rated Pediatric Quality of Life Inventory (PEDSQL) (26)
- *Child language* as measured by the Mullen Scales of Early Learning Receptive Language subscale (MSEL) (27)
- *Parental quality of life* as measured by the World Health Organization Quality of Life Assessment-BREF (WHOQOL-BREF) (28)
- *Parental Stress* as measured by the Parenting Stress Index (PSI) (29)

#### Exploratory outcomes

- *Child behaviour problems*
- *Joint attention*
- *Child adaptive functioning (not measured by VABS)*
- *Parent synchrony/sensitivity*
- *Attachment*
- *Parent fidelity to intervention implementation*
- *Family life functioning*
- *Autism characteristics (not measured by ADOS total score)*
- *Child’s social communication*
- *Child’s repetitive and restrictive behaviour*

For all outcomes, we used the trial results reported at the time point closest to the end of intervention as defined by trialists.

### Subgroup analyses

We predefined subgroup analyses for our outcomes by assessing risk of bias (high risk of bias versus low risk of bias), types of PMIs (behavioural, developmental, NDBI, or other), type of control groups (usual care, waiting list, or no intervention), parental household configuration (single versus two parents), age of the child (younger versus older) and autism severity.

We used the formal test for subgroup interactions in Stata (30).

### Assessment of risk of bias

We assessed risk of bias according to the Cochrane Handbook for Systematic Reviews of Interventions 5.1 (31). We evaluated domains of random sequence generation, allocation concealment, blinding of participants and intervention providers, blinding of outcome assessment, incomplete outcome data, selective outcome reporting, and other risks of bias. We used these domains to evaluate the included trials as being overall low or high risk of bias as described in our protocol (8).

The domains blinding of outcome assessment, incomplete outcome data, and selective outcome reporting were further assessed separately for each outcome result.

### Data Synthesis and Meta-analyses

#### Assessment of effect

We calculated mean differences (MDs) with a 95% confidence interval (CI) for the continuous primary and secondary outcomes. When there was more than one continuous outcome measure in an exploratory outcome, the standardised mean difference (SMD) with a 95% CI for continuous outcomes was calculated.

#### Assessment of heterogeneity

Firstly, we investigated forest plots to assess any sign of heterogeneity visually. Secondly, we assessed the presence of statistical heterogeneity by the chi^2^ test (threshold P-value < 0.10) and measured the quantities of heterogeneity by the *I^2^* statistic (32, 33). Possible heterogeneity through subgroup analyses was investigated, and we ultimately decided whether a meta-analysis should be avoided (34).

##### Assessment of reporting biases

We used a funnel plot to assess possible reporting bias, if ten or more trials were included. For continuous outcomes, the regression asymmetry test (35) and adjusted rank correlation (36) were used.

##### Assessment of statistical and clinical significance

We performed all meta-analyses using the statistical software Stata version 17 (30). We assessed the intervention effects with both random-effects (37) and fixed-effects meta-analyses (38). We reported the most conservative result (highest P-value) as the primary analysis result and the least conservative result as a sensitivity analysis (39). A total of seven primary and secondary outcomes were assessed; thus, a P-value of 0.013 or less was considered statistically significant (39). We used the eight-step procedure to assess if the thresholds for significance were crossed (39). When analysing the exploratory outcomes, we used a P-value of 0.05 as the threshold for statistical significance, as these outcomes were only considered hypothesis-generating (39).

We performed Trial Sequential Analysis (TSA) to control for the risk of false-positive results (type 1 errors) and false-negative results (type 2 errors) (40). Traditional meta-analyses run the risk of random errors due to sparse data and repetitive testing of accumulated data when updating reviews. The TSA was performed on the primary and secondary outcomes to calculate the required information size (i.e., number of participants needed in a meta-analysis to detect or reject a certain intervention effect) and the cumulative *Z*-curve’s breach of relevant trial sequential monitoring boundaries (41–48).

For continuous outcomes, the TSA used the observed SD, a mean difference of the observed SD/2, an alpha of 1.3% for all outcomes, a beta of 20%, and the observed diversity, as suggested by the trials in the meta-analysis.

To evaluate the quality of the evidence underlying the meta-analyses for the pre-specified outcomes, we applied the five GRADE (Grading of Recommendations Assessment, Development and Evaluation) considerations: risk of bias, consistency of the effect estimates, imprecision, indirectness, and publication bias (39, 40, 49, 50).

#### Patient and public involvement

A parent representative of a child with autism is a member of the steering committee of the DAN-PACT randomised clinical trial, within the context of which this systematic review was conducted. The steering committee was consulted on the research question and scope of the review, ensuring that the perspectives of families affected by autism informed the design of this work.

#### Differences between the protocol and the review

When more than one outcome type was available for explorative outcomes, clinician-rated outcomes were prioritised before parent-rated outcomes, as clinicians – unlike parents – were usually blinded to groups and because they use best-practice methods to increase the data validity.

The secondary outcome of child language as measured by MSEL has been specified as the subscale of receptive language.

A few terms e.g., labels of outcomes have been changed to avoid ableist language.

## Results

### Included studies

#### Participants

A total of 1,625 children (ages nine months to 12 years; 18.8% females) were included across the 34 publications, representing 32 trials (two trials were published in two papers investigating different aspects of the same trials) (51–54). All included studies investigated interventions mediated by parents during at least 80% of the intervention and targeted aspects of social communication and/or autistic behaviours. Due to their young age of 9-14 months, participants in one study (55) were included in the randomised clinical trial if they showed at least three of five early behavioural signs of autism on the Social Attention and Communication Surveillance-Revised (SACS-R). The largest study, (56) had 152 participants. The numbers of males and females was reported in 28 of 32 studies, with 1304 males and 305 females (male: female ratio of approximately 4.3:1) (51–80). Four trials did not report on the numbers of males and females (81–84). A detailed description of all studies is included in Table 1 and a narrative description (Appendix S3).

**Table 1.**
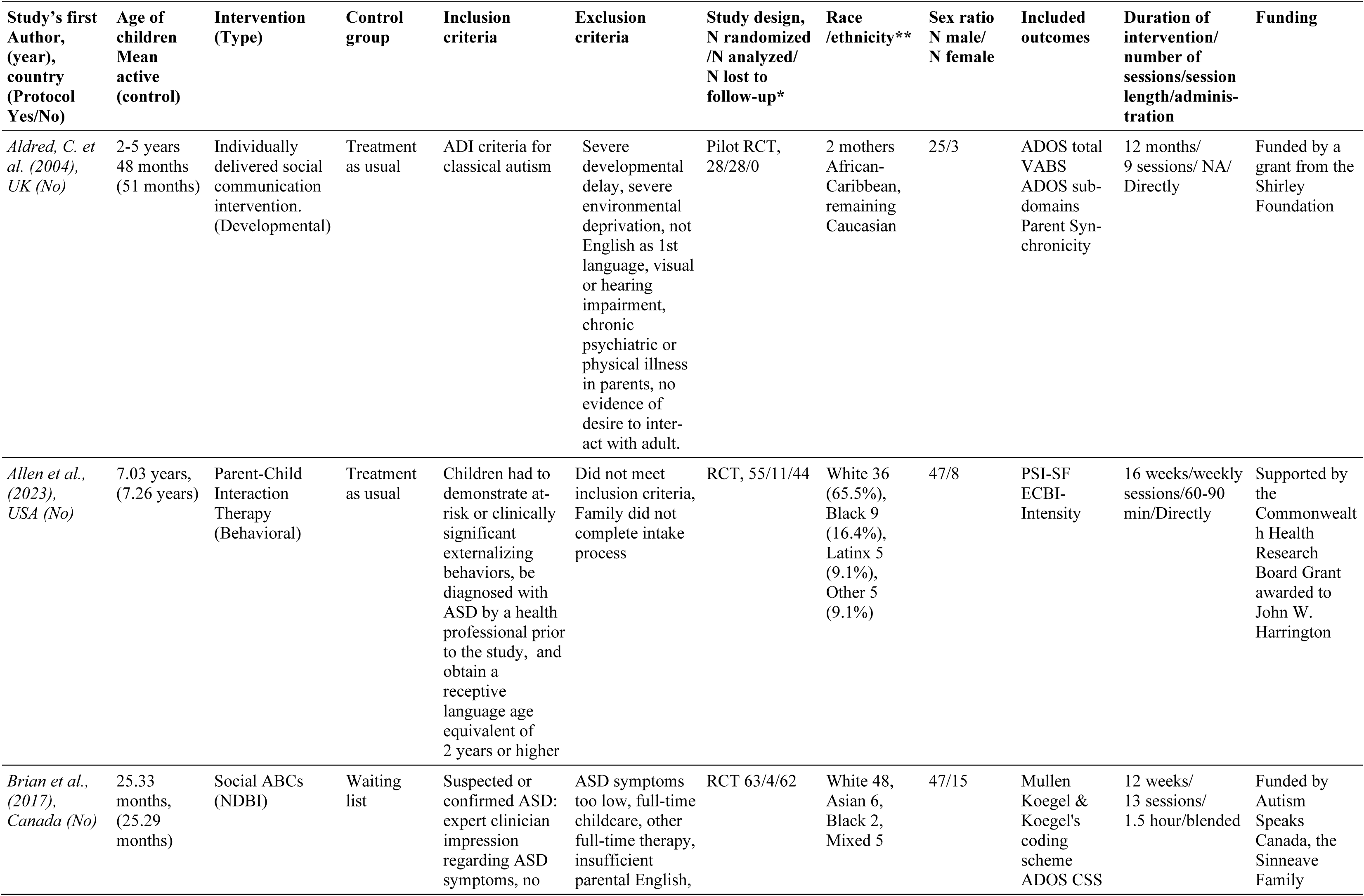
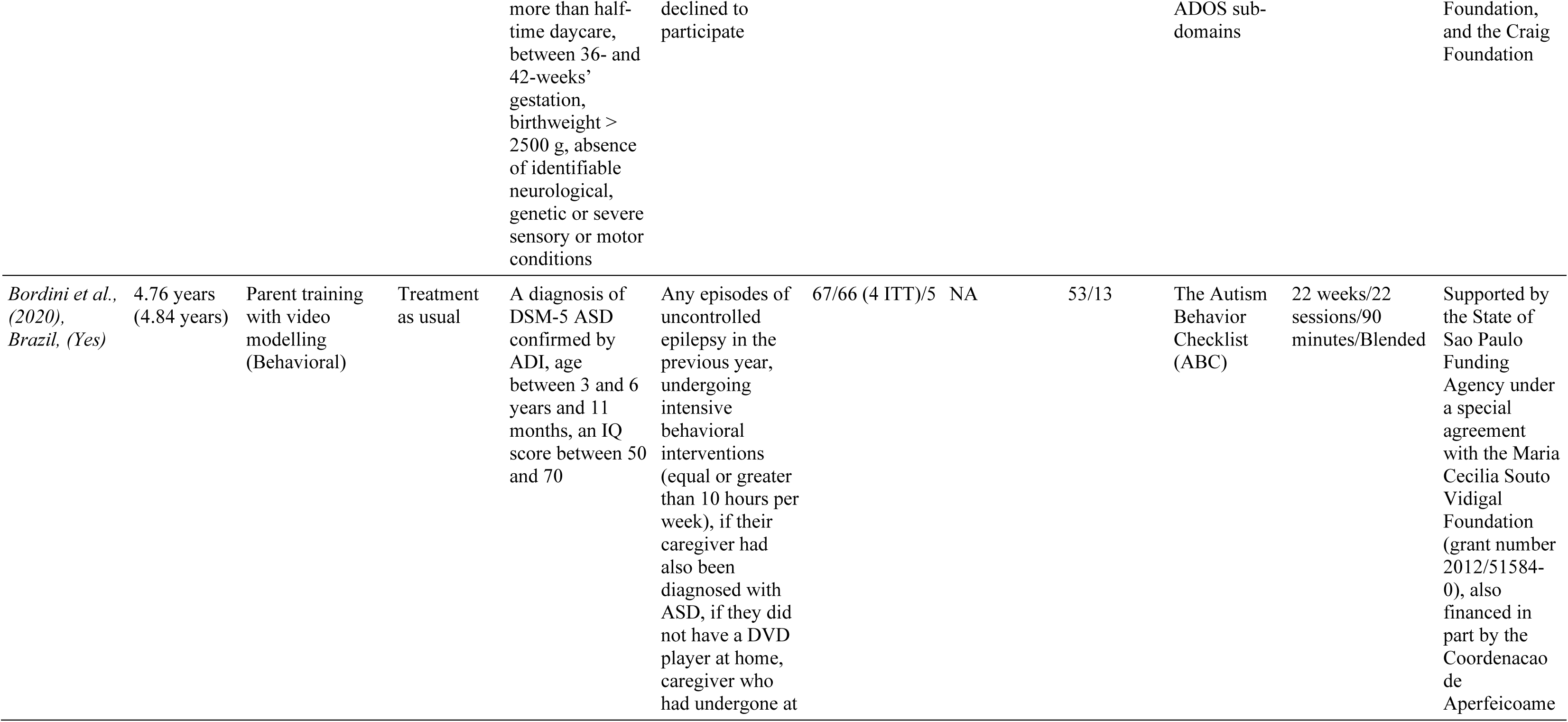
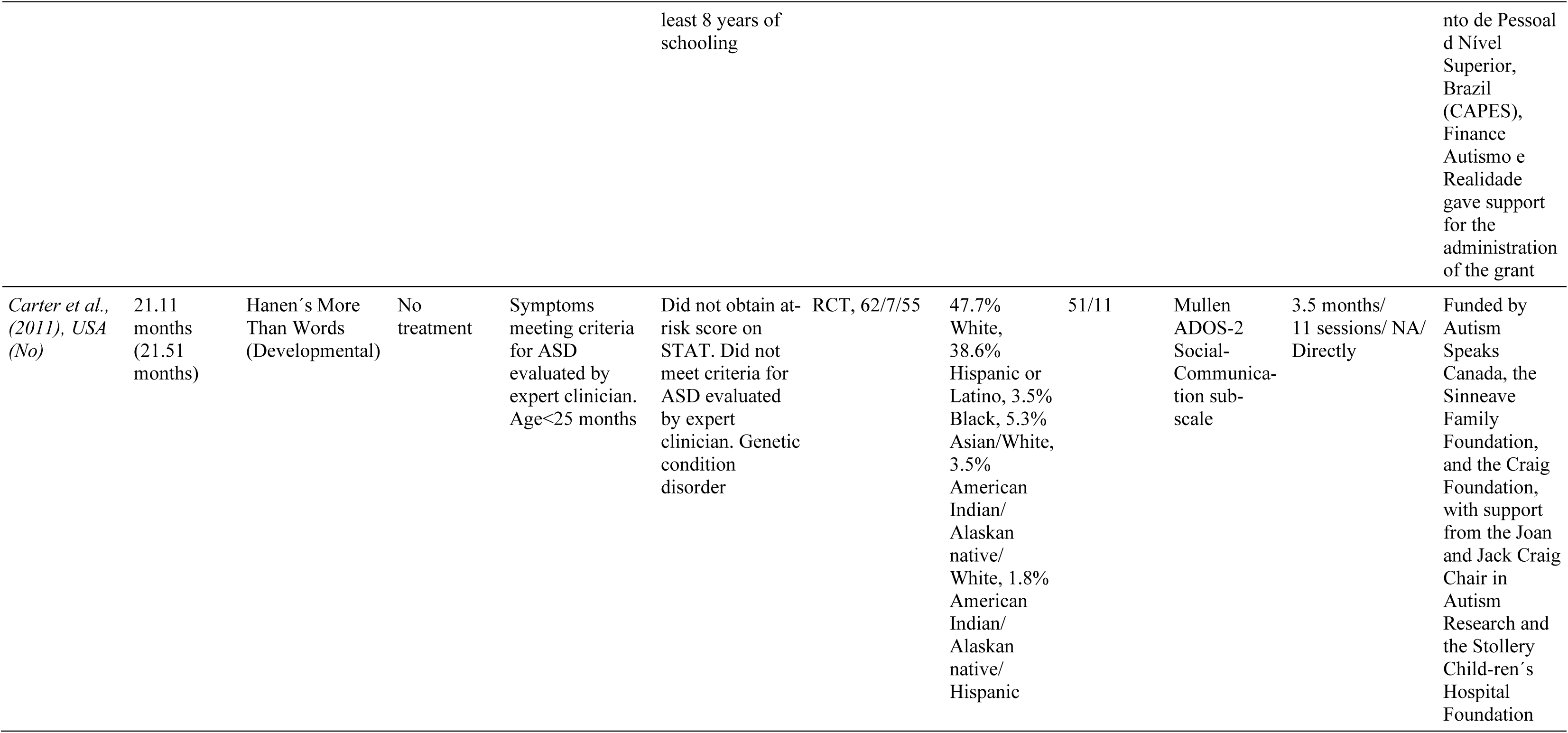
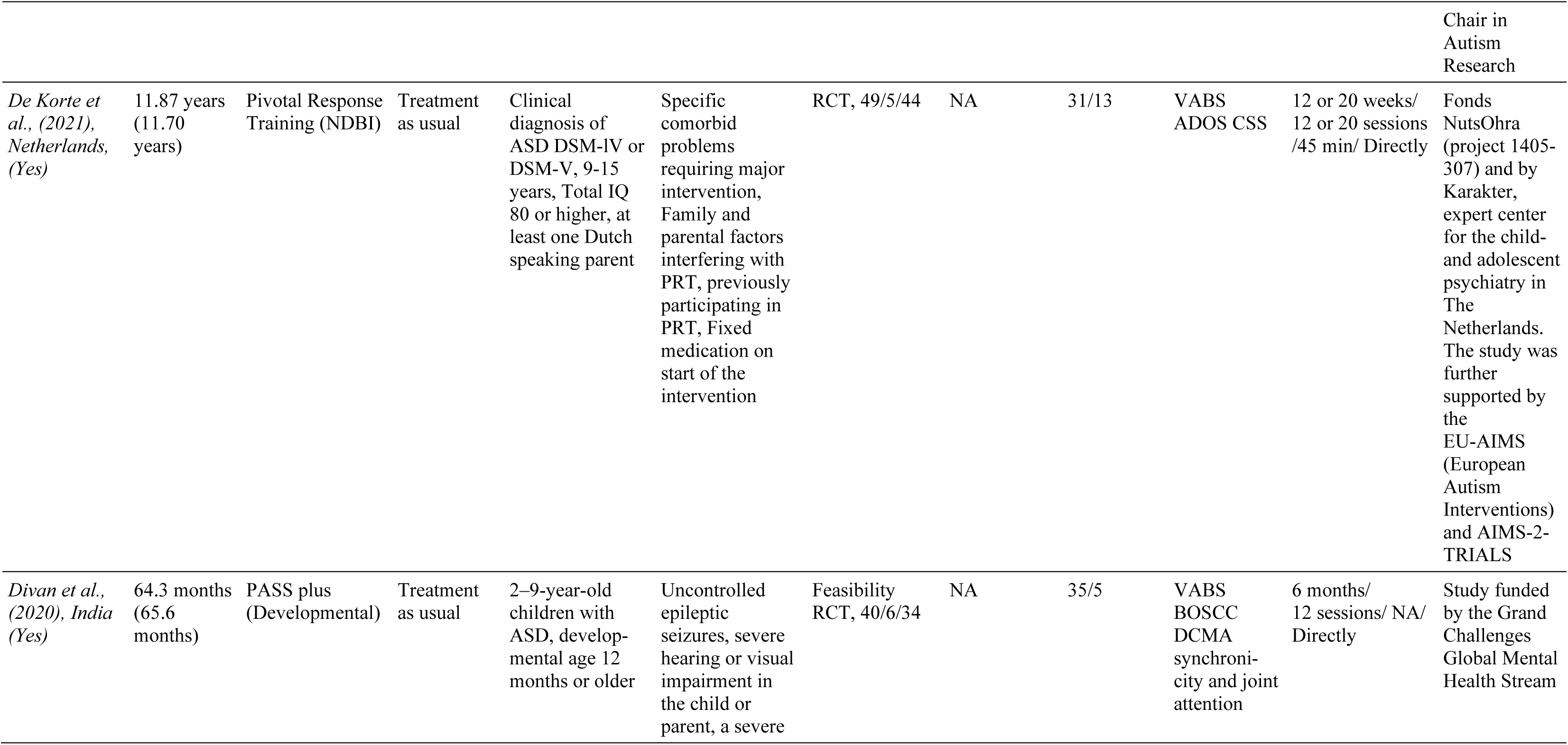
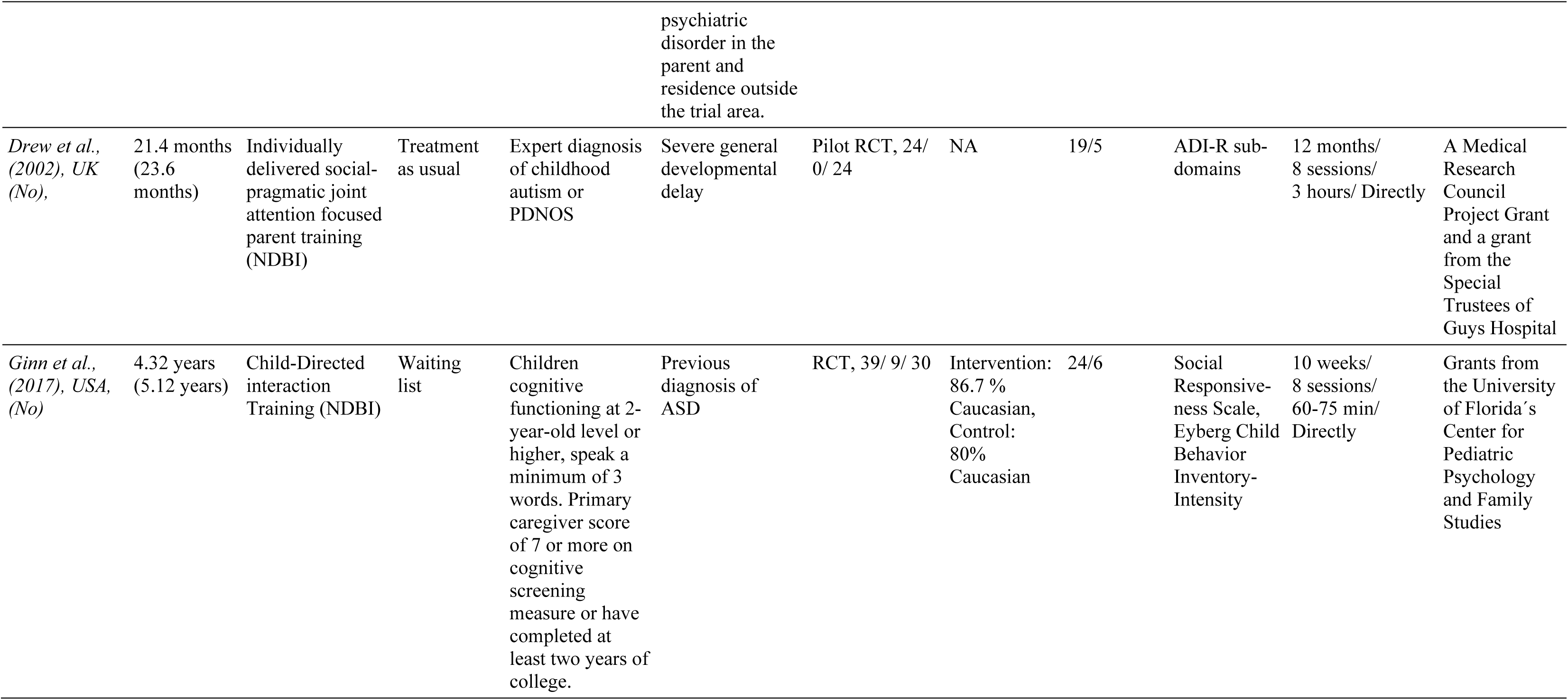
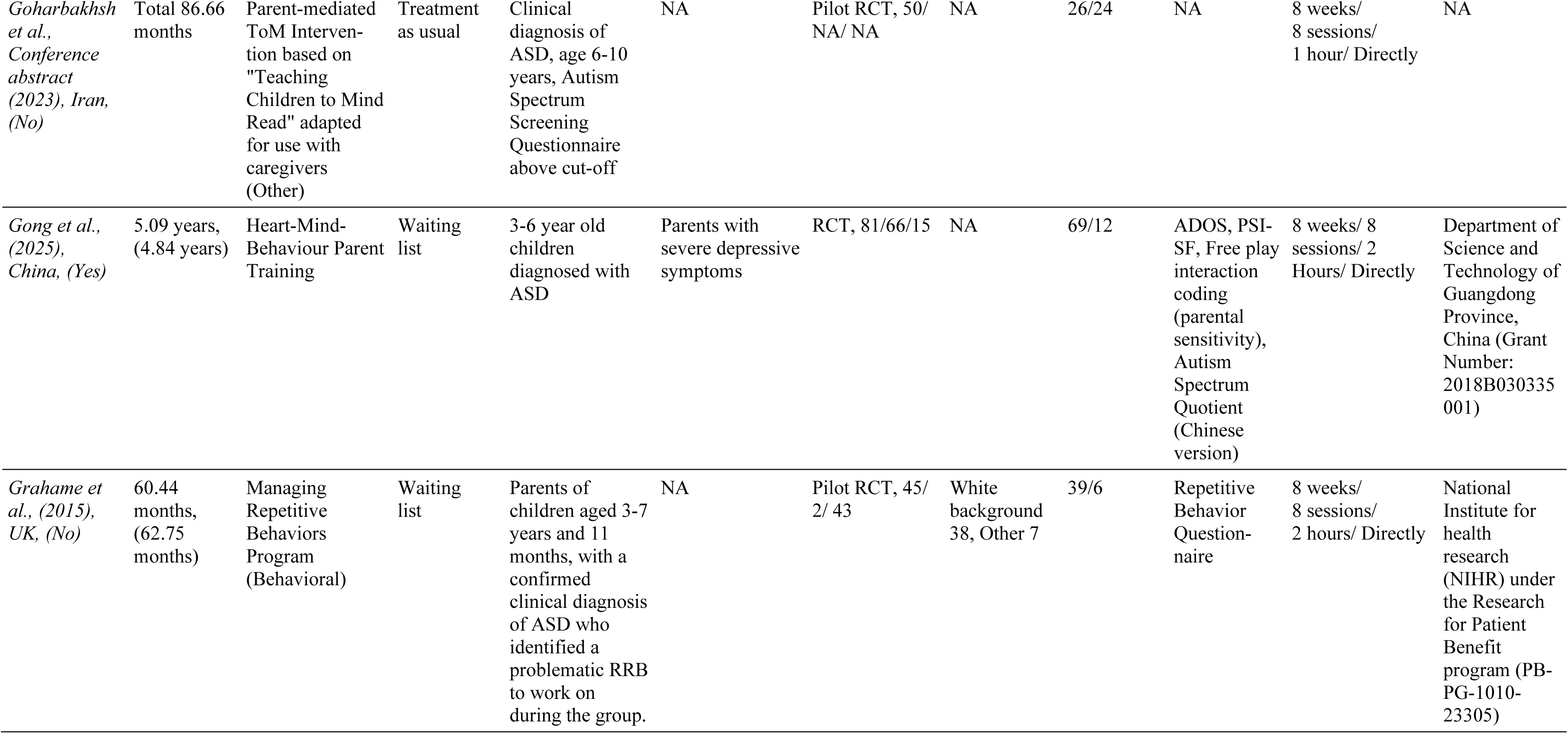
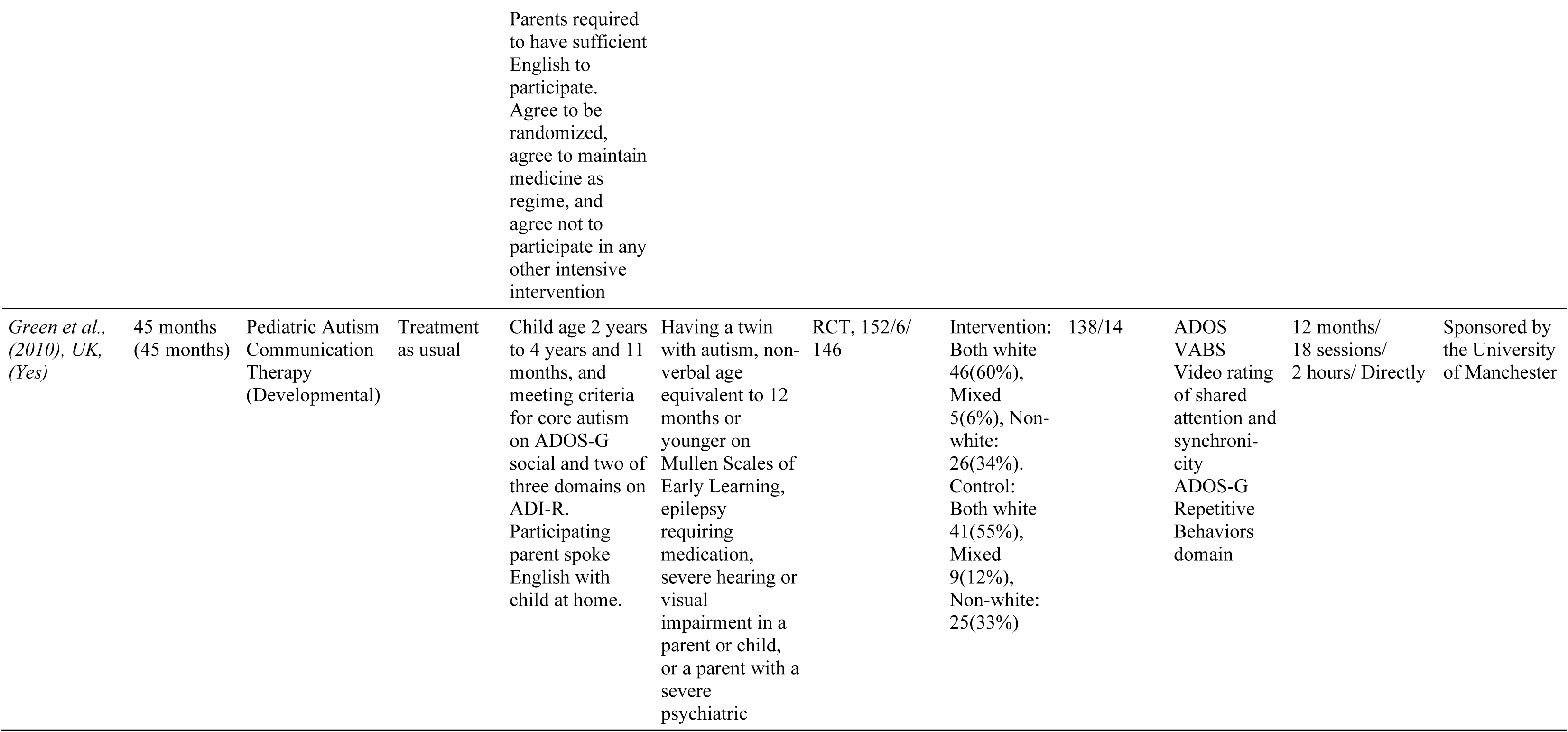
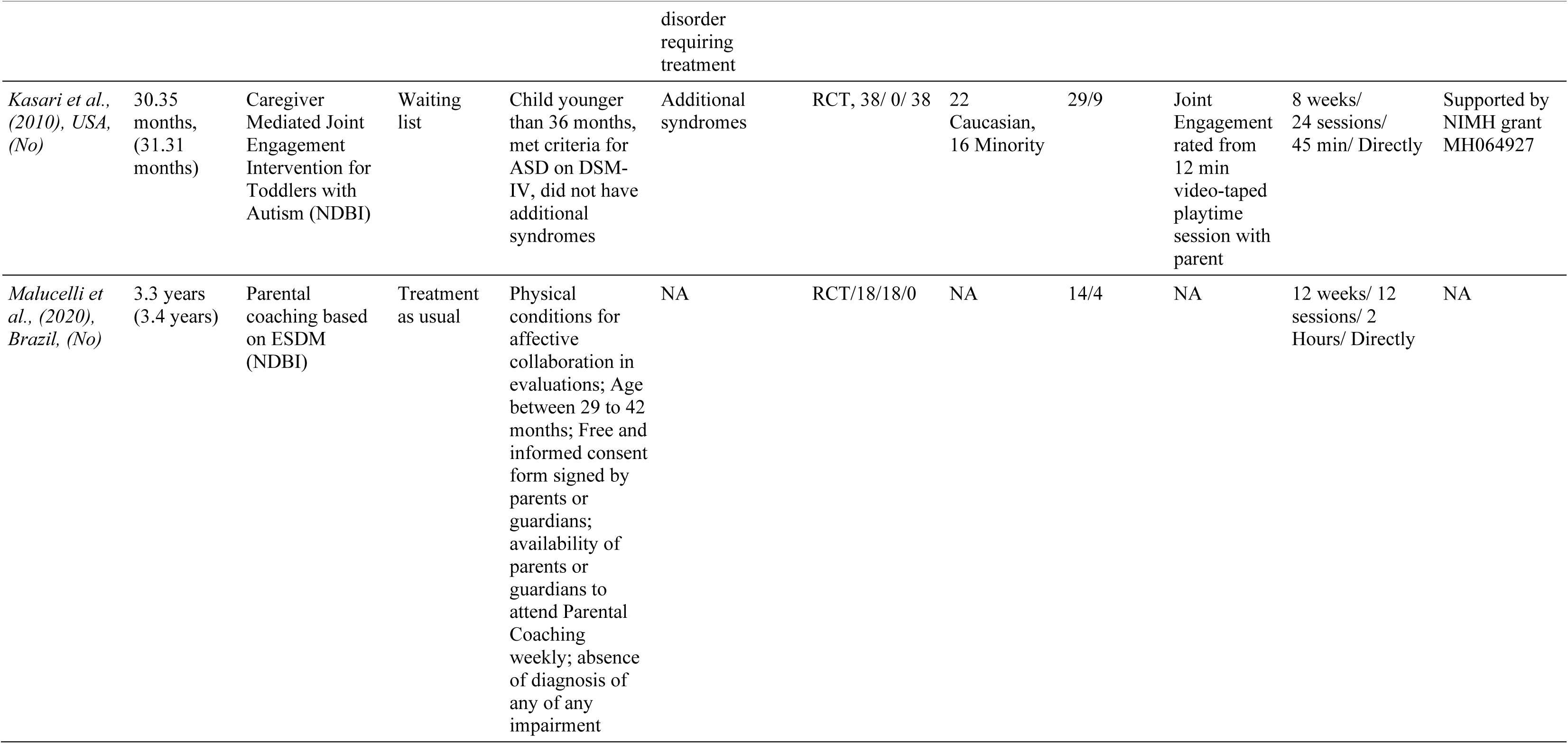
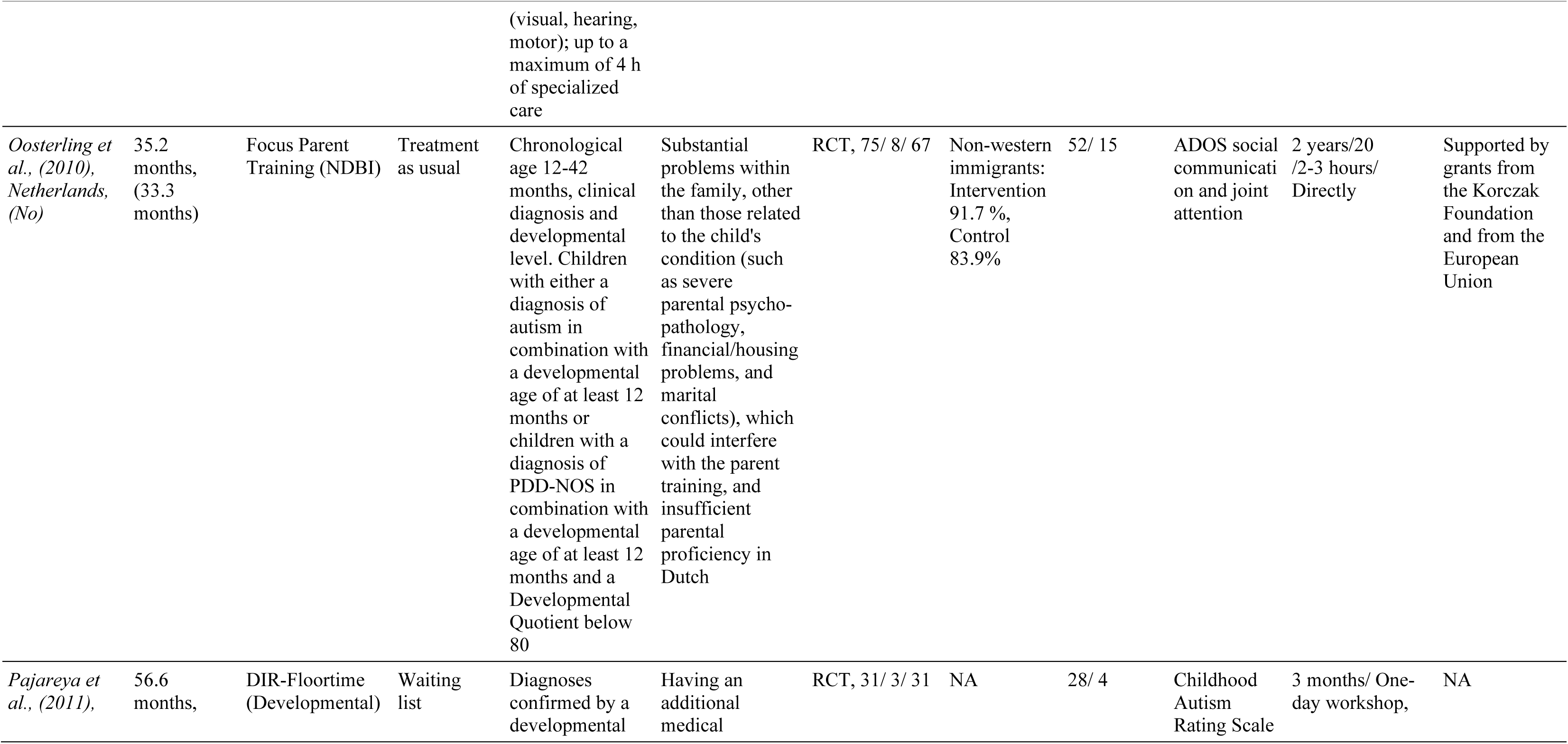
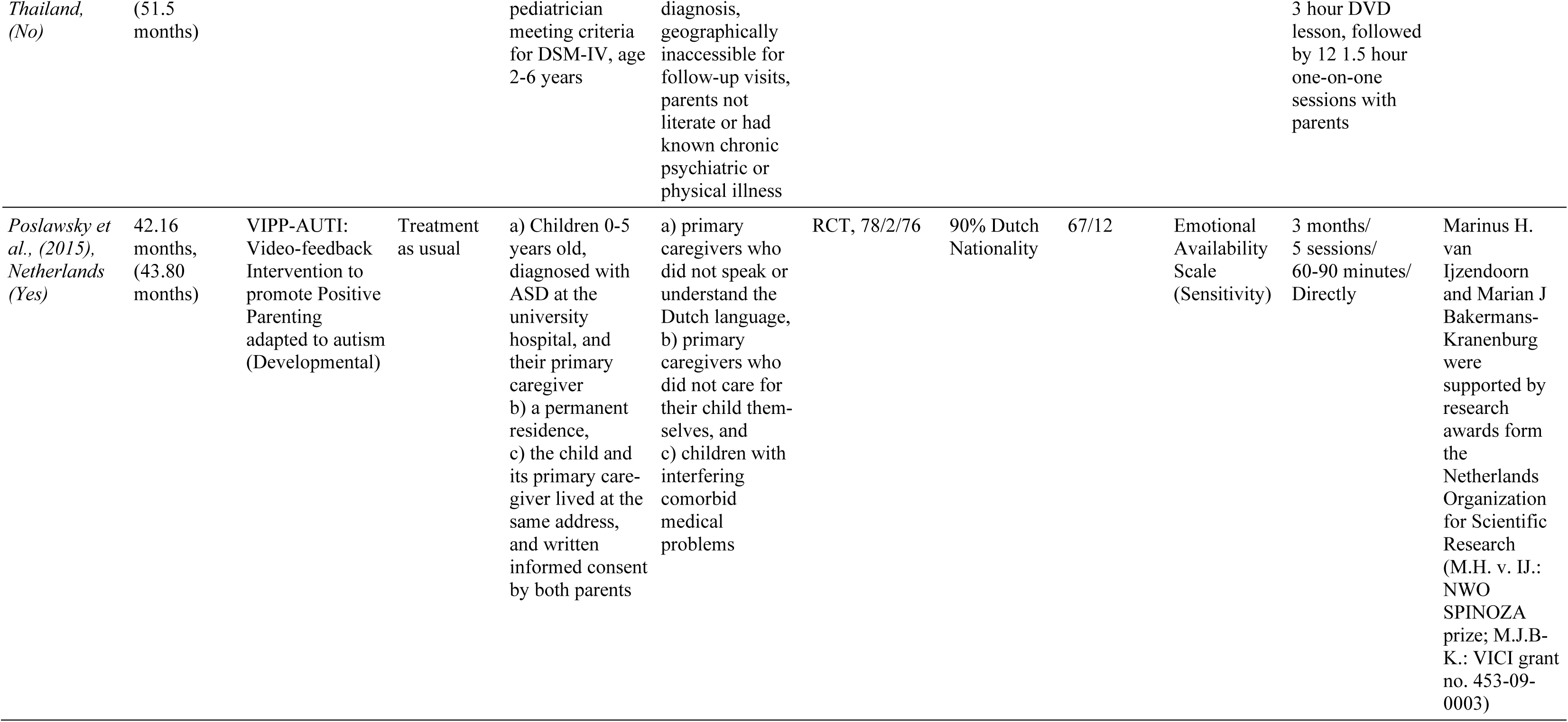
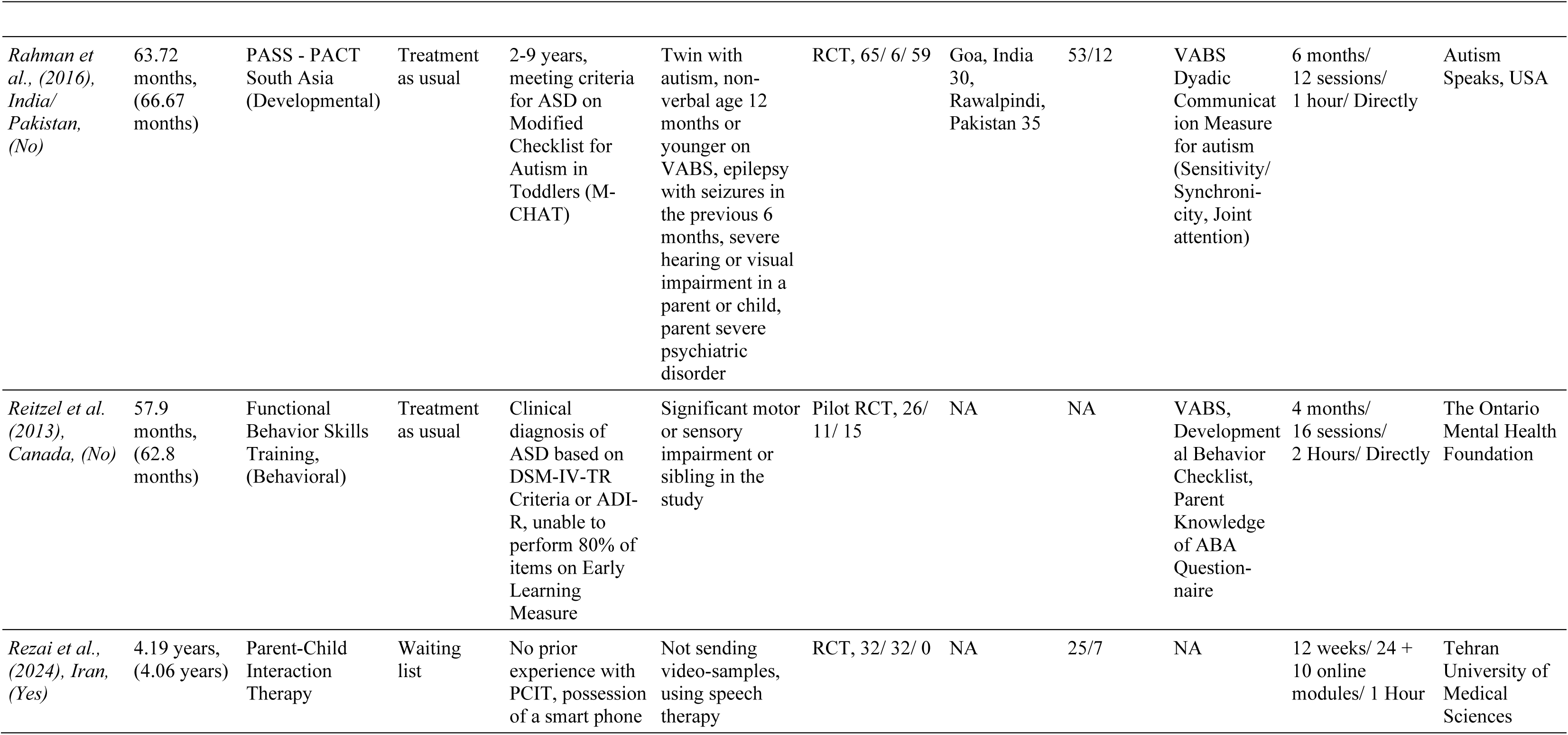
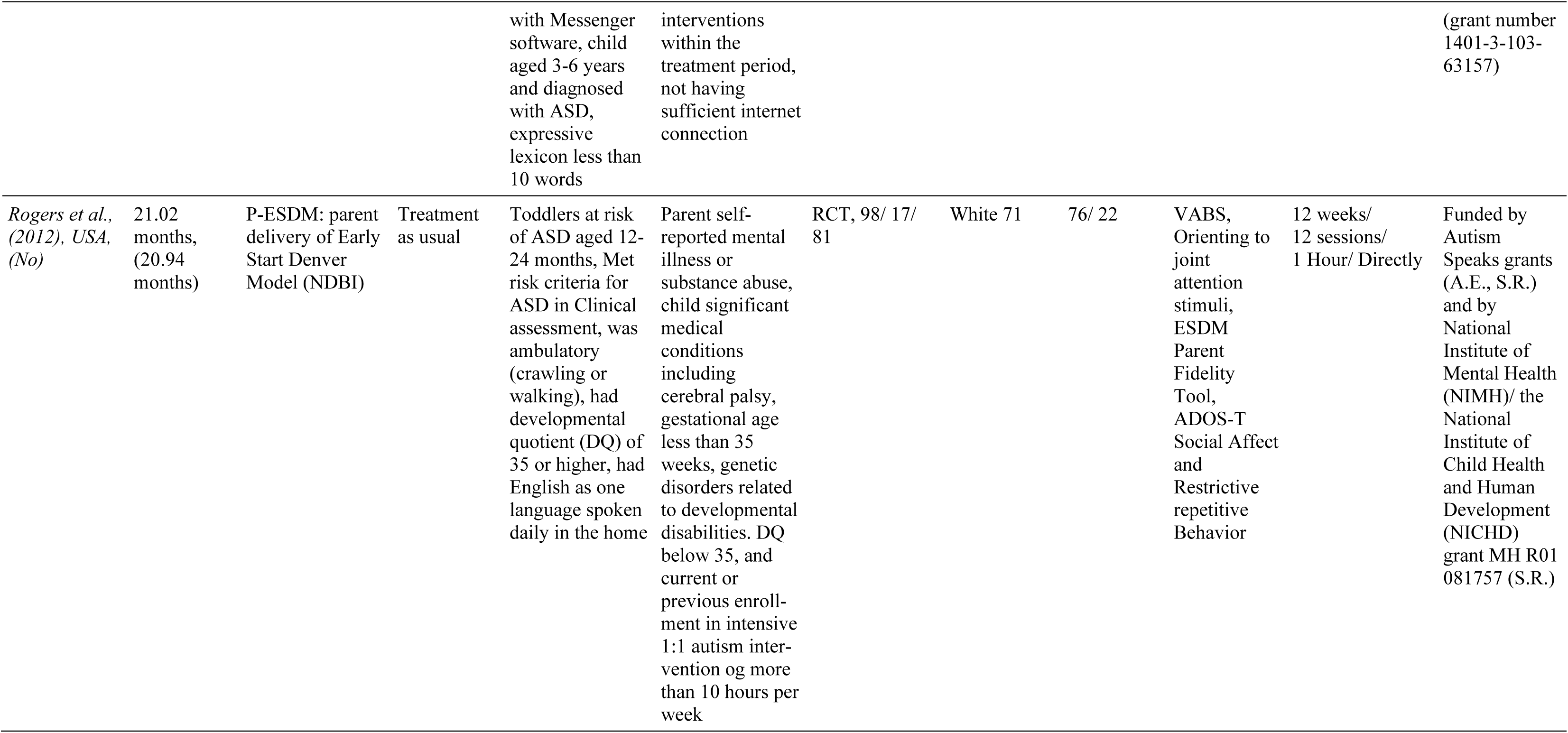
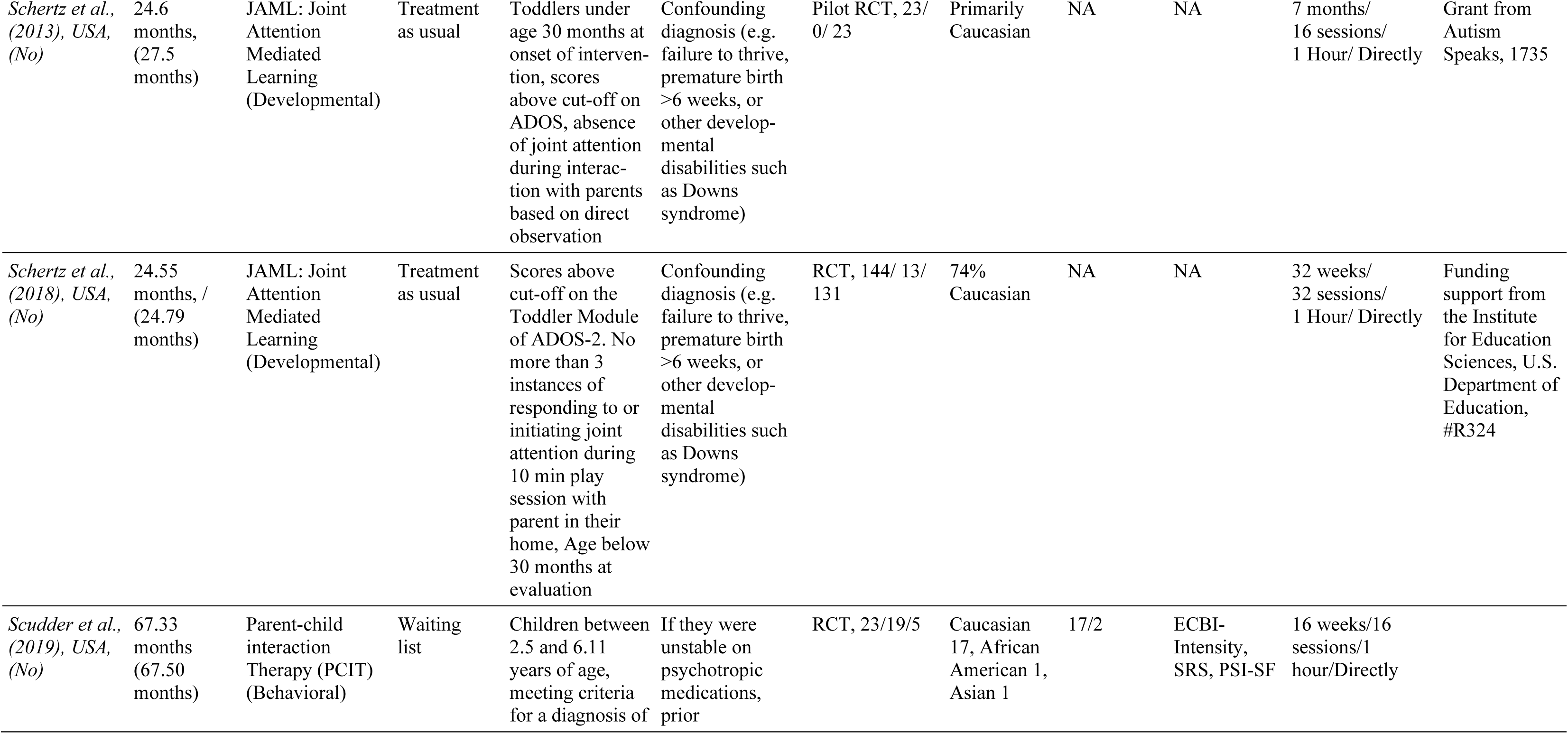
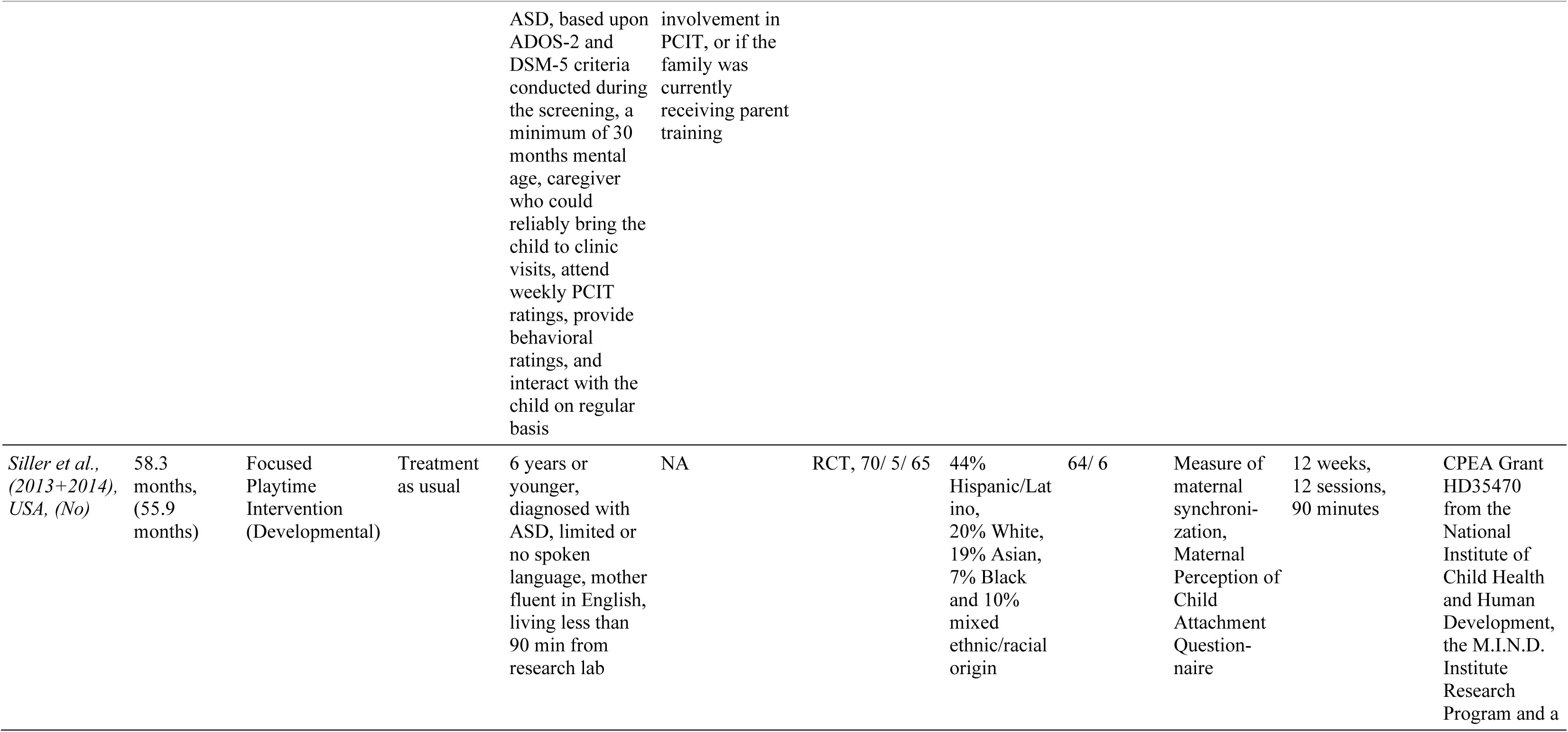
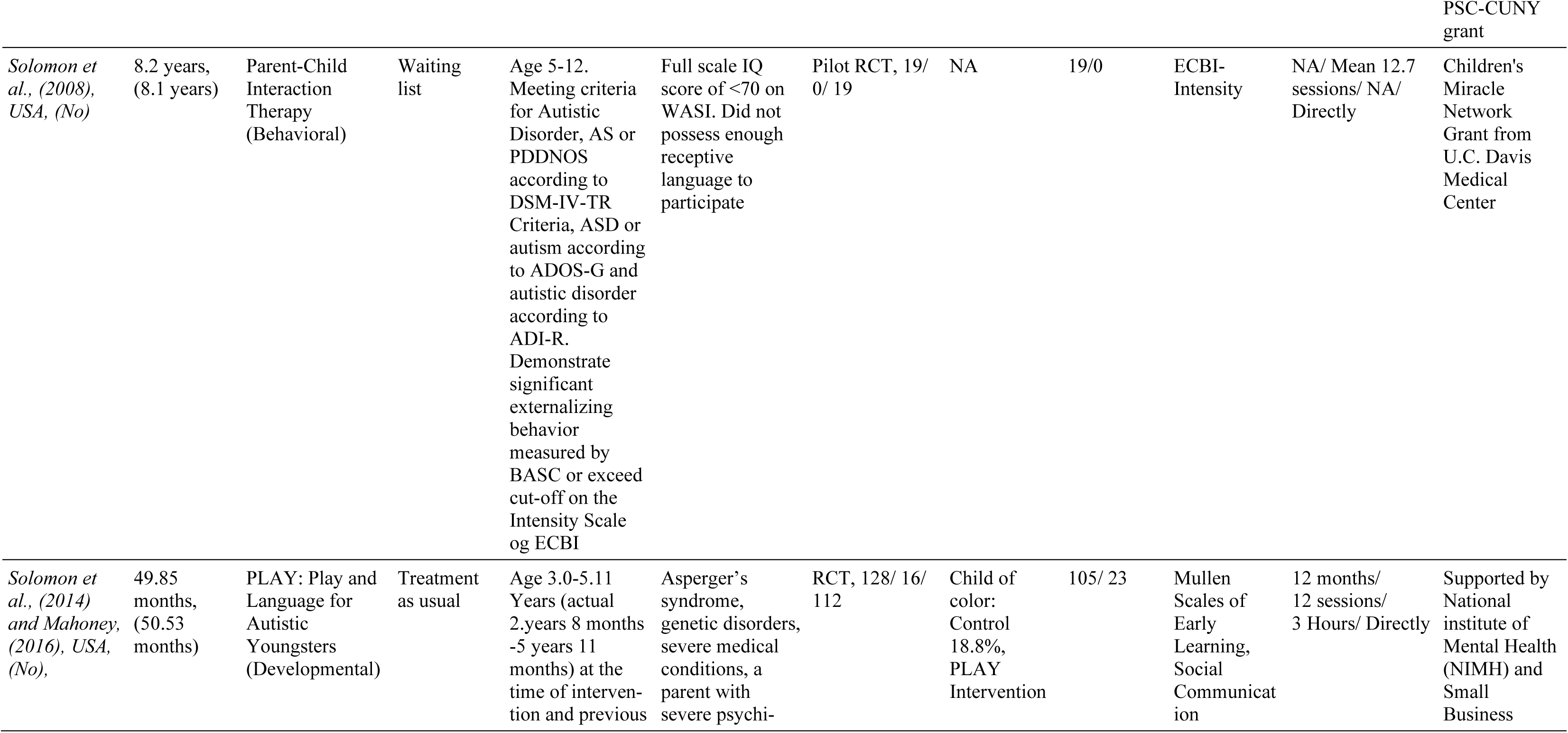
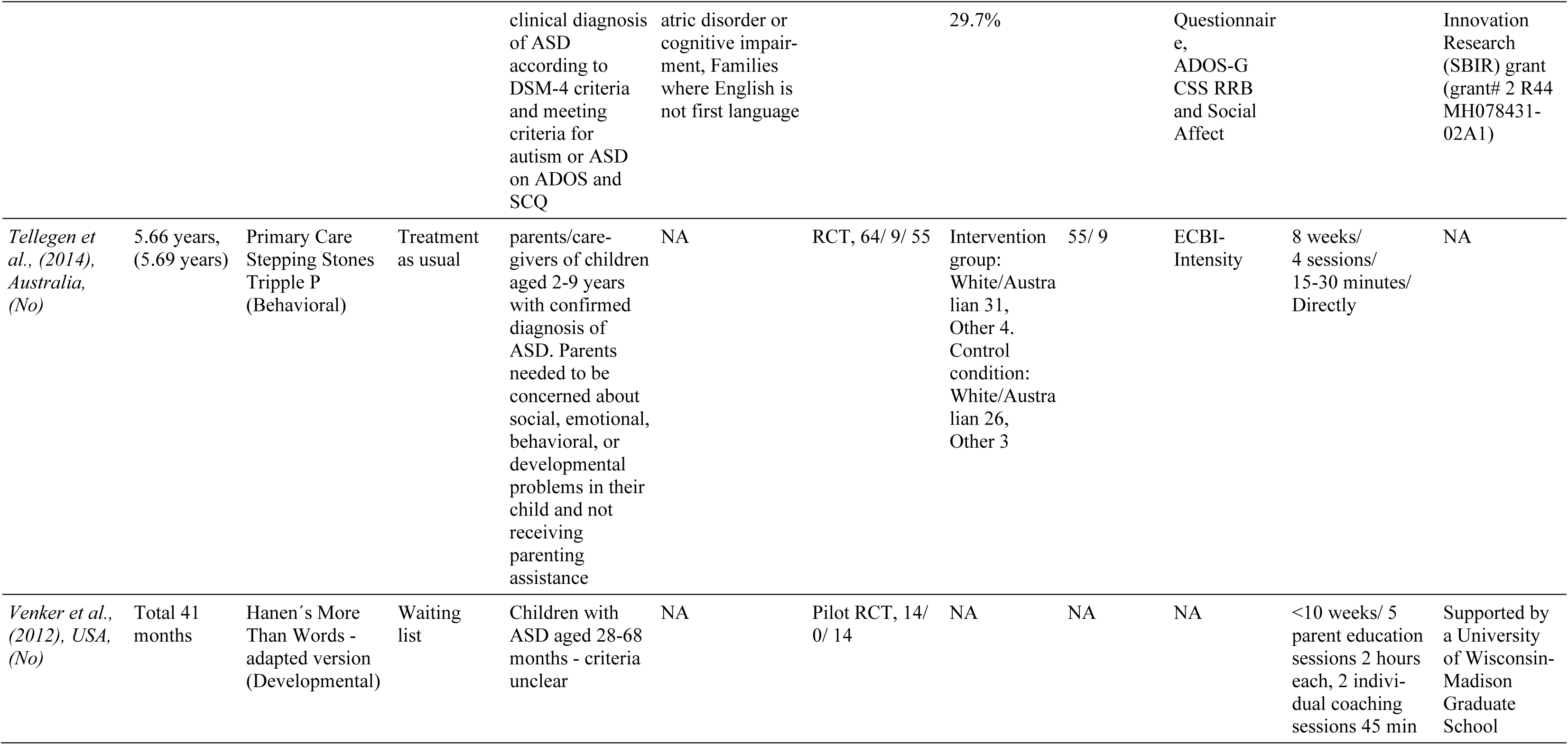
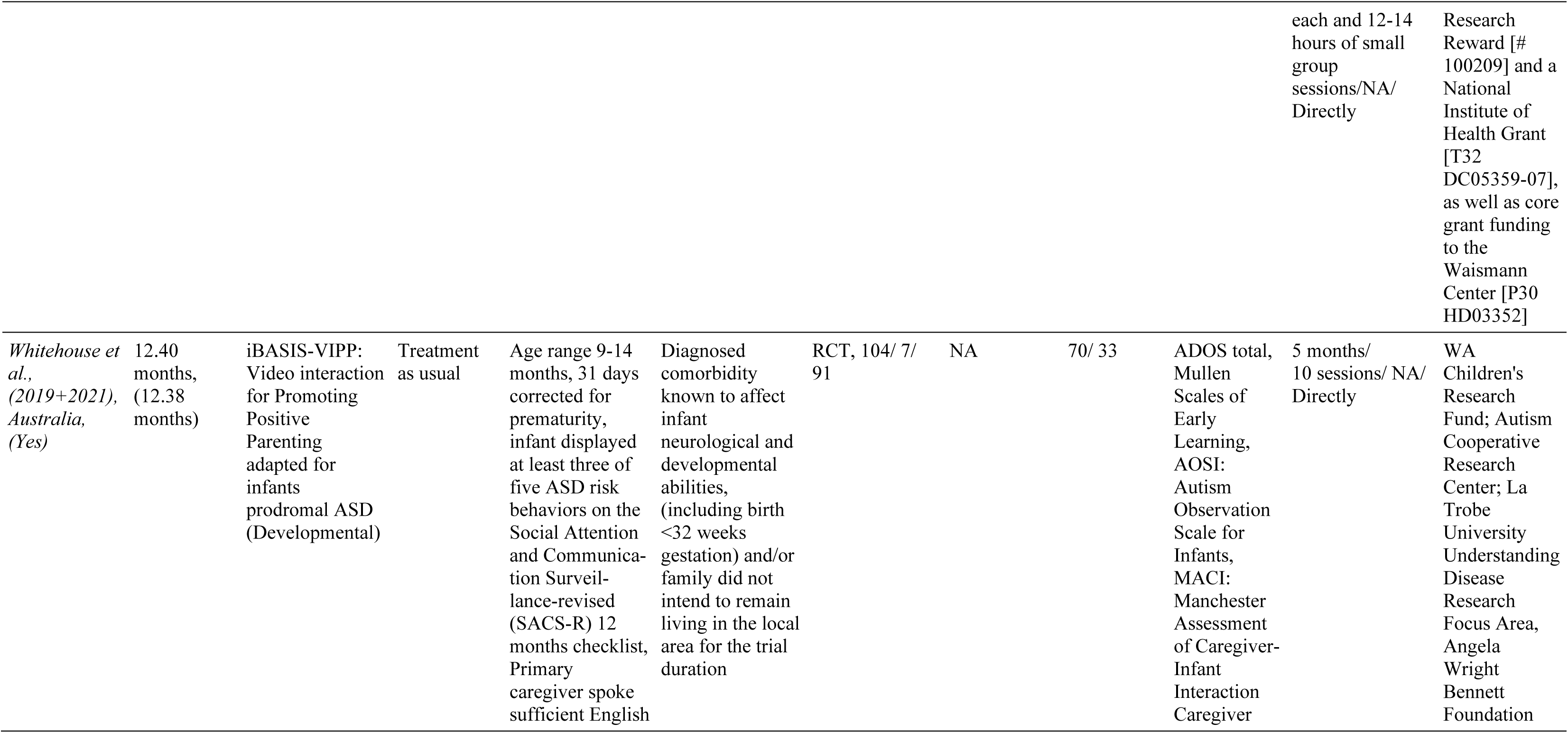
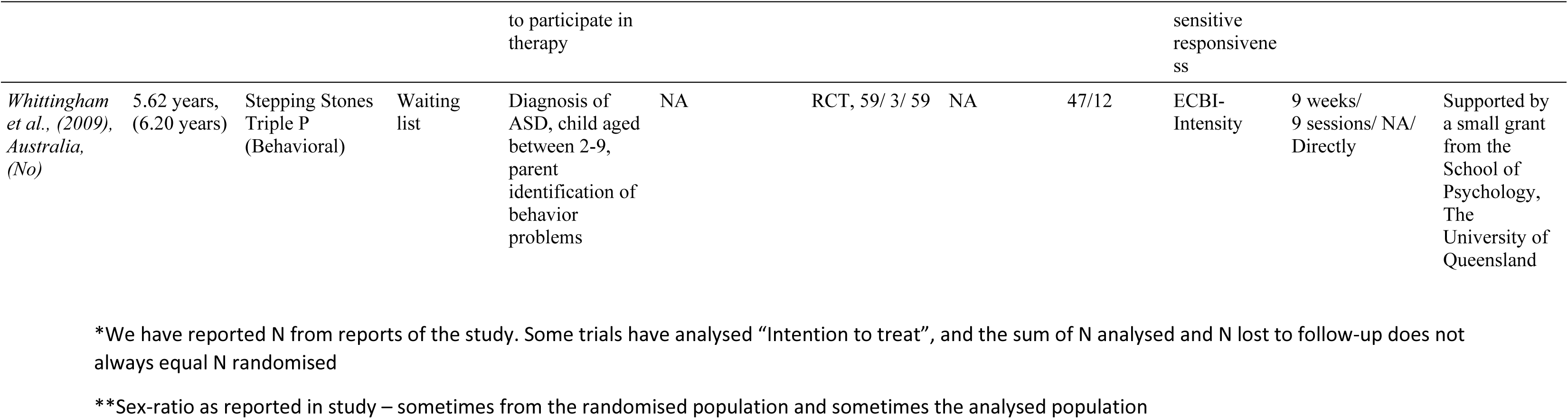
Characteristics table of included studies.

### Excluded Studies

Following the electronic search, a manual search of articles and reference lists and a screening process were conducted (described below in the PRISMA flowchart (Figure 1)). Sixty-eight articles were excluded after examining the full-text manuscripts. For a full list of the excluded studies and reasons for exclusion, see Appendix S4.

**Fig. 1.**
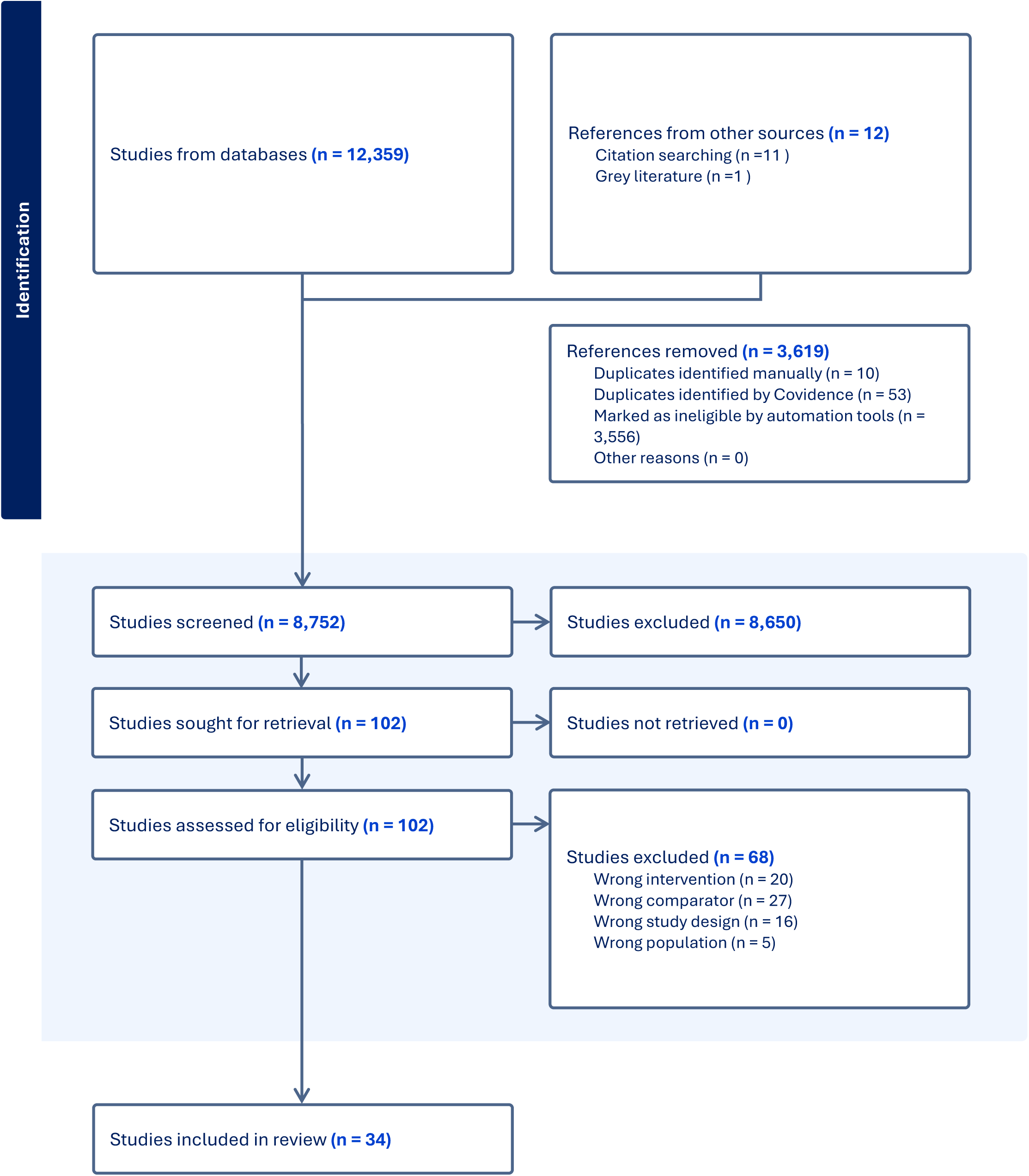
PRISMA Flowchart

### Risk of Bias of Included Studies

All trials were assessed as having an overall high risk of bias. Details on the assessment can be seen in Figure 2.

**Fig. 2.**
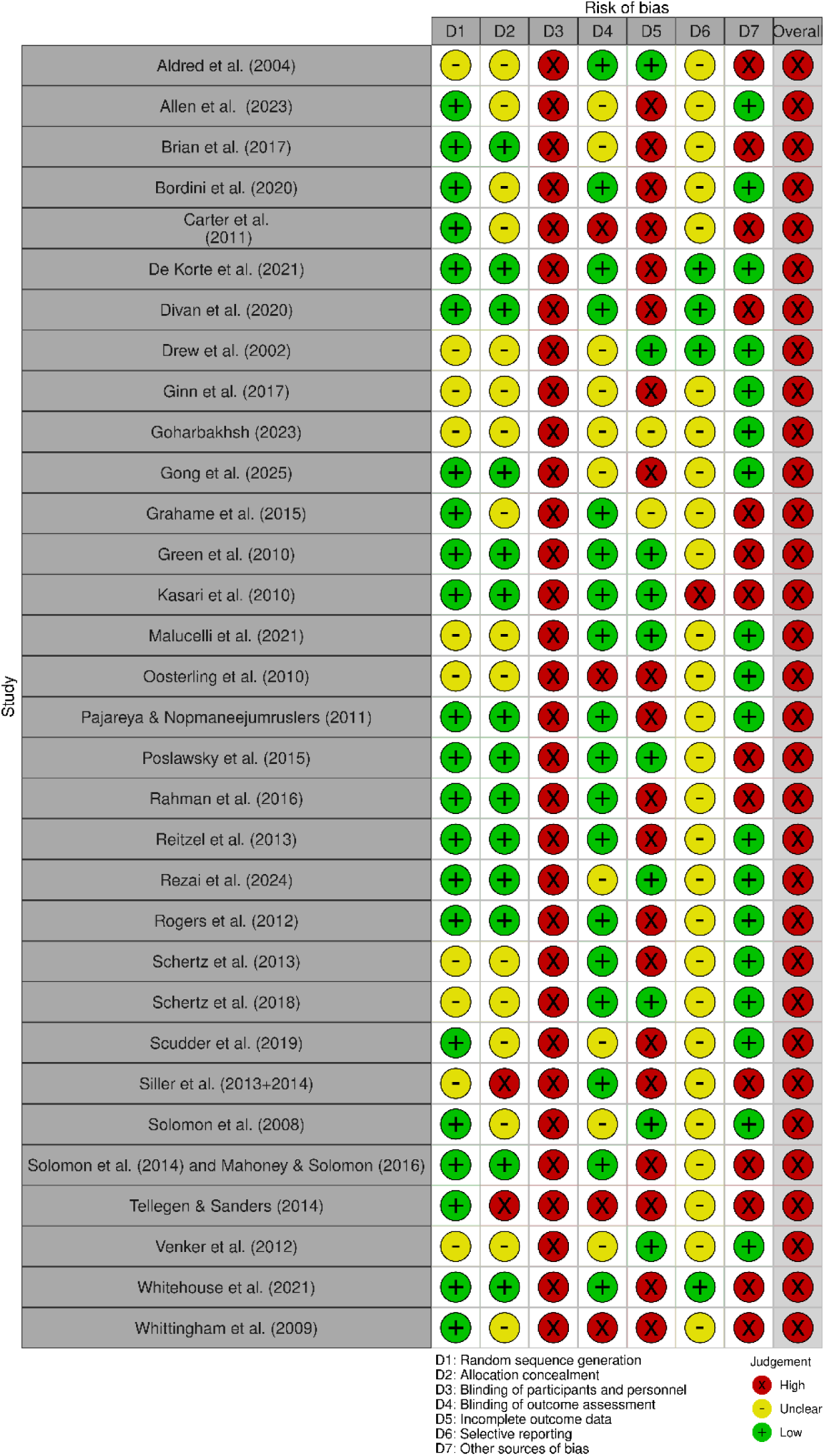
Risk of Bias in included studies

### Primary Outcome

Only four of 32 trials reported results on autism characteristics as measured by the ADOS total score (55–57, 79). All trials assessed the outcomes at the end of the intervention, i.e., 4-13 months after randomisation (Table 1). Both the pilot study by Aldred et al. (2004) and the randomised controlled trial by Green et al. (2010) assessed the effects of Paediatric Autism Communication Therapy (PACT) versus usual care, while Whitehouse et al. (2021) assessed the effects of a similar theory-based intervention for infants, iBASIS-VIPP versus usual care. Gong et al. (2025) assessed the effects of Heart-Mind-Behaviour Parent Training versus waiting list. The meta-analysis showed no evidence of a difference between the PMIs on ADOS total scores compared to usual care (MD −0.88 ADOS total score; 95% confidence interval −2.92 to 1.15; *p* = 0.05). The p-value was over our limit for the significance of 0.013, and the effect size of 0.88 was below our estimated minimal important difference of 3 points (8, 56). Visual inspection of the forest plot (Fig. 3) and statistical tests (*I^2^* = 63.53%) indicated signs of heterogeneity that could not be resolved.

**Fig. 3.**
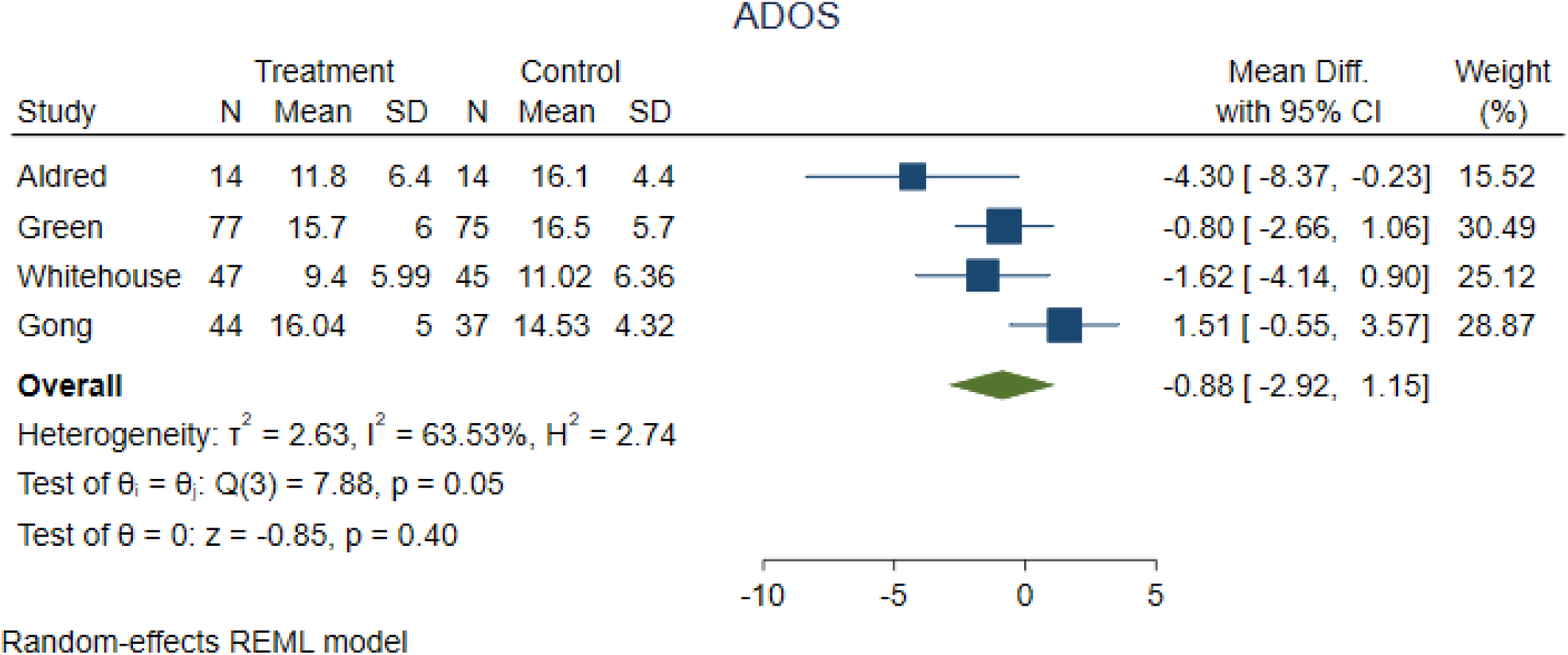
Forest plot autism characteristics as measured by ADOS total score

TSA showed that the meta-analysis was sufficiently powered (Fig. 4).

**Fig. 4.**
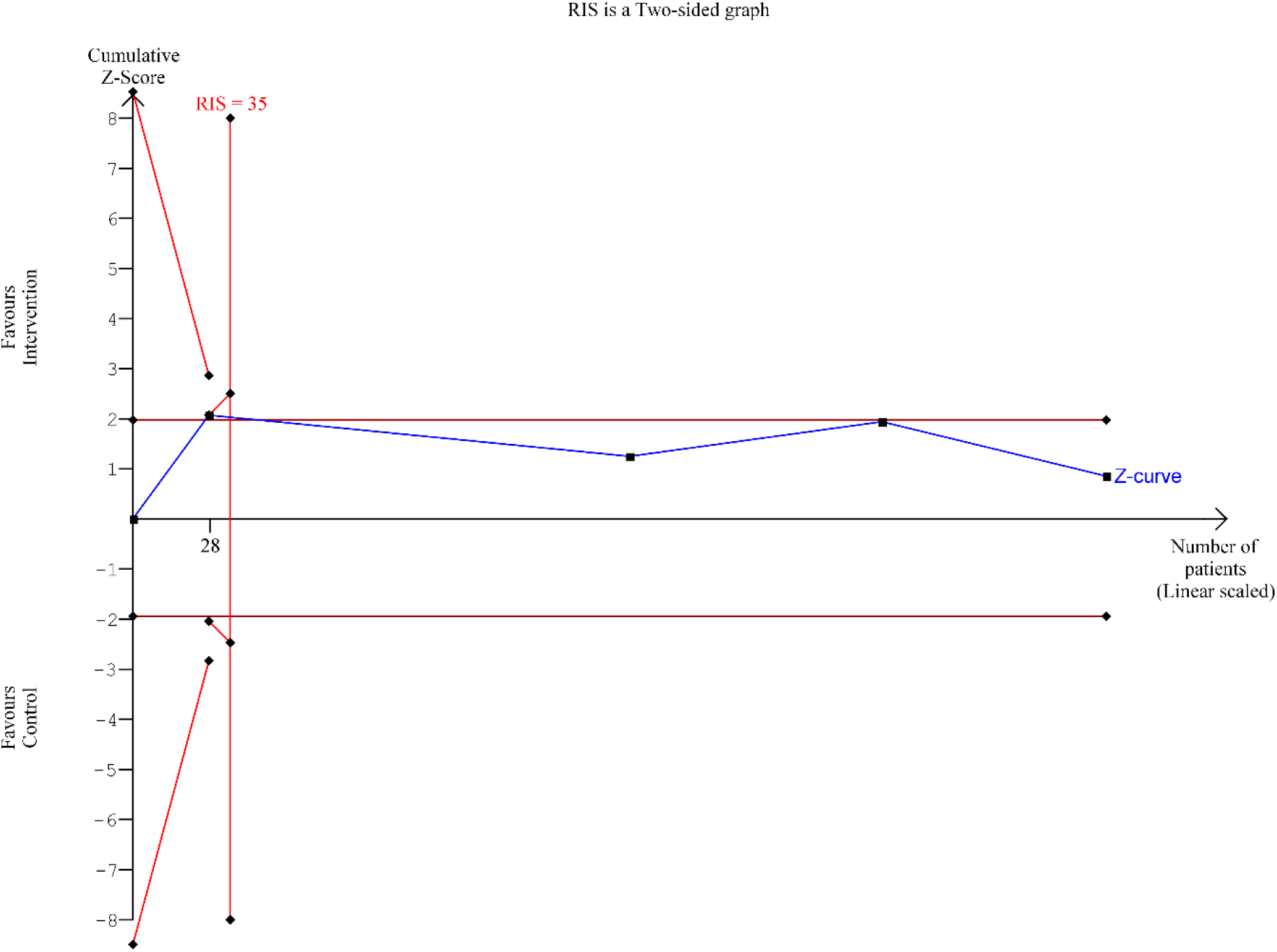
Trial Sequential Analysis of autism characteristics as measured by ADOS total score

This outcome result was assessed as having an overall high risk of bias, and the certainty of evidence was low (Table 2. Summary of Findings (SoF)).

**Table 2.**
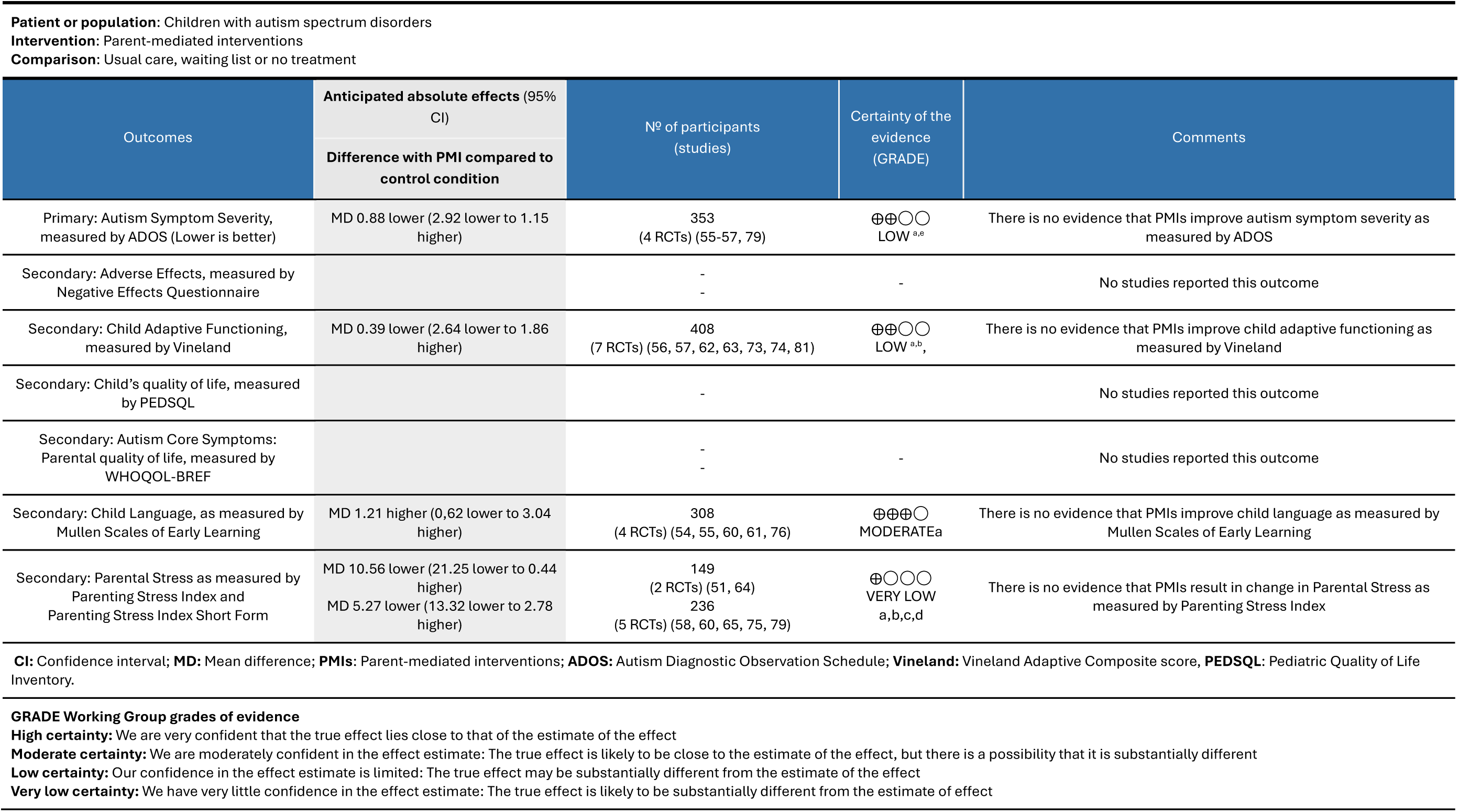
Summary of findings.

Test of interaction comparing the different control groups, i.e., usual care and waiting list showed no evidence of difference (*p* =0.02). When the trials using usual care as the control intervention were analysed separately, meta-analysis still showed no evidence of a difference (MD −1.47; 95% CI −2.88 to −0.07; *p* = 0.04; 3 trials) (55–57). Only one trial used waiting list as the control (79).

None of the remaining predefined subgroup analyses could be performed due to lack of relevant data.

### Secondary Outcomes

#### Child Adaptive Functioning

Seven of 32 trials reported results on child adaptive functioning as measured by the VABS (56, 57, 62, 63, 73, 74, 81). All trials assessed the outcomes at end of the intervention, i.e., 3-13 months after randomisation (Table 1). The meta-analysis showed no evidence of a difference between the PMIs and usual care on adaptive functioning (MD −0.39 Vineland adaptive composite score; 95% confidence interval −2.64 to 1.86; *p* = 0.49). Visual inspection of the forest plot (Appendix S6) and statistical tests (*I^2^* = 0.0%) indicated no clear signs of heterogeneity.

TSA showed that the meta-analysis was sufficiently powered (Figures S6). This outcome result was assessed as an overall high risk of bias, and the certainty of evidence was low (Table 2 (SoF)). Subgroup analysis showed no difference between the PMIs in different intervention groups compared to usual care (*p* = 0.97). Subgroup analysis of younger versus older children showed no difference between the age groups (*p* = 0.67). Subgroup analysis comparing the different groups of autism severity showed no evidence of a difference (*p* = 0.27). None of the remaining predefined subgroup analyses could be performed due to lack of relevant data.

#### Child Language

Only four of 32 trials reported results on child language as measured by MSEL (51, 54, 55, 60, 61). The trials assessed the outcomes at the end of intervention 3-12 months after randomisation (Table 1). The meta-analysis showed no evidence of a difference between the intervention groups and the control groups regarding child language (MD 0.39 MSEL score; 95% confidence interval −1.93 to 2.70; *p* = 0.40). Visual inspection of the forest plot (Appendix S6) and statistical tests (*I^2^* = 17.39%) indicated minor signs of heterogeneity that could not be resolved. TSA showed that the meta-analysis was sufficiently powered (no graph produced). This outcome result was assessed as having an overall high risk of bias, and the certainty of evidence was moderate (Table 2 (SoF)).

Subgroup analysis of the developmental interventions and NDBIs showed no difference between the PMIs in different intervention groups compared to usual care (*p* = 0.53). The subgroup analysis also showed no difference between the different control groups (*p* = 0.28). The remaining predefined subgroup analyses could not be performed due to lack of relevant data.

#### Parental Stress

Only seven of 32 trials reported results on parental stress as measured by the PSI (51, 60, 64, 65). The trials assessed outcomes at the end of the intervention 2.5-12 months after randomisation (Table 1). Five trials assessed the effects of the interventions on parental stress measured by the Parenting Stress Index Short Form (PSI-SF) (58, 60, 65, 75, 79), while two trials measured the effects measured by the full-scale PSI (51, 64). These two different measures were assessed individually in the meta-analysis. Meta-analysis of the full-scale PSI showed no evidence of difference between the intervention groups and the control groups on parental stress (MD −10.55 Parenting Stress Index full scale; 95% confidence interval −21.25 to 0.44; *p* = 0.06). Visual inspection of the forest plots (Appendix S6) and statistical tests (*I^2^* = 0.0%) indicated no clear signs of heterogeneity. TSA showed that the meta-analysis had sufficient power (no graph produced). This outcome result was assessed as having an overall high risk of bias, and certainty of the evidence was very low (Table 2 (SoF)). The meta-analysis of the PSI-SF showed no evidence of a difference between the PMIs and control groups on parental stress (MD −5.27 Parenting Stress Index Short Form; 95% confidence interval −13.32 to 2.78; *p* = 0.20). Visual inspection of the forest plot (Appendix S6) and statistical tests (*I^2^* = 48.53%) indicated signs of heterogeneity that could not be resolved. TSA showed that the meta-analysis had sufficient power (no graph produced). Outcome result was assessed as an overall high risk of bias, and the certainty of evidence was very low (Table 2 (SoF))

None of the predefined subgroup analyses could be performed due to a lack of relevant data. None of the included trials assessed the secondary outcomes: negative effects as assessed by the NEQ, child quality of life as assessed by the PEDSQL, or parental quality of life as assessed by the WHOQL-BREF.

All reported results are assessed with random-effects meta-analyses (37) having the most conservative result. Results of the fixed-effects meta-analyses (38) can be seen in Appendix S6.

### Exploratory Outcomes

#### Child Behaviour Problems

Seven of 32 trials reported results on child behaviour problems (58, 65, 75–78, 81). All trials assessed outcomes at the end of the intervention, between 2 and 4 months after randomisation (Table 1). The meta-analyses provided evidence of a beneficial effect of the PMIs on child behaviour problems (standardised mean difference (SMD) −0.81; 95% confidence interval - 1.27 to −0.36; *p* = 0.01). Visual inspection of the forest plot (Appendix S7) and statistical tests (*I^2^* = 63.97%) indicated signs of heterogeneity that could not be resolved. This outcome result was assessed as having an overall high risk of bias, and the certainty of evidence was low (Table 3).

**Table 3.**
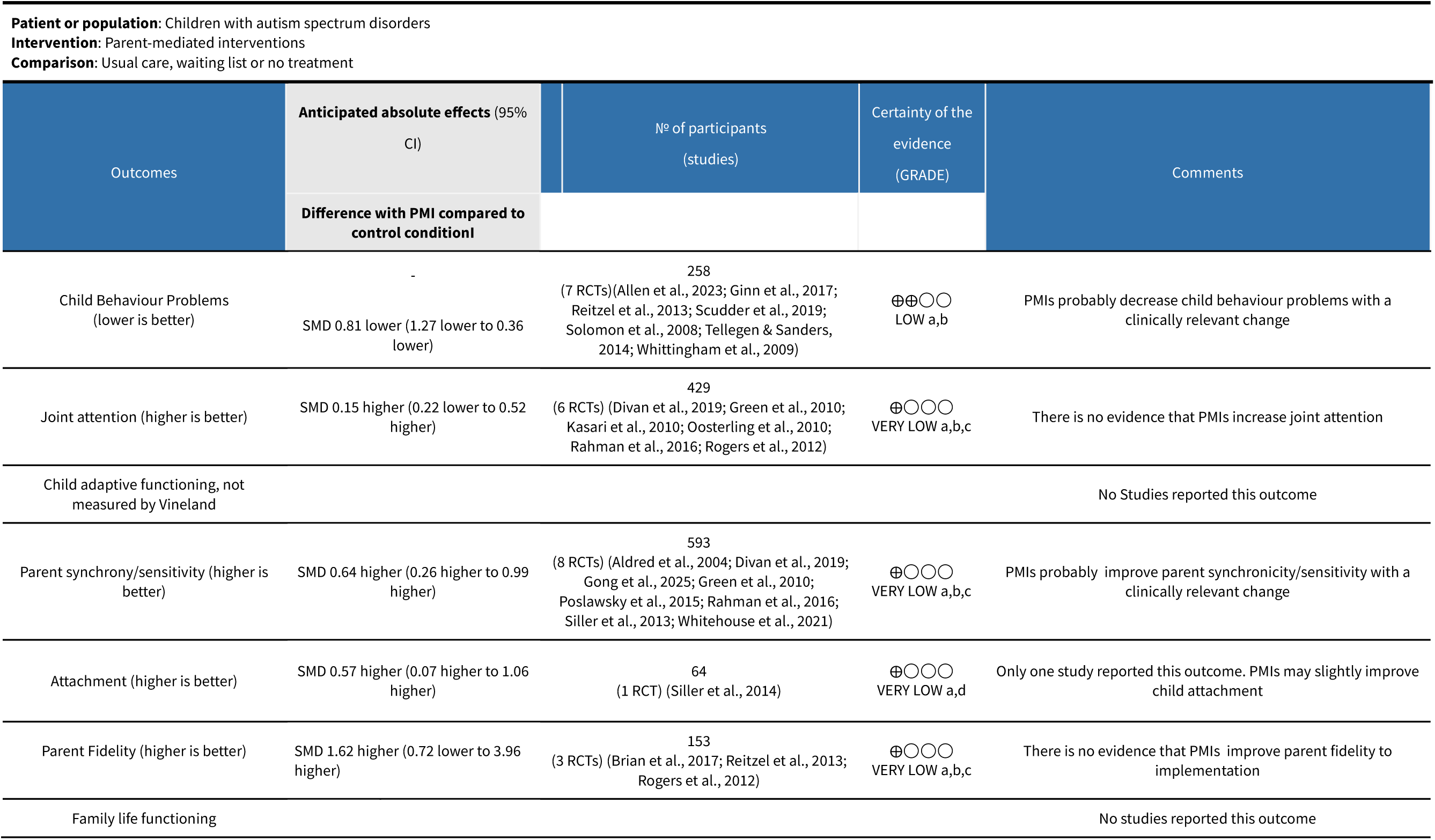
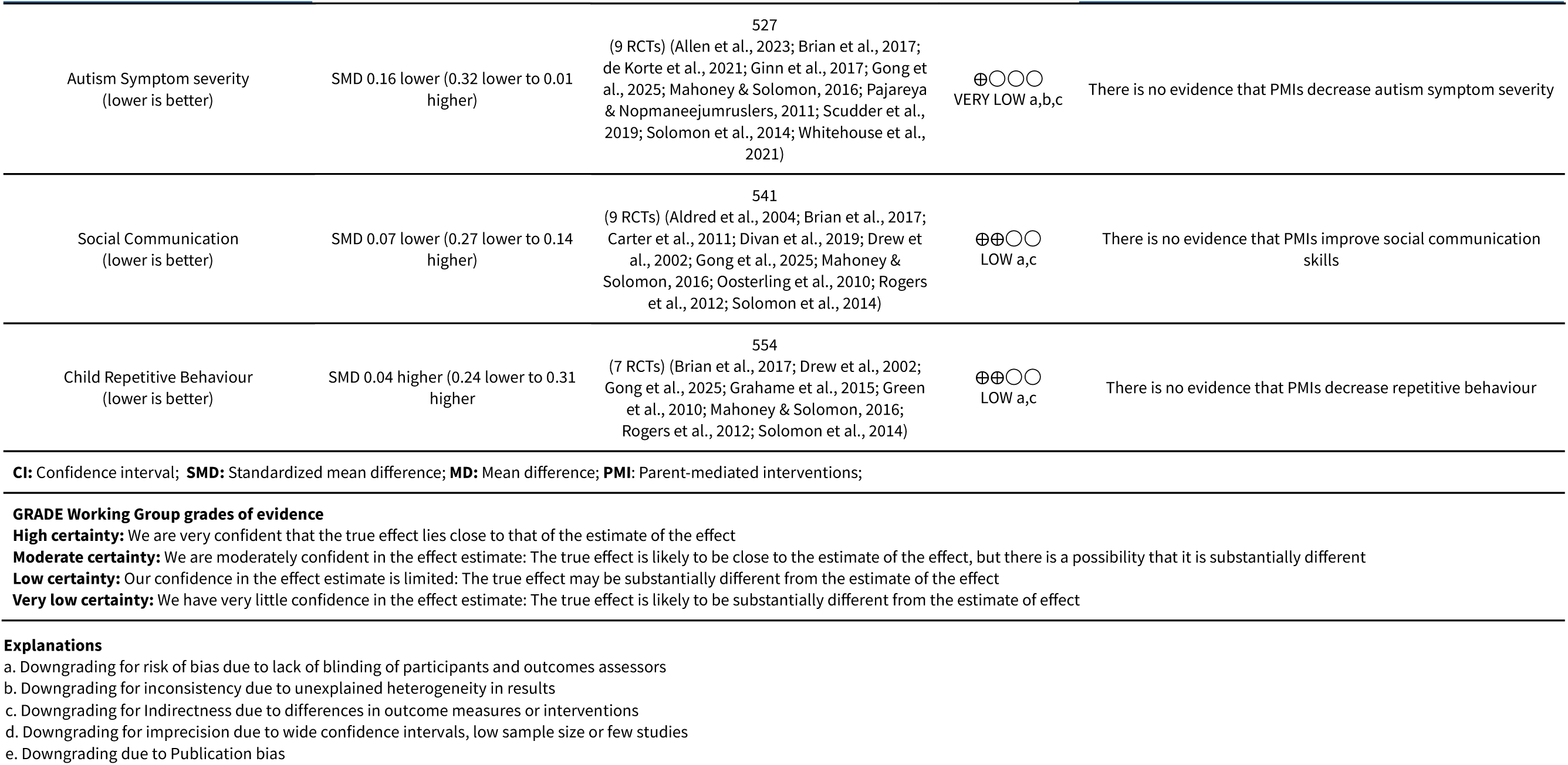
Summary of findings Exploratory outcomes:

| Outcomes | Anticipated absolute effects (95% CI) | Nº of participants (studies) | Certainty of the evidence (GRADE) | Comments |
| --- | --- | --- | --- | --- |
|  | Difference with PMI compared to control condition<sup>1</sup> |  |  |  |
| Child Behaviour Problems (lower is better) | -<br>SMD 0.81 lower (1.27 lower to 0.36 lower) | 258<br>(7 RCTs) (Allen et al., 2023; Ginn et al., 2017; Reitzel et al., 2013; Scudder et al., 2019; Solomon et al., 2008; Tellegen & Sanders, 2014; Whittingham et al., 2009) | ⊕⊕○○<br>LOW a,b | PMIs probably decrease child behaviour problems with a clinically relevant change |
| Joint attention (higher is better) | SMD 0.15 higher (0.22 lower to 0.52 higher) | 429<br>(6 RCTs) (Divan et al., 2019; Green et al., 2010; Kasari et al., 2010; Oosterling et al., 2010; Rahman et al., 2016; Rogers et al., 2012) | ⊕○○○<br>VERY LOW a,b,c | There is no evidence that PMIs increase joint attention |
| Child adaptive functioning, not measured by Vineland |  |  |  | No Studies reported this outcome |
| Parent synchrony/sensitivity (higher is better) | SMD 0.64 higher (0.26 higher to 0.99 higher) | 593<br>(8 RCTs) (Aldred et al., 2004; Divan et al., 2019; Gong et al., 2025; Green et al., 2010; Poslawsky et al., 2015; Rahman et al., 2016; Siller et al., 2013; Whitehouse et al., 2021) | ⊕○○○<br>VERY LOW a,b,c | PMIs probably improve parent synchronicity/sensitivity with a clinically relevant change |
| Attachment (higher is better) | SMD 0.57 higher (0.07 higher to 1.06 higher) | 64<br>(1 RCT) (Siller et al., 2014) | ⊕○○○<br>VERY LOW a,d | Only one study reported this outcome. PMIs may slightly improve child attachment |
| Parent Fidelity (higher is better) | SMD 1.62 higher (0.72 lower to 3.96 higher) | 153<br>(3 RCTs) (Brian et al., 2017; Reitzel et al., 2013; Rogers et al., 2012) | ⊕○○○<br>VERY LOW a,b,c | There is no evidence that PMIs improve parent fidelity to implementation |
| Family life functioning |  |  |  | No studies reported this outcome |
|  | Difference with PMI compared to control condition<sup>1</sup> |  |  |  |
| Autism Symptom severity (lower is better) | SMD 0.16 lower (0.32 lower to 0.01 higher) | 527<br>(9 RCTs) (Allen et al., 2023; Brian et al., 2017; de Korte et al., 2021; Ginn et al., 2017; Gong et al., 2025; Mahoney & Solomon, 2016; Pajareya & Nopmaneejumrulers, 2011; Scudder et al., 2019; Solomon et al., 2014; Whitehouse et al., 2021) | ⊕○○○<br>VERY LOW a,b,c | There is no evidence that PMIs decrease autism symptom severity |
| Social Communication (lower is better) | SMD 0.07 lower (0.27 lower to 0.14 higher) | 541<br>(9 RCTs) (Aldred et al., 2004; Brian et al., 2017; Carter et al., 2011; Divan et al., 2019; Drew et al., 2002; Gong et al., 2025; Mahoney & Solomon, 2016; Oosterling et al., 2010; Rogers et al., 2012; Solomon et al., 2014) | ⊕⊕○○<br>LOW a,c | There is no evidence that PMIs improve social communication skills |
| Child Repetitive Behaviour (lower is better) | SMD 0.04 higher (0.24 lower to 0.31 higher) | 554<br>(7 RCTs) (Brian et al., 2017; Drew et al., 2002; Gong et al., 2025; Grahame et al., 2015; Green et al., 2010; Mahoney & Solomon, 2016; Rogers et al., 2012; Solomon et al., 2014) | ⊕⊕○○<br>LOW a,c | There is no evidence that PMIs decrease repetitive behaviour |

**CI:** Confidence interval; **SMD:** Standardized mean difference; **MD:** Mean difference; **PMI:** Parent-mediated interventions;

#### Joint Attention

Six of 32 trials reported results on the exploratory outcome of joint attention (56, 63, 68, 70, 73, 74). All trials assessed the outcomes at the end of the intervention, 6-15 months after randomisation (Table 1). The meta-analysis showed no evidence of a difference between PMIs and control conditions on joint attention (SMD 0.15; 95% confidence interval −0.22 to 0.52; *p* = 0.43). Visual inspection of the forest plot (Appendix S7) and statistical tests (I^2^ = 71.03%) indicated signs of heterogeneity that could not be resolved. This outcome result was assessed as having an overall high risk of bias, and the certainty of evidence was very low (Table 3).

#### Parent sensitivity/synchronicity

Eight of 32 interventions reported results on the exploratory outcome of parent sensitivity/synchronicity (53, 55–57, 63, 72, 73, 79). All trials assessed outcomes at the end of the intervention, 3-13 months after randomisation (Table 1). Meta-analysis showed evidence of a beneficial effect of the PMIs on parent sensitivity/synchronicity (SMD 0.64; 95% confidence interval 0.26 to 0.99; *p* < 0.01). Visual inspection of the forest plot (Appendix S7) and statistical tests (*I^2^* = 76.41%) indicated signs of heterogeneity that could not be resolved. This outcome result was assessed as having an overall high risk of bias, and the certainty of evidence was very low (Table 3).

#### Attachment

Only one of 32 trials reported results on attachment (52). The trial assessed outcomes at the end of the intervention three months after randomisation. The trial by Siller et al. (2014) assessed the effect of Focused Playtime Intervention versus usual care (Table 1). No meta-analysis was performed due to a lack of studies. The trial showed evidence of a beneficial effect of the PMIs Focused Playtime intervention on child attachment compared to management as usual on child attachment, as measured by the Maternal Perception of Child Attachment questionnaire with a significant main effect of intervention group allocation on gains in observed attachment behaviours (mean), t(54) = 2.0, *p* < 0.05 (52).

#### Parent Fidelity to Intervention Implementation

Only three of 32 trials reported results on parent fidelity (60, 74, 81). All trials assessed outcomes at the end of the intervention, 3-4 months after randomisation (Table 1). The meta-analysis showed no evidence of a difference between PMIs and usual care or waiting list on parent fidelity (SMD 1.62; 95% confidence interval −0.72 to 3.96; *p* = 0.18). Visual inspection of the forest plot (Appendix S7) and statistical tests (*I^2^* = 96.38%) indicated signs of heterogeneity that could not be resolved. This outcome result was assessed as having an overall high risk of bias, and the certainty of evidence was very low (Table 3).

#### Autism Characteristics

Nine of 32 trials reported results on autism characteristics (not measured by the ADOS total score) (51, 54, 55, 60, 62, 65, 71, 79). All trials assessed outcomes at the end of the intervention, 3-14 months after randomisation (Table 1). The meta-analysis showed no evidence of a difference between the intervention groups and control conditions on autism characteristics (not measured by the ADOS total score) (SMD −0.16; 95% confidence interval −0.32 to 0.01; *p* = 0.07). Visual inspection of the forest plot (Appendix S7) and statistical tests (*I^2^* =0.00%) indicated no signs of heterogeneity. This outcome result was assessed as having an overall high risk of bias, and the certainty of evidence was very low (Table 3).

#### Child Social Communication

Nine of 32 trials reported results on the exploratory outcome of child social communication features (51, 54, 60, 61, 63, 64, 70, 74, 79). All trials assessed outcomes at the end of the intervention, 3-15 months after randomisation (Table 1). The meta-analysis showed no evidence of a difference between PMIs and control conditions on child social communication features (SMD −0.07; 95% confidence interval −0.27 to 0.14; *p* = 0.15). Visual inspection of the forest plot (Appendix S7) and statistical tests (*I^2^* = 28.52%) indicated signs of heterogeneity that could not be resolved. This outcome result was assessed as having an overall high risk of bias, and the certainty of evidence was low (Table 3).

#### Child Repetitive Behaviour

Seven of 32 trials reported results on child repetitive behaviour (51, 54, 56, 57, 60, 67, 74, 79). All trials assessed outcomes at the end of the intervention, 8 weeks-14 months after randomisation (Table 1). The meta-analysis showed no evidence of a difference between PMIs and control conditions on child repetitive behaviour (SMD 0.04; 95% confidence interval −0.24 to 0.31; *p* = 0.80). Visual inspection of the forest plot (Appendix S7) and statistical tests (*I^2^* = 59.47%) indicated signs of heterogeneity that could not be resolved. This outcome result was assessed as having an overall high risk of bias, and the certainty of evidence was low (Table 3).

All sub-group analyses of exploratory outcomes can be found in Appendix S7. None of the included trials assessed the exploratory outcomes of child adaptive functioning (not measured by the Vineland Adaptive Behavior Scales) or family life functioning.

## Discussion

The objective of this review was to evaluate the positive and adverse effects of PMIs versus care as usual, waiting list, or no intervention for children with autism and their parents. Thirty-two randomised clinical trials were included.

Regarding our primary outcome, both the meta-analysis and TSA demonstrated that PMIs did not lead to a significant reduction in autism characteristics as measured by the ADOS total score, and TSA confirmed that the meta-analysis had sufficient statistical power. The point estimate indicated that the effect size fell below the pre-specified minimal important difference.

Meta-analysis and TSA additionally showed that we could reject that PMIs improved child adaptive functioning, child language and parental stress based on our predefined anticipated intervention effects and the selected outcome measures. None of the included trials investigated secondary outcomes of adverse effects or child/parental quality of life as measured by the assessment tools chosen for the analysis. We did not miss any substantial secondary outcome results by limiting our outcome assessments to only one assessment tool for these outcomes, as we did not find other measures of negative effects or child or parental quality of life when extracting the data from the articles. None of the sub-group analyses concerning primary and secondary outcomes showed any differences between groups.

These conclusions were based on a strict methodology of a predefined p-value of 0.013 and the minimal important differences. There is uncertainty in determining a minimal important difference, and the “true” value may be lower, or the minimal important difference may be unknown for the ADOS and the other included measures.

The lack of data measuring adverse effects or the child and parental quality of life could be important for future research. The negative effects of an intervention should always be monitored, and when the benefits of the intervention are still questionable, it may be even more crucial to assess the negative effects as they could outweigh the benefits. Within PMIs, child and parental quality of life may be meaningful outcomes for participants.

With respect to our primary and secondary outcomes, our findings could not confirm results from previous reviews indicating evidence of a slight reduction in autism symptom severity (13, 15). One previous review (85) found some benefits of PMIs regarding parental stress, which could not be confirmed in either our meta-analyses or the Cochrane review by Oono et al. (2013).

In terms of exploratory outcomes, meta-analyses showed a beneficial effect of PMIs on child behaviour problems (*p* = 0.01) and parental sensitivity/synchronicity (*p* < 0.01). Regarding the remaining exploratory outcomes, i.e., joint attention, parent fidelity to intervention implementation, autism symptom severity (not measured by ADOS total score), child social communication symptoms, and child repetitive behaviour meta-analyses, we did not find evidence of a difference between the intervention groups and control groups.

The finding that PMIs reduce child behaviour problems confirms and strengthens findings from previous reviews (14, 19–21). The result of PMIs improving parental sensitivity/synchronicity opens the possibility of further investigation of related outcomes such as child attachment.

## Strengths

The present review has several strengths. The methodology was predefined and described in detail in our published protocol (8). The risks of bias in the trials were assessed following instructions from the *Cochrane Handbook for Systematic Reviews of Interventions* (31). Statistical heterogeneity was assessed by the chi^2^ test (threshold *p* < 0.10), and the quantities of heterogeneity were assessed by the *I^2^* statistic (31–33). TSA was performed to calculate the required information size and minimise risk of type I or type II errors. Predefining the minimal important difference in our protocol ensured an objective assessment of the clinical importance of the findings. We used a strict inclusion criterion of what is considered a PMI to ensure that the trial interventions were verified as parent-mediated. We limited our ability to assess primary and secondary outcomes by defining one assessment measure for each outcome, ensuring a stronger result when calculating the MD and not the SMD.

## Limitations

The review has several limitations. All trials were at high risk of bias, thus creating a risk of overestimating or underestimating the beneficial and adverse effects of PMIs. The included studies did not include any assessments of the adverse effects of PMIs. When only assessing one outcome measure for each primary and secondary outcome, relevant results from some of the trials were potentially missed. It was evident that there were large differences in which assessment tools were used by the different studies to measure the same outcomes. The studies in this field lack consistency in the choice of outcome measures, and a maximum of eight studies were included in any of the primary and secondary meta-analyses. A potential methodological issue is that the assessment tools are administered differently. For example, VABS is administered as a structured interview in some studies, while in others, it is a self-administered questionnaire. Additionally, some assessment tools are rated by clinicians and others by parents.

The findings were based on studies all assessed as having a high risk of bias and with the certainty of evidence rated as low or very low for all our primary and secondary outcomes. Several studies were small and of low methodological quality. The methodological limitations seen in all the included studies underline a dearth of stringent, high-quality research in this area.

The trials investigate many different manualised interventions, which all have parent mediation in common, even though they may differ in other ways. Within the field, only a few interventions, e.g., PRT and PACT, are investigated in more than one trial. Some of the interventions have shown promising results in the trials in which they were examined, but these results may be obscured when they are pooled with results from trials of other interventions. When choosing a PMI, clinicians may look at results from individual trials demonstrating positive results for this intervention. Future research should replicate previous trials to improve the evidence base for specific PMIs.

This review collected outcome results at the end of the intervention, which may have limited the ability to capture meaningful developmental change over time. Specifically, the developmental interventions and NDBIs are designed to have effects on child development that may accumulate over time. This has been demonstrated e.g. in a six year follow-up study of the PACT intervention (86). A future review may focus on capturing follow-up developmental changes in children participating in PMIs.

Since parent fidelity to the implementation may vary, future research could explore whether families where parents demonstrate high intervention implementation fidelity experience more beneficial outcomes for both parents and children. It is also possible that some children may show a reduction in autism characteristics that surpasses the threshold for minimal important difference.

This review and meta-analysis’ modest findings raise the question of which measures capture change through PMIs. A recent qualitative study of the effects of the PMI PACT contradicts our findings, as parents report experiences of substantial improvements in the child’s language and social communication development, resulting in relational improvements and positive cascading effects in the family and day-care facilities (87). Participants’ experience raises the question of whether the standardised scales are sensitive in capturing meaningful changes in autistic children and their parents. Some of the effects may be better captured in future trials by other measures, e.g., quality of life and family life functioning.

We recommend future research that incorporates an assessment of quality of life, adverse effects, parental sensitivity/synchronicity and child attachment outcomes. Importantly, our findings do not support abandoning PMIs. Rather, they underscore the need for more high-quality research to better understand their effects and to identify which children and families benefit most.

Finally, we acknowledge that some autistic individuals and autism communities object to terminology such as intervention, symptoms, and symptomatology, as well as deficit-focused descriptors more broadly. The terminology used in this review reflects that of its pre-registered protocol (8). Future research may better serve all communities by adopting growth-focused language, such as developmental progress, as primary outcome descriptors.

## Conclusion

PMIs, compared to the control condition, showed no significant benefits as measured on child autism characteristics, child adaptive functioning, child language, or parental stress. We recommend that future research use high-quality, predefined methodology in randomised clinical trials by designing the protocol in accordance with the Standard Protocol Items: Recommendations for Interventional Trials (SPIRIT) guidelines (88). When choosing outcomes, validated assessment tools used in previous research should be chosen. Future trials could, in addition to more commonly used outcomes, assess adverse effects, child and parental quality of life, family-life functioning, and child attachment.

## Supporting Information

Appendix S1 PRISMA checklist

Appendix S2 Search History

Appendix S3 Description of included studies

Appendix S4 List of excluded studies

Appendix S5 Forest plots and sub-group analyses of secondary outcomes

Appendix S6 Meta-analysis fixed effects

Appendix S7 Forest plots and subgroup analyses of exploratory outcomes

## Data availability

The data that support the findings of this review are available from the corresponding author on reasonable request.

## Author Contributions

Conceptualization: All authors. Data collection and extraction of data: CEC, SMTZ, CK, MV and MBL. Data analysis: HJ, EF and CEC. Supervision of data analysis: JCJ. Writing the manuscript: CEC and SMTZ. MBL, JCJ and CK made major contributions to the draft and revisions. All authors critically revised the manuscript and approved of the final version of the manuscript.

## Acknowledgements

The authors thank Jette Frost Jepsen, of the Medical Library, Aalborg University Hospital and Sarah Klingenberg from Copenhagen Trial Unit for their help conducting the systematic search.

## PRISMA 2020 Checklist

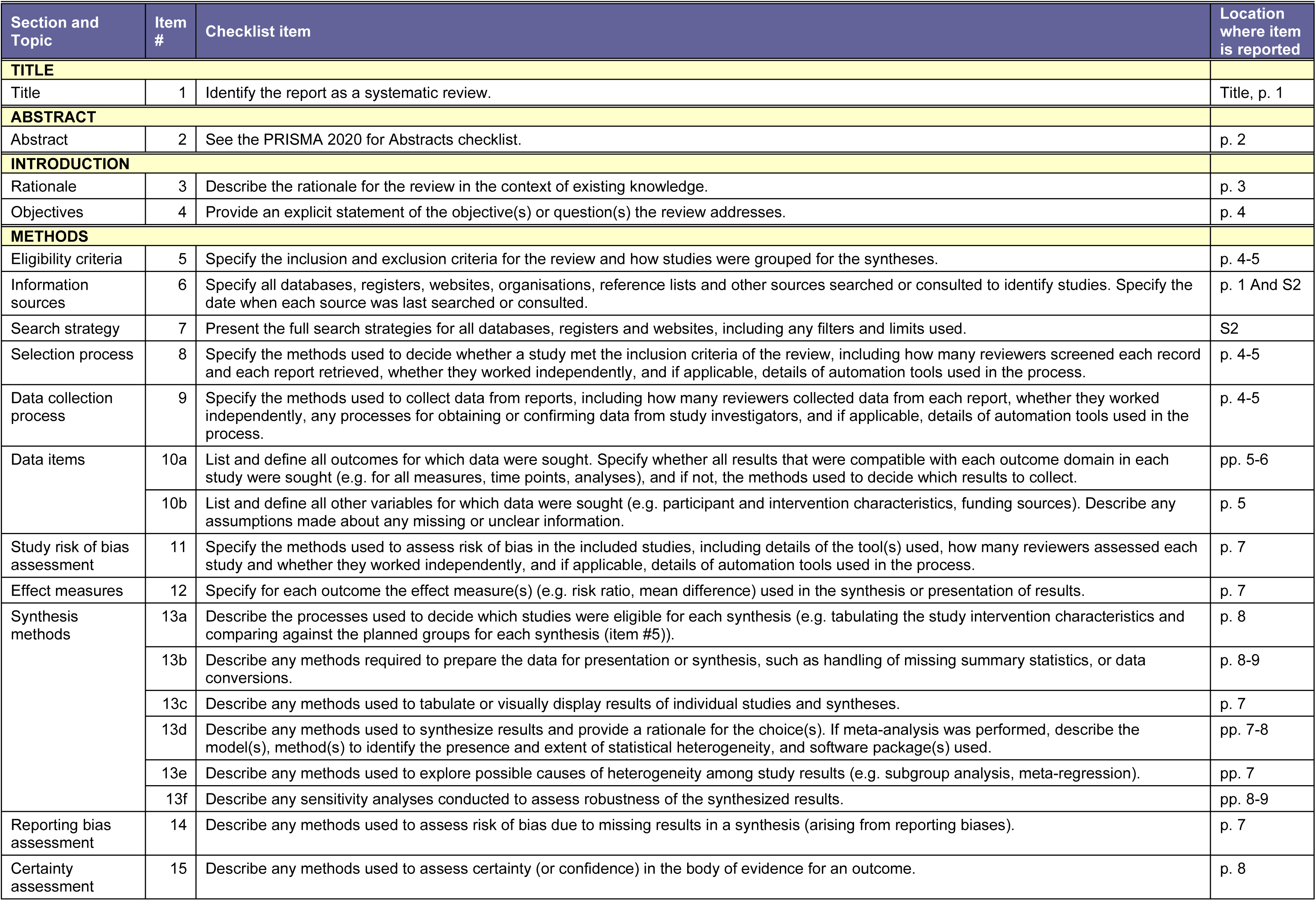
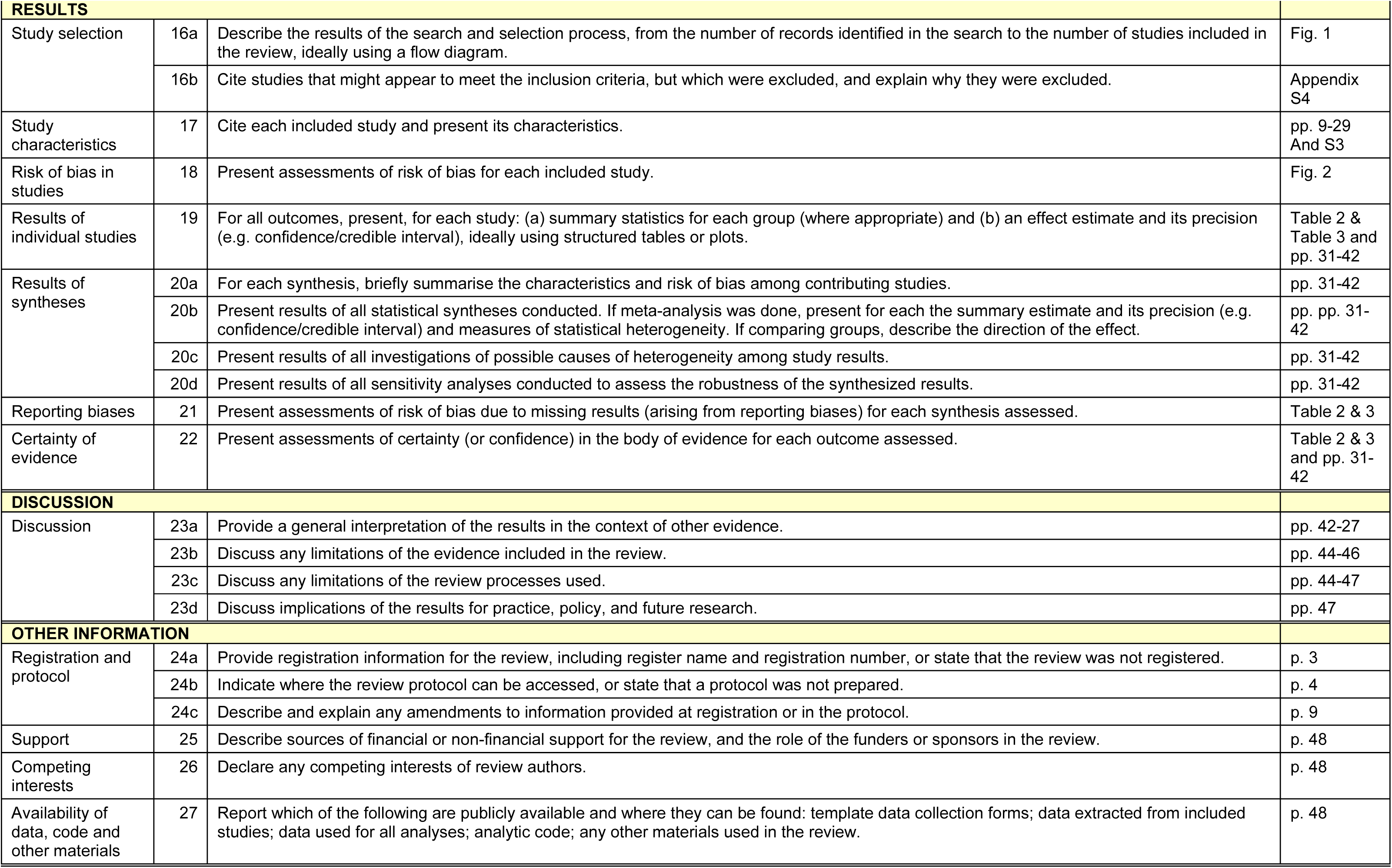
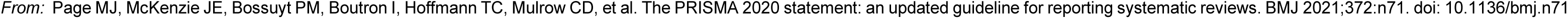

## S2 Search History

Search strategies for ‘Parent-mediated interventions versus usual care in children with autism spectrum disorders’ Databases and results

Databases and results
| Database | Platform | Results | Date |
| --- | --- | --- | --- |
| Medline | Ovid | 4333 | 14.08.2025 |
| Embase | Ovid | 7171 | 14.08.2025 |
| Cochrane Library | Wiley | 4753 | 14.08.2025 |
| Web of Science | Clarivate | 693 | 14.08.2025 |
| PsycINFO | APA | 1637 | 14.08.2025 |
| LILACS | Regional Portal of the Virtual Health Library (VHL) | 1164 | 14.08.2025 |
| All |  | 19751 |  |
| ÷ duplicates with EndNote |  | 12341 |  |

Ovid MEDLINE(R) Epub Ahead of Print and In-Process, In-Data-Review & Other Non-Indexed Citations and Daily August 13, 2025

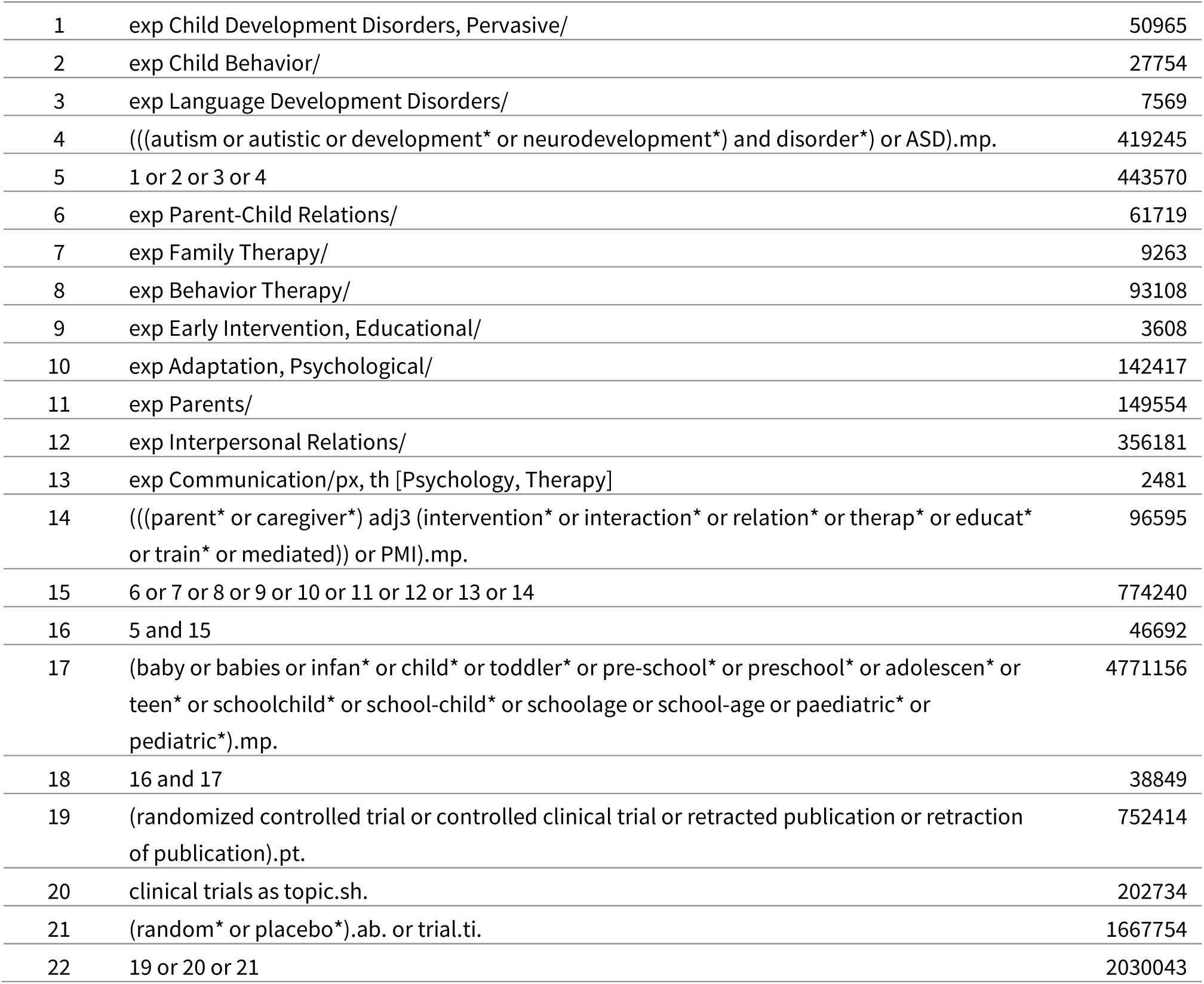
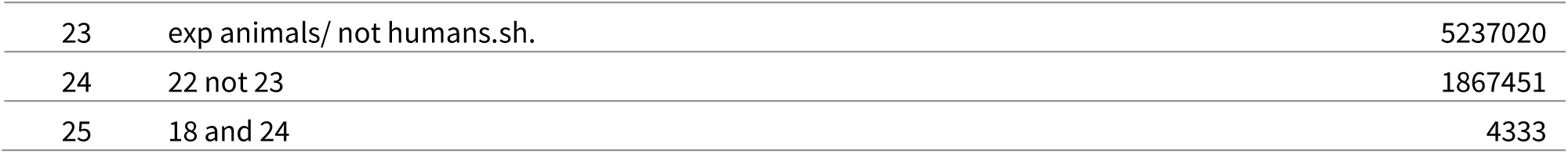

Embase <1974 to 2025 August 12>

|  |  |  |
| --- | --- | --- |
| 1 | exp *autism/ | 62800 |
| 2 | exp *child behavior/ | 23012 |
| 3 | exp *developmental language disorder/ | 1562 |
| 4 | ((autism or autistic or development* or neurodevelopment*) and disorder*) or ASD).mp. | 634495 |
| 5 | 1 or 2 or 3 or 4 | 668776 |
| 6 | exp *child parent relation/ | 44988 |
| 7 | exp *family therapy/ | 7159 |
| 8 | exp *behavior therapy/ | 30311 |
| 9 | exp *early childhood intervention/ | 1672 |
| 10 | exp *psychological adjustment/ | 3971 |
| 11 | exp *parent/ | 71700 |
| 12 | exp *human relation/ | 310784 |
| 13 | exp *interpersonal communication/ | 226378 |
| 14 | ((parent* or caregiver*) adj3 (intervention* or interaction* or relation* or therap* or educat* or train* or mediated)) or PMI).mp. | 139665 |
| 15 | 6 or 7 or 8 or 9 or 10 or 11 or 12 or 13 or 14 | 705261 |
| 16 | 5 and 15 | 38352 |
| 17 | (baby or babies or infan* or child* or toddler* or pre-school* or preschool* or adolescen* or teen* or schoolchild* or school-child* or schoolage or school-age or paediatric* or pediatric*).mp. | 4808230 |
| 18 | 16 and 17 | 32229 |
| 19 | *Randomized controlled trial/ or *Controlled clinical study/ or *randomization/ or *intermethod comparison/ or *double blind procedure/ or *human experiment/ or *retracted article/ | 44433 |
| 20 | (random\$ or placebo or parallel group\$1 or crossover or cross over or assigned or allocated or volunteer or volunteers).ti,ab. | 2716303 |
| 21 | (compare or compared or comparison or trial).ti. | 1040028 |
| 22 | ((evaluated or evaluate or evaluating or assessed or assess) and (compare or compared or comparing or comparison)).ab. | 2952327 |
| 23 | (open adj label).ti,ab. | 116471 |
| 24 | ((double or single or doubly or singly) adj (blind or blinded or blindly)).ti,ab. | 284141 |
| 25 | ((assign\$ or match or matched or allocation) adj5 (alternate or group\$1 or intervention\$1 or patient\$1 or subject\$1 or participant\$1)).ti,ab. | 436695 |
| 26 | (controlled adj7 (study or design or trial)).ti,ab. | 476076 |
| 27 | (erratum or tombstone).pt. or yes.ne. | 319993 |
| 28 | or/19-27 | 6136892 |
| 29 | (random\$ adj sampl\$ adj7 ('cross section\$' or questionnaire\$ or survey\$ or database\$1)).ti,ab. not (comparative study/ or controlled study/ or randomi?ed controlled.ti,ab. or randomly assigned.ti,ab.) | 10045 |
| 30 | *Cross-sectional study/ not (*randomized controlled trial/ or *controlled clinical study/ or *controlled study/ or randomi?ed controlled.ti,ab. or control group\$1.ti,ab.) | 14352 |
| 31 | ((((case adj control\$) and random\$) not randomi?ed controlled).ti,ab. | 22549 |
| 32 | (Systematic review not (trial or study)).ti. | 290441 |
| 33 | (nonrandom\$ not random\$).ti,ab. | 19590 |
| 34 | 'Random field\$.ti,ab. | 3087 |
| 35 | (random cluster adj3 sampl\$).ti,ab. | 1660 |
| 36 | (review.ab. and review.pt.) not trial.ti. | 1209375 |
| 37 | 'we searched'.ab. and (review.ti. or review.pt.) | 53571 |
| 38 | 'update review'.ab. | 143 |
| 39 | (databases adj4 searched).ab. | 69311 |
| 40 | (rat or rats or mouse or mice or swine or porcine or murine or sheep or lambs or pigs or piglets or rabbit or rabbits or cat or cats or dog or dogs or cattle or bovine or monkey or monkeys or trout or marmoset\$1).ti. and animal experiment/ | 1258378 |
| 41 | Animal experiment/ not (human experiment/ or human/) | 2646788 |
| 42 | or/29-41 | 4196966 |
| 43 | 28 not 42 | 5451939 |
| 44 | 18 and 43 | 7171 |

Cochrane Central Register of Controlled Trials Issue 7 of 12, July 2025

|  |  |  |
| --- | --- | --- |
| #1 | MeSH descriptor: [Child Development Disorders, Pervasive] explode all trees | 2900 |
| #2 | MeSH descriptor: [Child Behavior] explode all trees | 2898 |
| #3 | MeSH descriptor: [Language Development Disorders] explode all trees | 288 |
| #4 | ((((autism or autistic or development* or neurodevelopment*) and disorder*) or ASD):ti,ab,kw | 22678 |
| #5 | {OR #1-#4} | 25247 |
| #6 | MeSH descriptor: [Parent-Child Relations] explode all trees | 2705 |
| #7 | MeSH descriptor: [Family Therapy] explode all trees | 1184 |
| #8 | MeSH descriptor: [Behavior Therapy] explode all trees | 25045 |
| #9 | MeSH descriptor: [Early Intervention, Educational] explode all trees | 704 |
| #10 | MeSH descriptor: [Adaptation, Psychological] explode all trees | 6918 |
| #11 | MeSH descriptor: [Parents] explode all trees | 8803 |
| #12 | MeSH descriptor: [Interpersonal Relations] explode all trees | 8846 |
| #13 | MeSH descriptor: [Communication] explode all trees | 13330 |
| #14 | ((((parent* or caregiver*) near/3 (intervention* or interaction* or relation* or therap* or educat* or train* or mediated)) or PMI):ti,ab,kw | 17596 |
| #15 | {OR #6-#14} | 68233 |
| #16 | #5 AND #15 | 5240 |
| #17 | (baby or babies or infan* or child* or toddler* or pre-school* or preschool* or adolescen* or teen* or schoolchild* or school-child* or schoolage or school-age or paediatric* or pediatric*):ti,ab,kw | 367367 |
| #18 | #16 AND #17 in Trials | 4753 |

Web of Science: Science Citation Index Expanded (1900-present) and Conference Proceedings Citation Index – Science (1990-present)

|  |  |  |
| --- | --- | --- |
| #1 | ((autism or autistic or development* or neurodevelopment*) and disorder*) or ASD) (Topic) | 324525 |
| #2 | ((parent* or caregiver*) near/3 (intervention* or interaction* or relation* or therap* or educat* or train* or mediated)) or PMI) (Topic) | 62080 |
| #3 | (baby or babies or infan* or child* or toddler* or pre-school* or preschool* or adolescen* or teen* or schoolchild* or school-child* or schoolage or school-age or paediatric* or pediatric*) (Topic) | 2640254 |
| #4 | #1 AND #2 AND #3 | 3445 |
| #5 | (random* or blind* or placebo* or meta-analys* or trial*) (Title) OR (random* or blind* or placebo* or meta-analys*) (Topic) | 3087460 |
| #6 | #4 AND #5 | 693 |

APA PsycInfo, APA PsycArticles

|  |  |  |
| --- | --- | --- |
| 8 | 5 AND 6 AND Publication Type: Peer Reviewed Journal | 1637 |
| 7 | 5 AND 6 | 1884 |
| 6 | Index Terms: {Randomized Controlled Trials} OR {Clinical Trials} OR Title: trial OR Abstract: random* OR Abstract: placebo* | 289915 |
| 5 | 3 AND 4 | 30523 |
| 4 | Title: baby OR Title: babies OR Title: infan* OR Title: child* OR Title: toddler* OR Title: "pre-school*" OR Title: preschool* OR Title: adolescen* OR Title: teen* OR Title: schoolchild* OR Title: "school-child*" OR Title: schoolage OR Title: "school-age" OR Title: paediatric* OR Title: pediatric* OR Abstract: baby OR Abstract: babies OR Abstract: infan* OR Abstract: child* OR Abstract: toddler* OR Abstract: "pre-school*" OR Abstract: preschool* OR Abstract: adolescen* OR Abstract: teen* OR Abstract: schoolchild* OR Abstract: "school-child*" OR Abstract: schoolage OR Abstract: "school-age" OR Abstract: paediatric* OR Abstract: pediatric* OR Keywords: baby OR Keywords: babies OR Keywords: infan* OR Keywords: child* OR Keywords: toddler* OR Keywords: "pre-school*" OR Keywords: preschool* OR Keywords: adolescen* OR Keywords: teen* OR Keywords: schoolchild* OR Keywords: "school-child*" OR Keywords: schoolage OR Keywords: "school-age" OR Keywords: paediatric* OR Keywords: pediatric* | 1129017 |
| 3 | 1 AND 2 | 36991 |
| 2 | Index Terms: {Parent Child Relations} OR {Father Child Relations} OR {Mother Child Relations} OR {Parental Attitudes} OR Index Terms: {Family Therapy} OR {Conjoint Therapy} OR {Strategic Family Therapy} OR {Structural Family Therapy} OR Index Terms: {Early Intervention} OR {Coping Behavior} OR Index Terms: {Parents} OR {Adoptive Parents} OR {Expectant Parents} OR {Fathers} OR {Foster Parents} OR {Homosexual Parents} OR {Mothers} OR {Parental Characteristics} OR {Single Parents} OR {Stepparents} OR {Surrogate Parents (Humans)} OR Index Terms: {Interpersonal Relationships} OR {Caregivers} OR {Close Relationships} OR {Couples} OR {Family Relations} OR {Friendship} OR {Kinship} OR {Marital Relations} OR {Mentor} OR {Partners} OR {Peers} OR {Relationship Quality} OR {Relationship Termination} OR {Role Models} OR {Significant Others} OR Index Terms: {Interpersonal Communication} OR {Active Listening} OR {Apology} OR {Arguments} OR {Body Language} OR {Conversation} OR {Cross Cultural Communication} OR {Double Bind Interaction} OR {Eye | 523016 |
|  | Contact} OR {Gossip} OR {Group Discussion} OR {Intersubjectivity} OR {Interviewing} OR<br>{Interviews} OR {Job Applicant Interviews} OR {Listening (Interpersonal)} OR {Negotiation} OR<br>{Parent Child Communication} OR {Psychological Contracts} OR {Secrecy} OR {Self-Disclosure}<br>OR ((Title: parent* OR Title: caregiver*) NEAR/3 (Title: intervention* OR Title: interaction* OR<br>Title: relation* OR Title: therap* OR Title: educat* OR Title: train* OR Title: mediated)) OR<br>Title: PMI OR ((Abstract: parent* OR Abstract: caregiver*) NEAR/3 (Abstract: intervention* OR<br>Abstract: interaction* OR Abstract: relation* OR Abstract: therap* OR Abstract: educat* OR<br>Abstract: train* OR Abstract: mediated)) OR Abstract: PMI OR ((Keywords: parent* OR<br>Keywords: caregiver*) NEAR/3 (Keywords: intervention* OR Keywords: interaction* OR<br>Keywords: relation* OR Keywords: therap* OR Keywords: educat* OR Keywords: train* OR<br>Keywords: mediated)) OR Keywords: PMI |  |
| 1 | Index Terms: {Neurodevelopmental Disorders} OR {Attention Deficit Disorder} OR {Autism<br>Spectrum Disorders} OR {Developmental Disabilities} OR {Disruptive Behavior Disorders} OR<br>{Emotional and Behavioral Disorders} OR {Intellectual Development Disorder} OR {Learning<br>Disorders} OR Index Terms: {Child Behavior} OR Index Terms: {Language Development} OR<br>{Language Delay} OR {Reading Development} OR {Speech Development} OR {Delayed Speech}<br>OR ((Title: autism OR Title: autistic OR Title: development* OR Title: neurodevelopment*) AND<br>Title: disorder*) OR Title: ASD OR ((Abstract: autism OR Abstract: autistic OR Abstract:<br>development* OR Abstract: neurodevelopment*) AND Abstract: disorder*) OR Abstract: ASD<br>OR ((Keywords: autism OR Keywords: autistic OR Keywords: development* OR Keywords:<br>neurodevelopment*) AND Keywords: disorder*) OR Keywords: ASD | 276943 |

Latin American and Caribbean Health Sciences Literature (LILACS)

|  |  |  |
| --- | --- | --- |
| 1 | ((((autism OR autistic OR development* OR neurodevelopment*) AND disorder*) OR asd) )<br>AND (((((parent* OR caregiver*) AND (intervention* OR interaction* OR relation* OR therap*<br>OR educat* OR train* OR mediated)) OR pmi)) AND ((baby OR babies OR infan* OR child* OR<br>toddler* OR pre-school* OR preschool* OR adolescen* OR teen* OR schoolchild* OR school-<br>child* OR schoolage OR school-age OR paediatric* OR pediatric*)) AND ( db:("LILACS")) | 1164 |

## S3 Description of included studies

### Control conditions

Eleven studies reported a waiting list control condition (1–9), twenty studies described a “care as usual” control condition (10–28), and one (29) reported a control condition of “no treatment”. A detailed description of all studies is included in a descriptive table (Table S3).

### Intervention content

The content and theoretical basis of the interventions across all studies varied. The interventions in eight studies were NDBIs (1, 2, 4, 12, 19, 22, 23, 26). 14 studies examined developmental interventions (5, 7, 10, 11, 13–16, 20, 24, 25, 27, 29–32). Eight studies were based on behavioral interventions (6, 8, 9, 17, 18, 21, 29, 33). Goharbakhsh et al. (2023) examined the unclassified intervention “Teaching Children to Mind Read”.

### Duration

The duration of the intervention in the included studies varied greatly. Interventions lasted seven weeks (7), eight weeks (3, 4, 28, 32, 34), 9 weeks (8), 10 weeks (2), 12 weeks (1, 5, 19, 20, 23, 27, 30, 33), 14 weeks (6, 29), 16 weeks (9, 17), 20 weeks (21), 22 weeks (18), 24 weeks (10, 11, 15), 28 weeks with a range of 12 to 52 weeks (24), 32 weeks (25), 12 months (13, 14, 22, 31), and 2 years (12). One study (26) provided either 12 or 20 weeks of intervention depending on the participant’s rated level of improvement at week 12.

### Location

The trials were conducted in eleven countries: 12 in the US (2, 4, 6, 7, 9, 13, 17, 20, 23–25, 29–31); four in the UK (3, 14, 16, 22); three in Australia (8, 10, 34); three in the Netherlands (12, 26, 27); two in Brazil (18, 19); two in Canada (1, 21); two in Iran (28, 33); one was conducted in both India and Pakistan (11); one in India (15); one in China (32); and one was conducted in Thailand (5).

## S4 List of Excluded studies – reason for exclusion

A total of 25 articles were excluded for the reason of wrong intervention e.g., 80 percent or more of the intervention was not implemented by parents (1–27). Twenty articles were excluded as their comparison groups did not meet criteria for inclusion (28–47). Sixteen articles were excluded as they did not have a correct study design, e.g., quasi-randomisation (48–63) Finally, in five studies participants did not meet criteria for inclusion in this review (64–68).

## S5 Forest plots of secondary outcomes

**Fig. 5.**
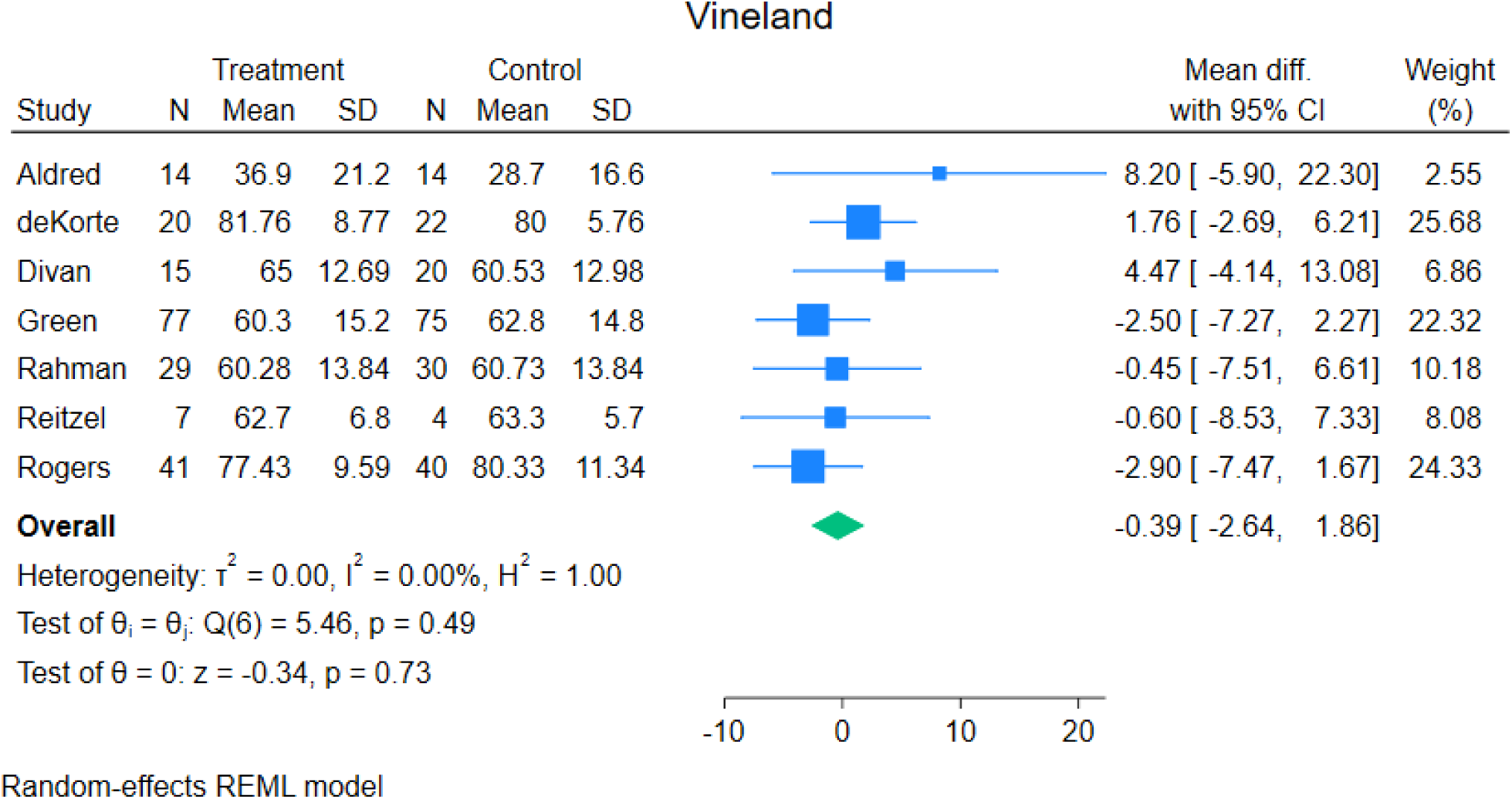
Forest plot of child adaptive functioning as measured by Vineland

**Fig. 6.**
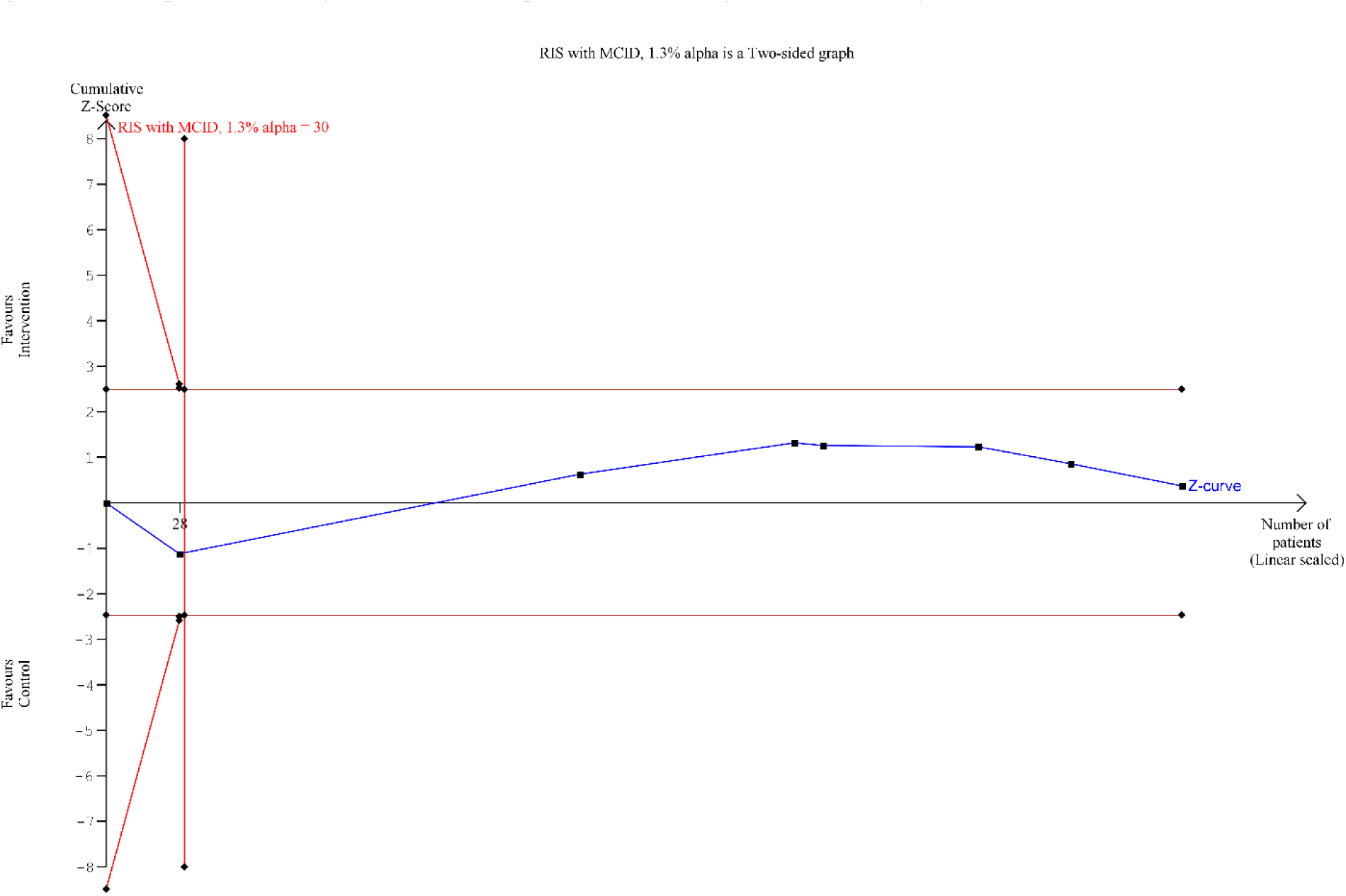
Trial Sequential Analysis of child adaptive functioning as measured by Vineland

**Fig. 7.**
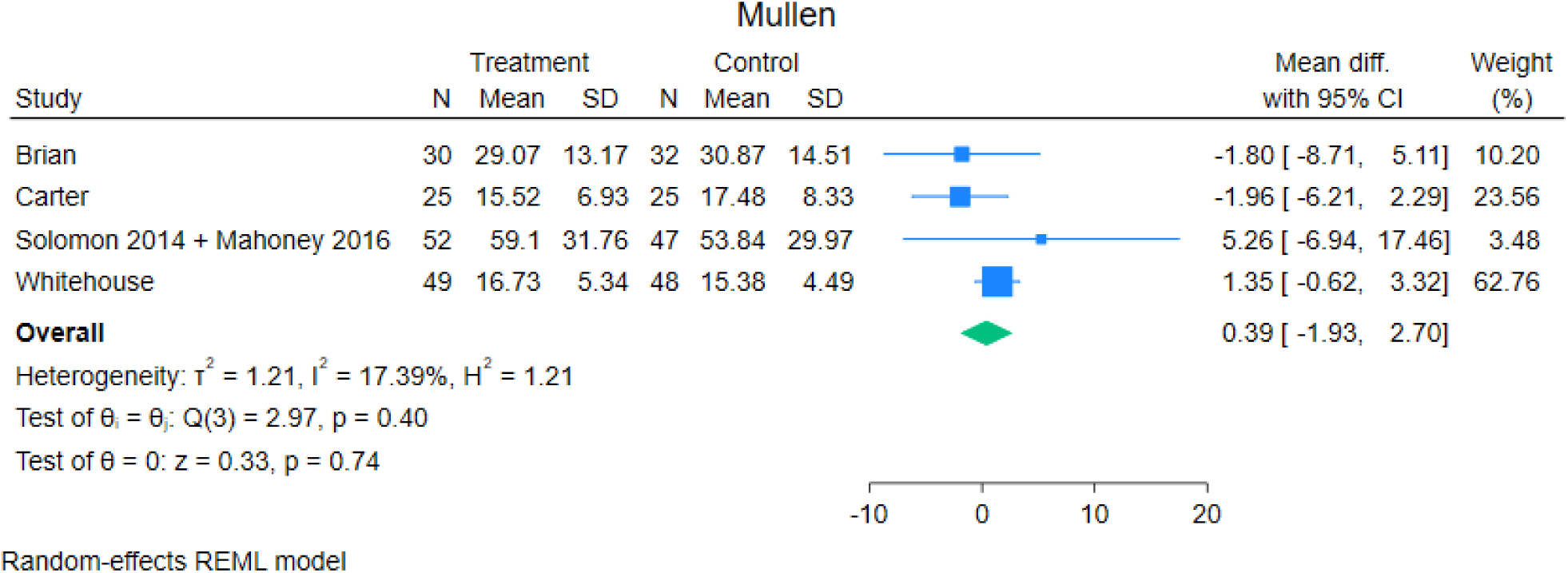
Forest plot of child language as measured by Mullen Scales of Early Learning Receptive Language

**Fig. 8.**
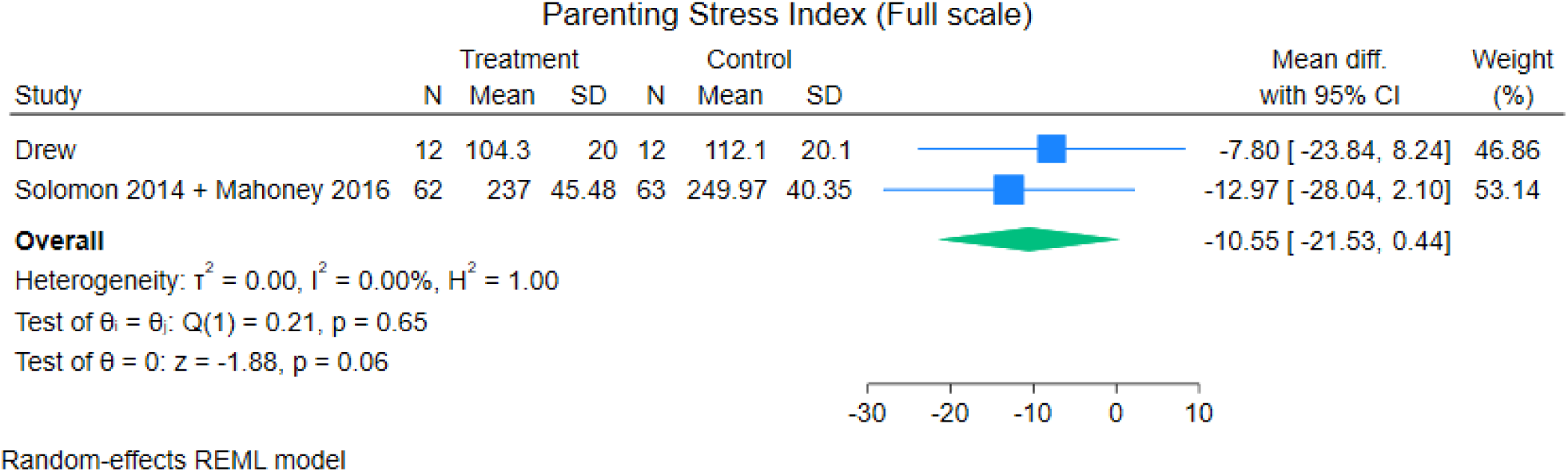
Forest plot of parental stress as measured by the full-scale Parenting Stress Index

**Fig. 9.**
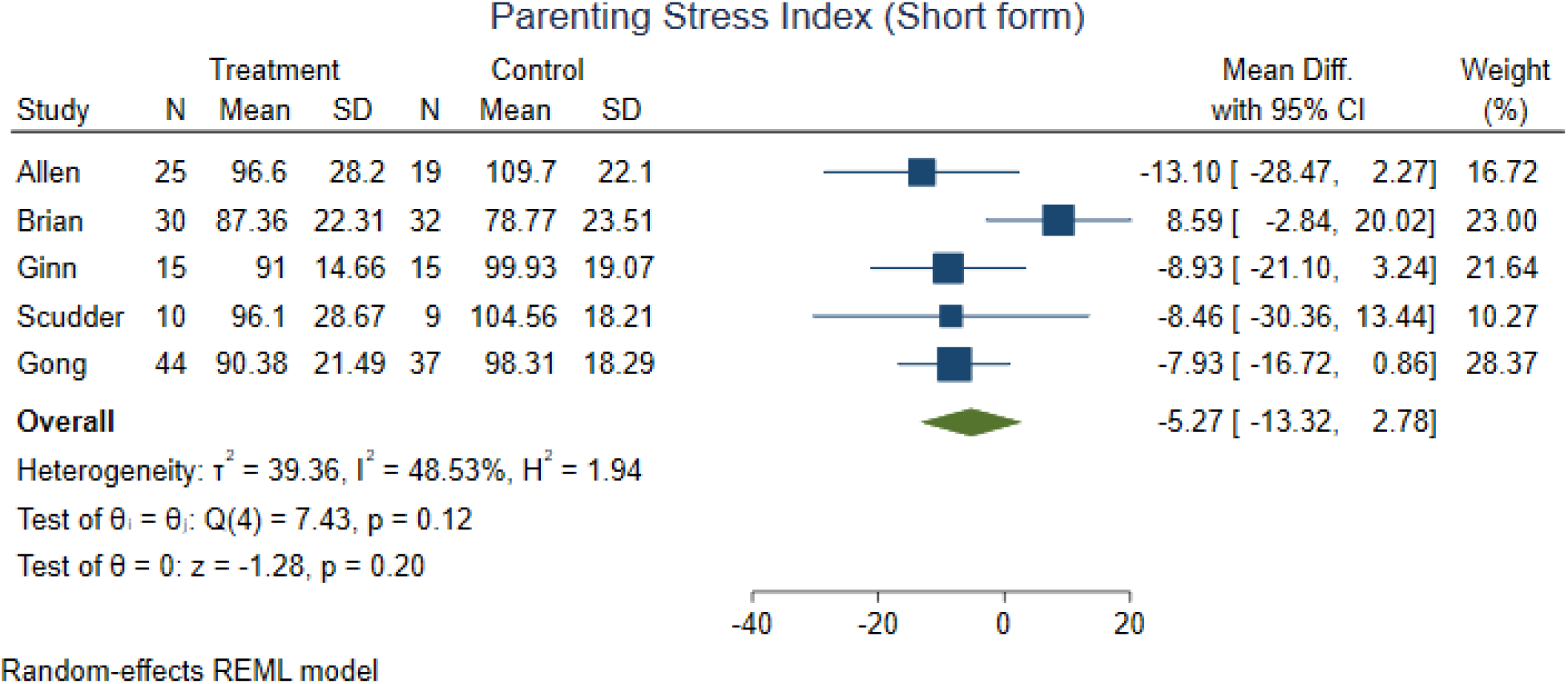
Forest plot of parental stress as measured by the Parenting Stress Index Short Form

## S7 Meta-analysis fixed effects

**Forest plot Autism Characteristics measured by ADOS total score**

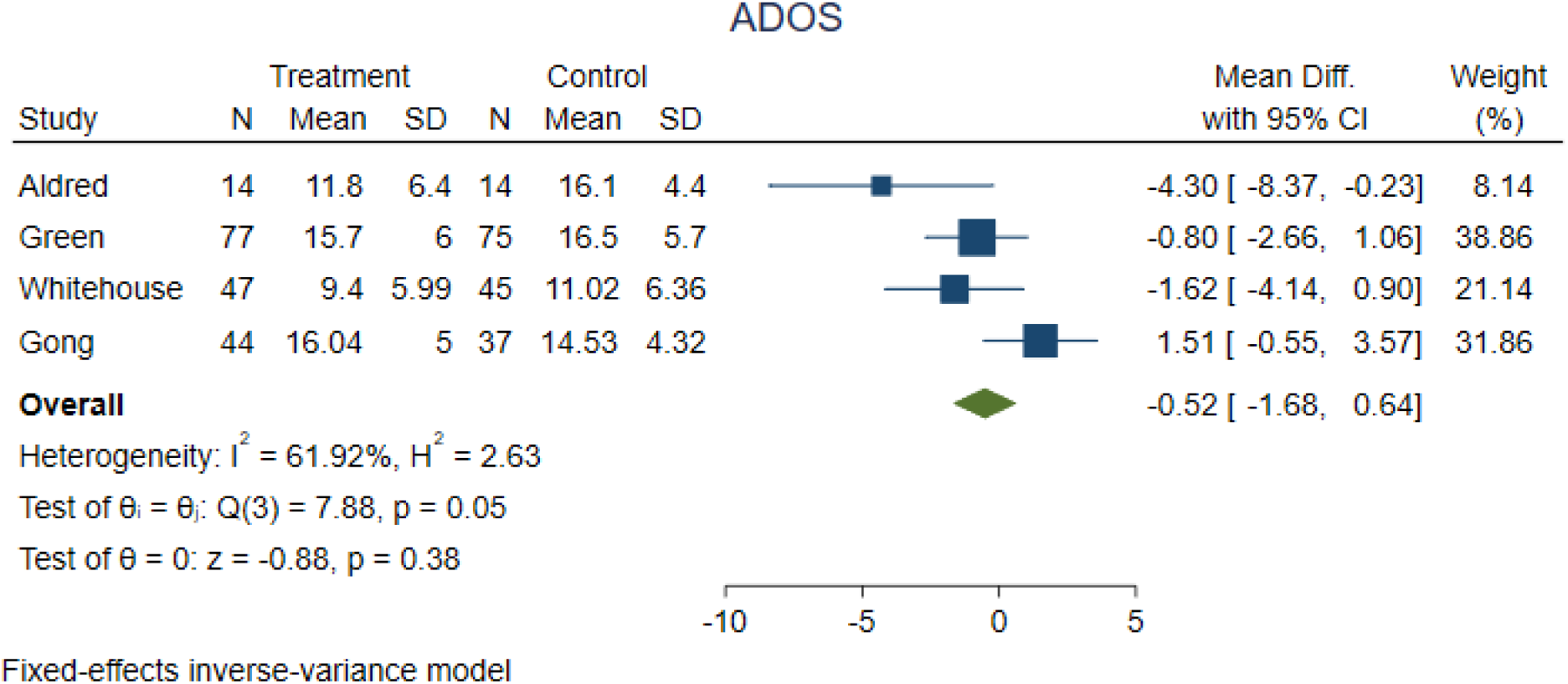

**Forest plot Child Adaptive Functioning as measured by Vineland**

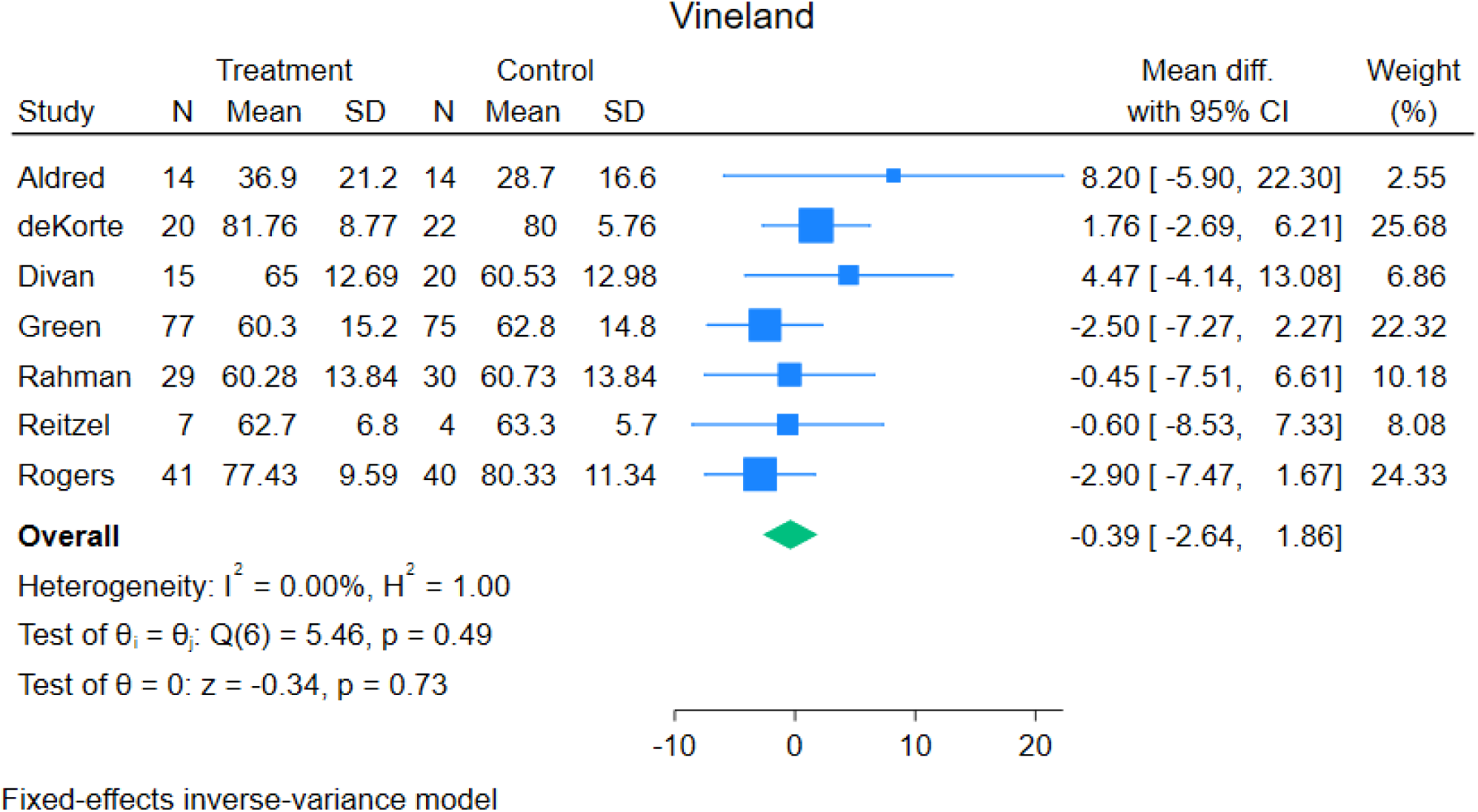

**Forest plot Child Language as measured by Mullen Scales of Early Learning**

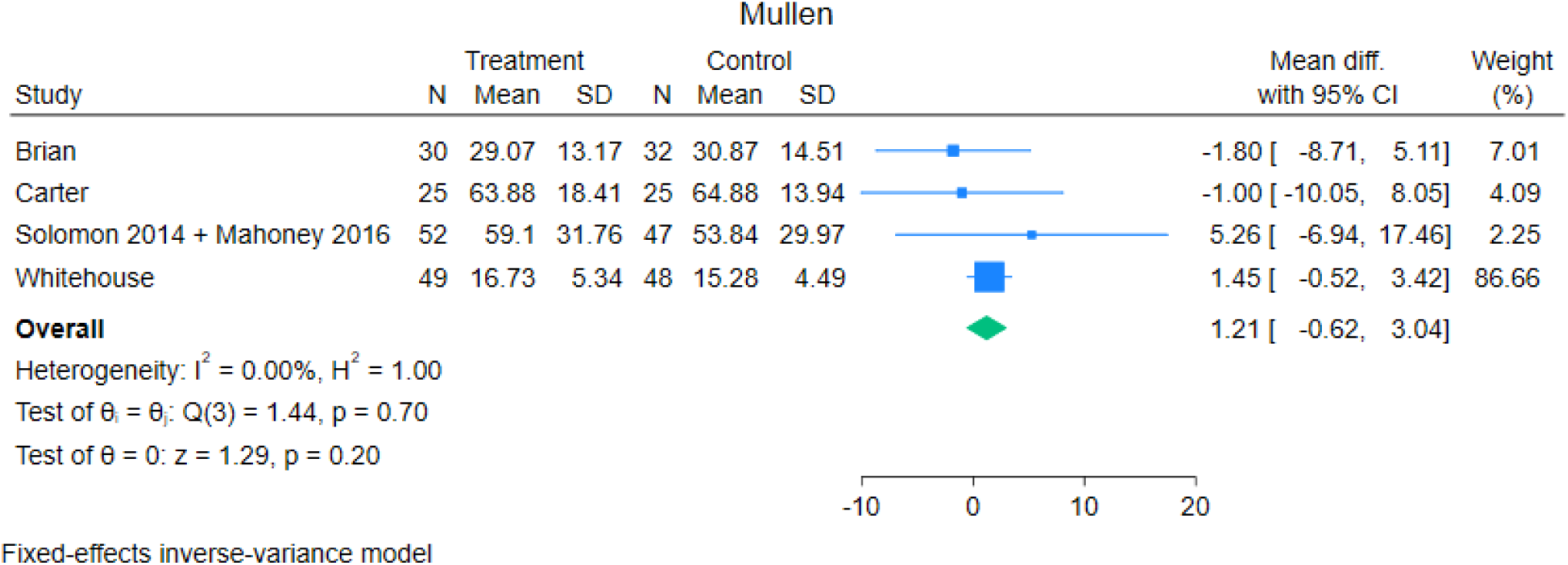

**Forest Plot, Parental Stress as measured by Parenting Stress Index full-scale**

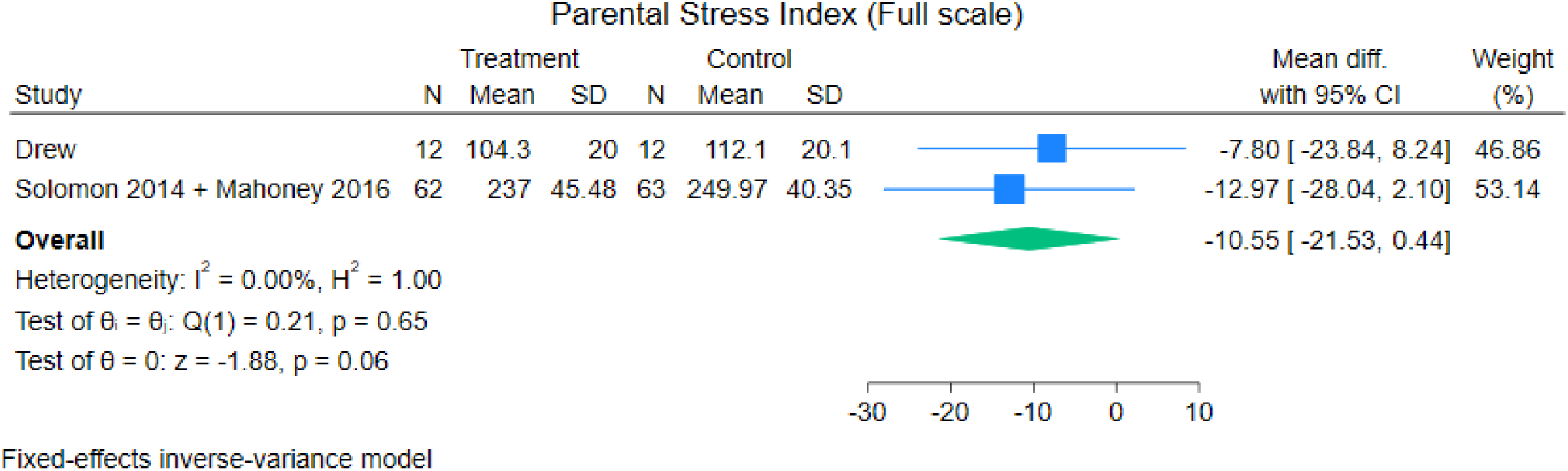

**Forest Plot, Parental Stress as measured by Parenting Stress Index short form**

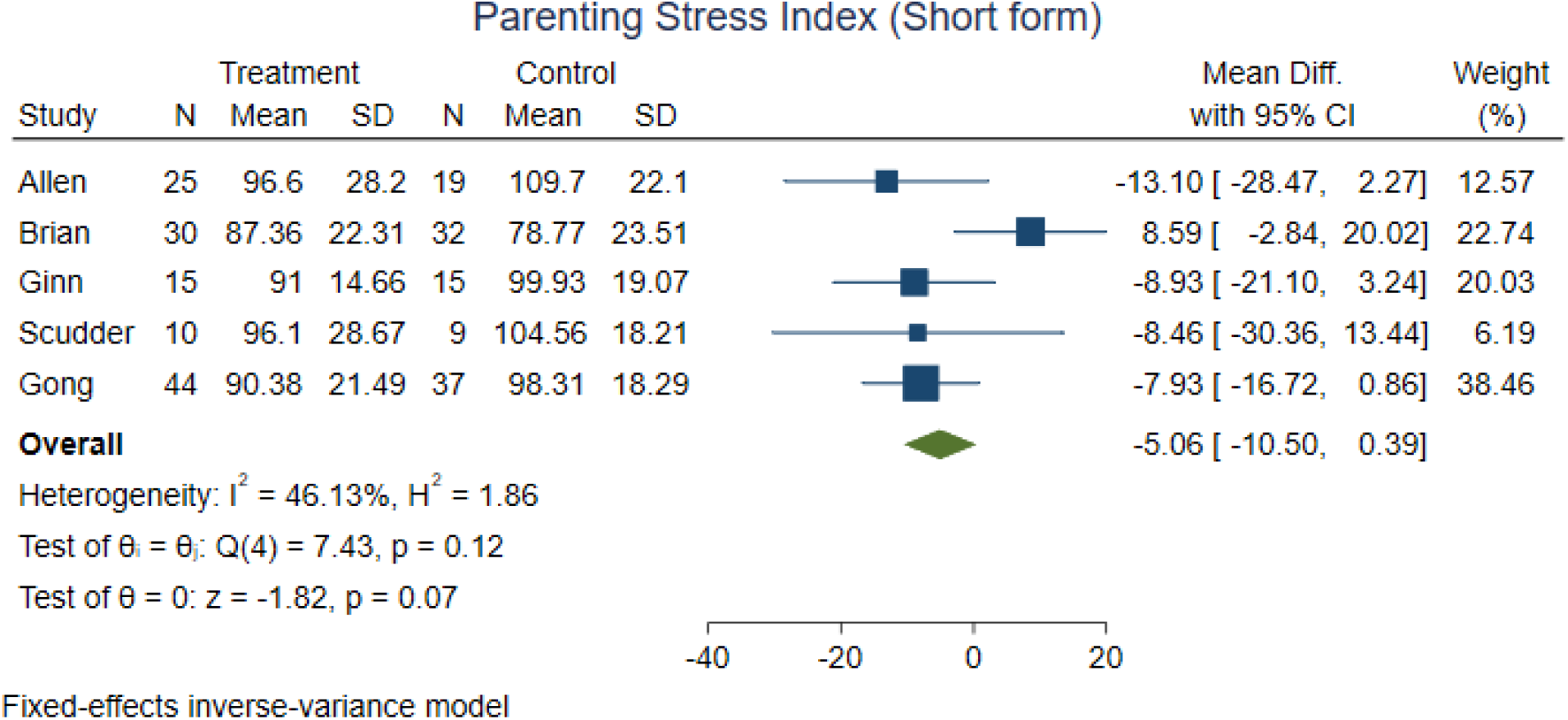

**Forest plot, Child Behaviour Problems**

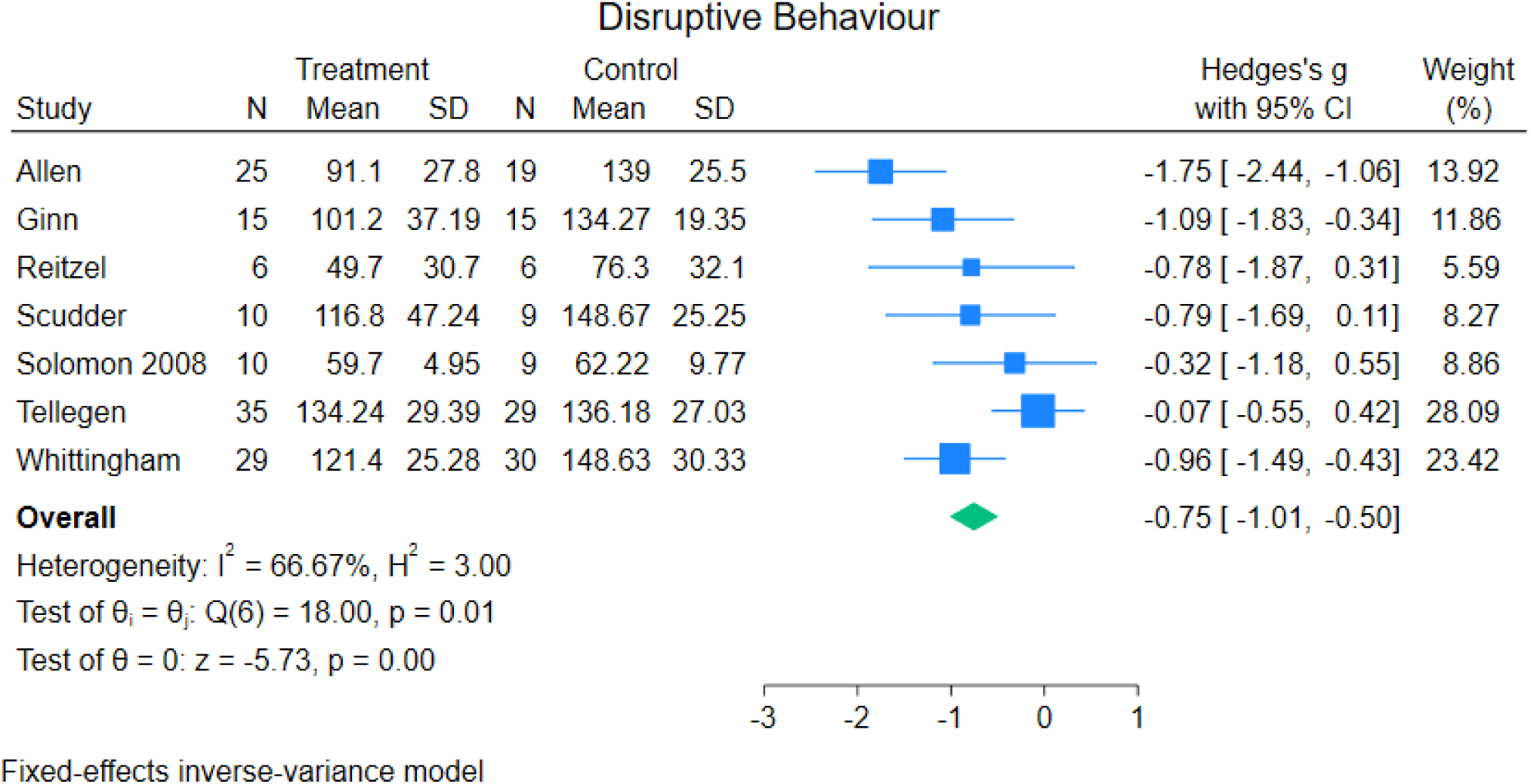

**Forest plot, Joint Attention**

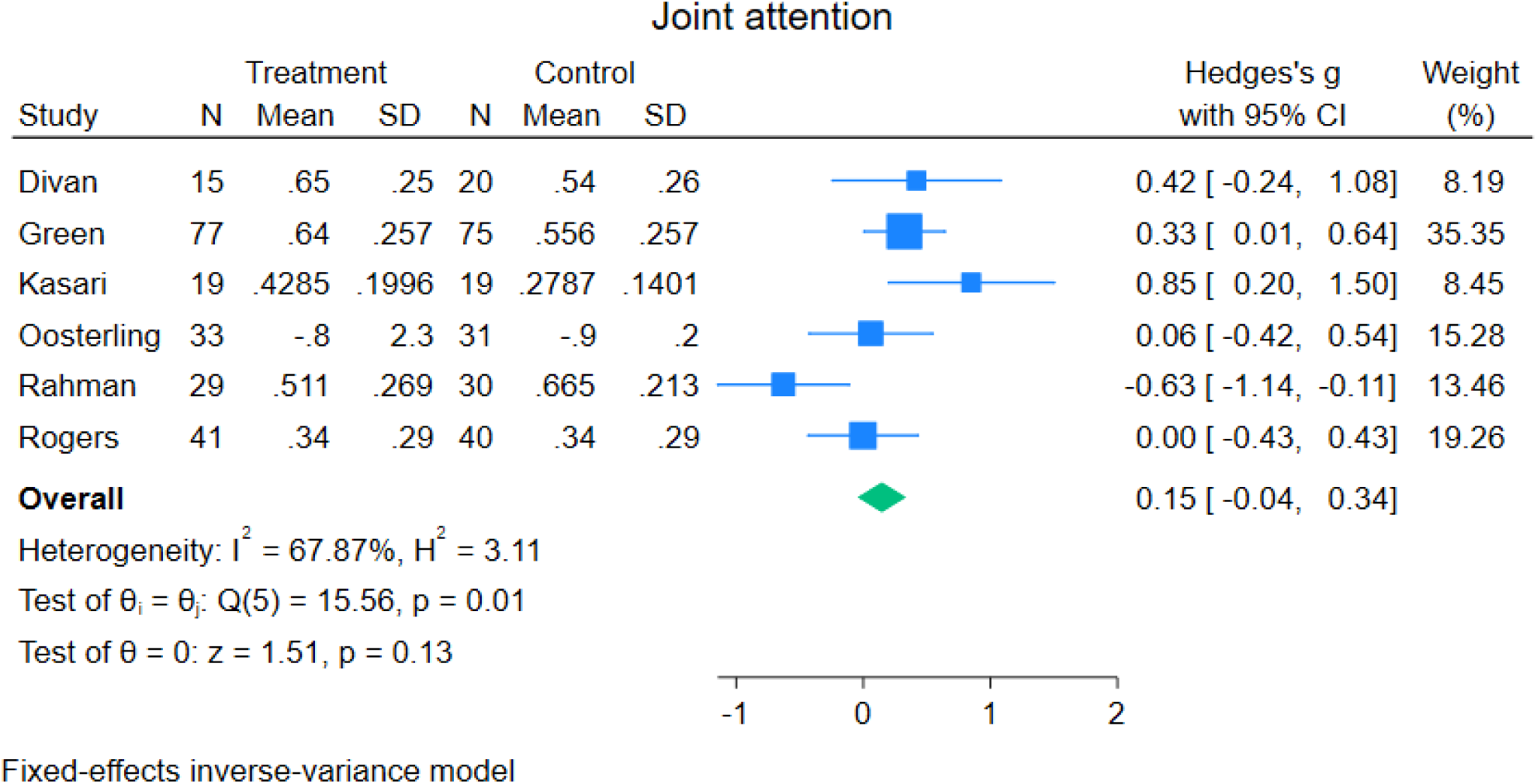

**Forest plot, Parent sensitivity/synchronicity**

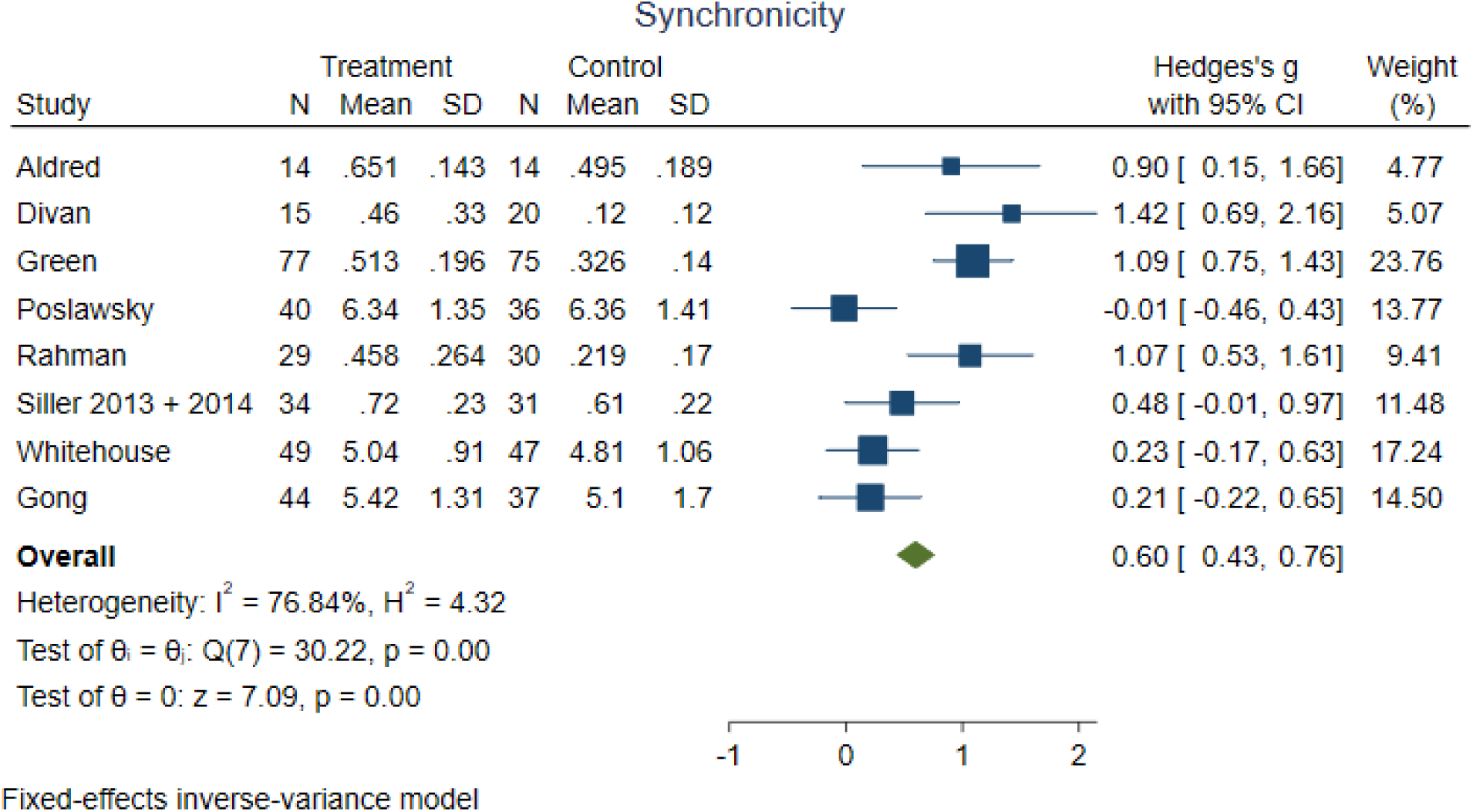

**Forest plot, Parent Fidelity**

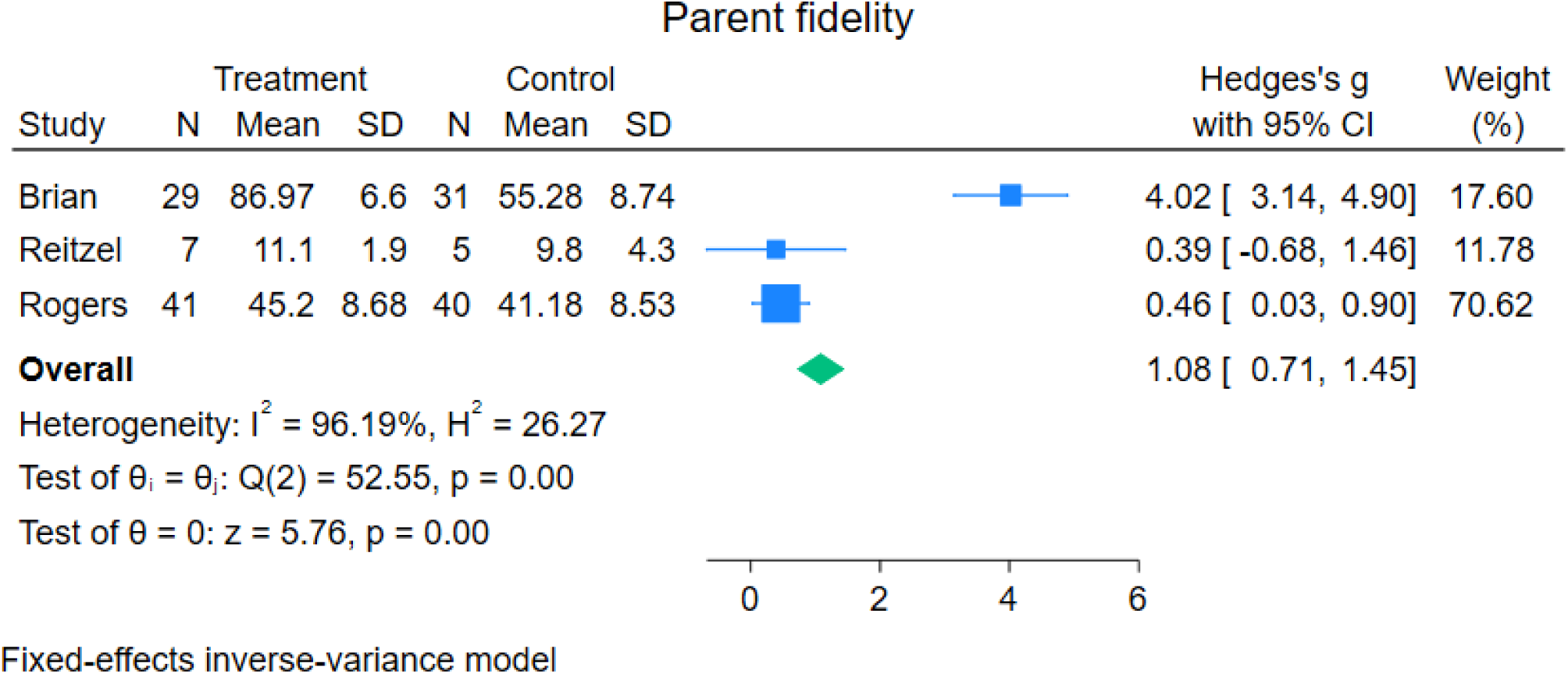

**Forest plot, Autism Characteristics (Not ADOS total)**

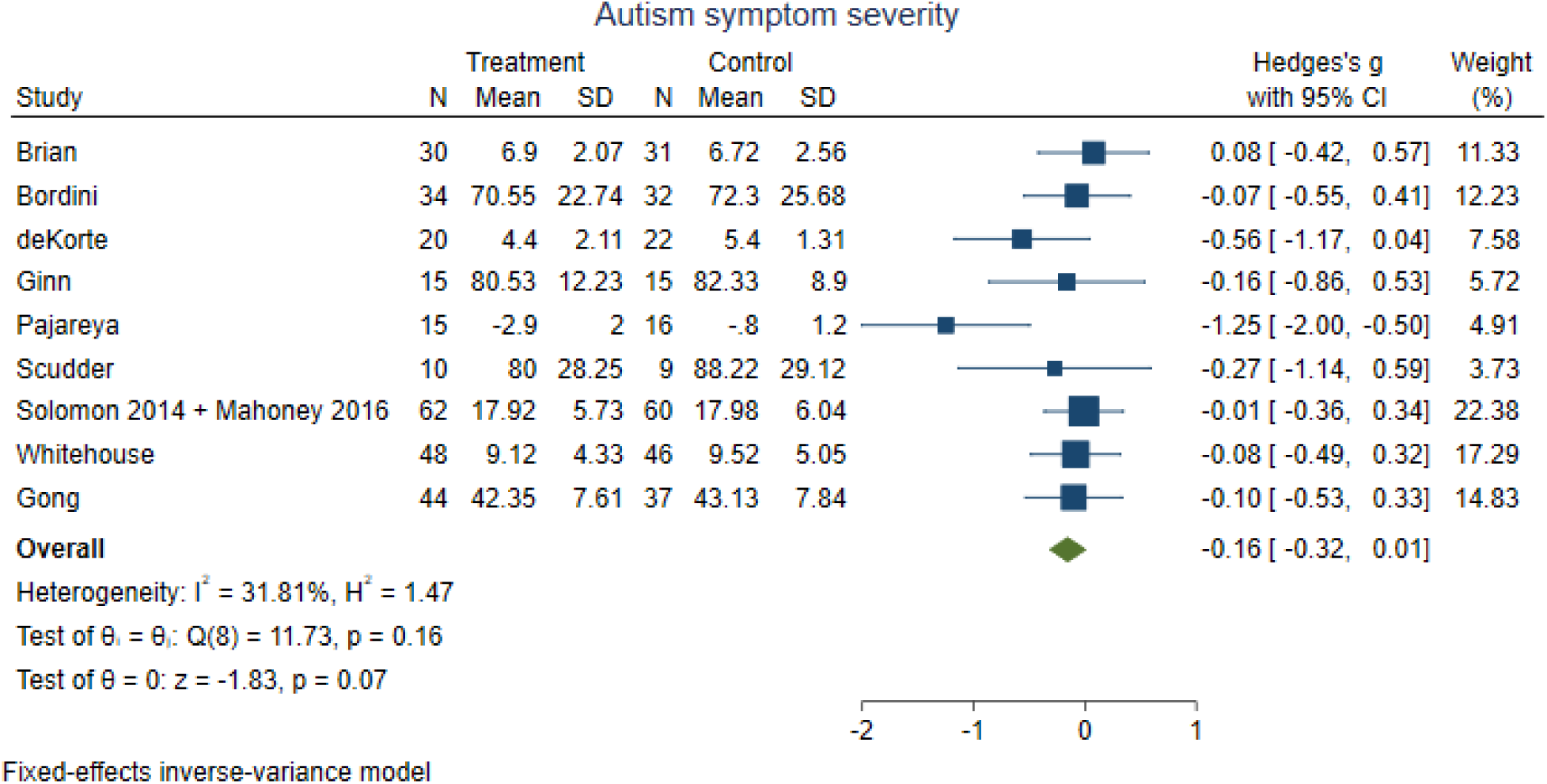

**Forest plot, Child Social Communication difficulties**

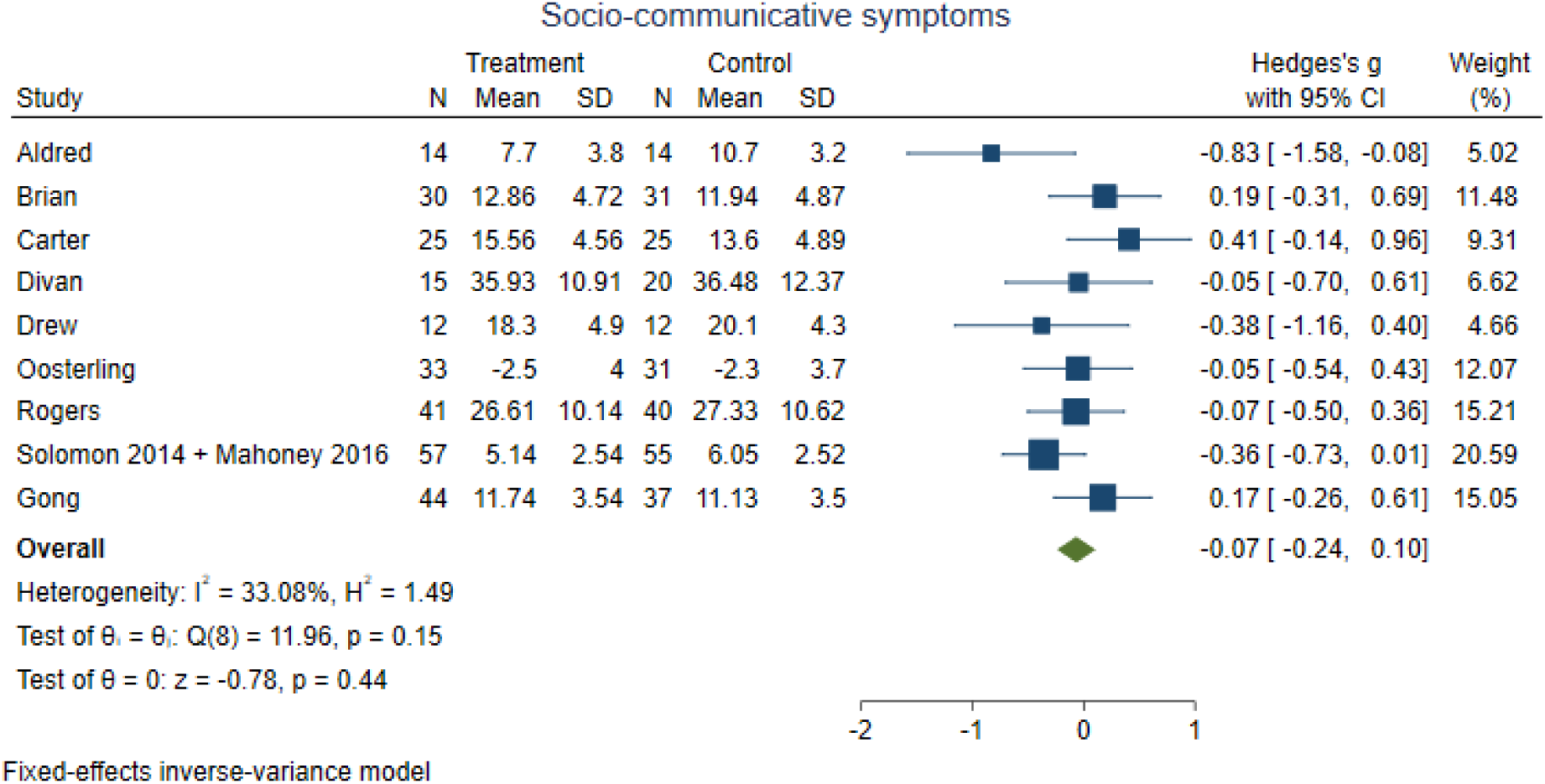

**Forest plot, Child Repetitive Behaviour**

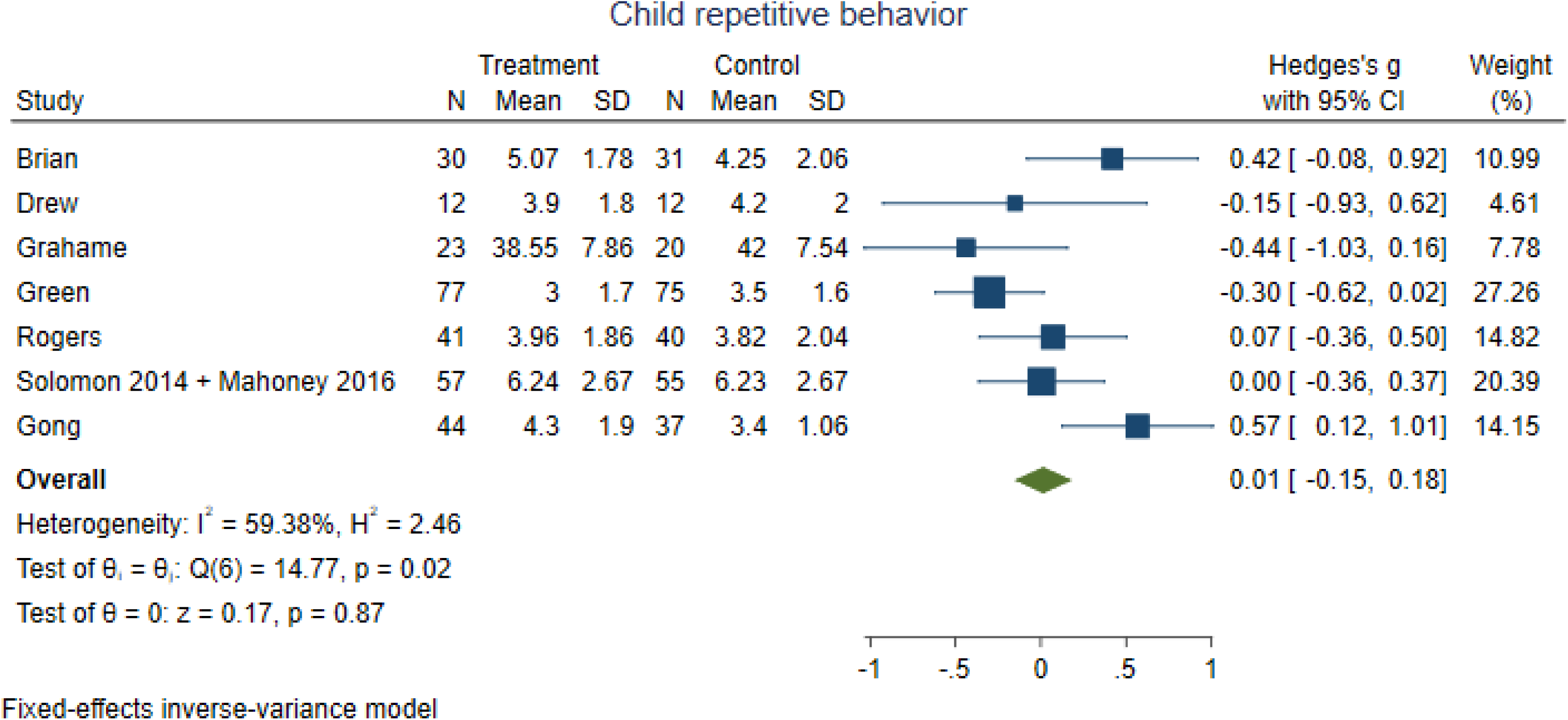

## S7 Forest plots and sub-group analyses of exploratory outcomes

### Child Behaviour Problems

Two trials assessed effects of Stepping Stones Triple-P versus usual care (1) and waiting list (2), respectively. Ginn et al. (2017) examined effects of Child-Directed Training (CDIT) versus waiting list, Reitzel et al. (2013) assessed effects of Functional Behavior Skills Training versus usual care, and Solomon et al. (2008) assessed effects of Parent-Child Interaction Therapy versus waiting list.

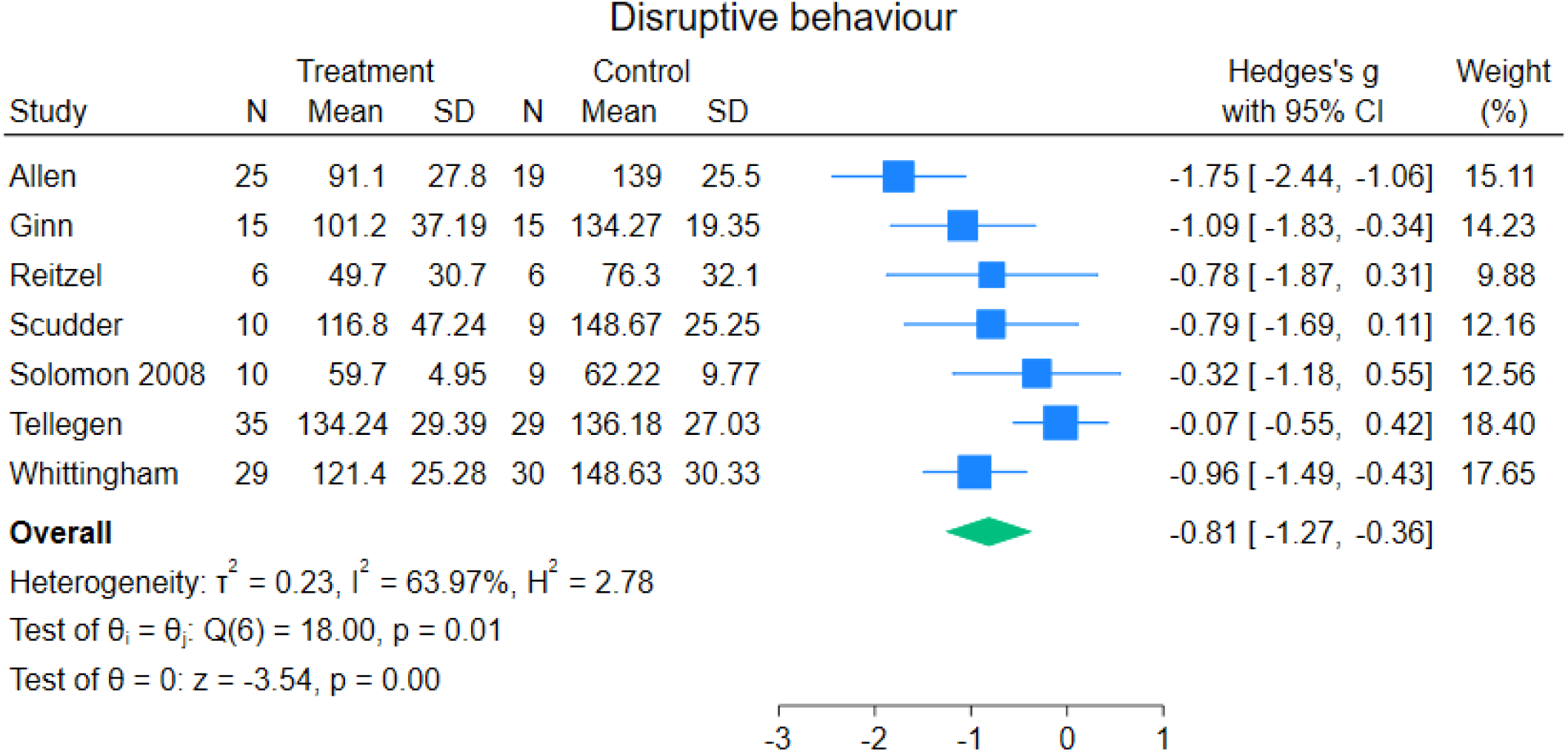

Test of interaction comparing the different control groups, i.e., usual care and waiting list, showed no evidence of difference (*p* = 0.10). Test of interaction comparing the effects between the behavioural interventions and the NDBIs showed no evidence of a difference (*p* = 0.49). Test of interaction comparing the different age-groups showed no evidence of a difference (*p* = 0.76). None of the remaining predefined subgroup analyses could be performed due to lack of relevant data.

### Joint Attention

Three of the trials assessed the effects of PACT versus usual care (3–5). Three of the trials assessed effects of NDBIs: Caregiver Mediated Joint Engagement Intervention for Toddlers with Autism versus waiting list (6), Focus Parent Training versus usual care (7), and parent delivery of Early Start Denver Model versus usual care (8).

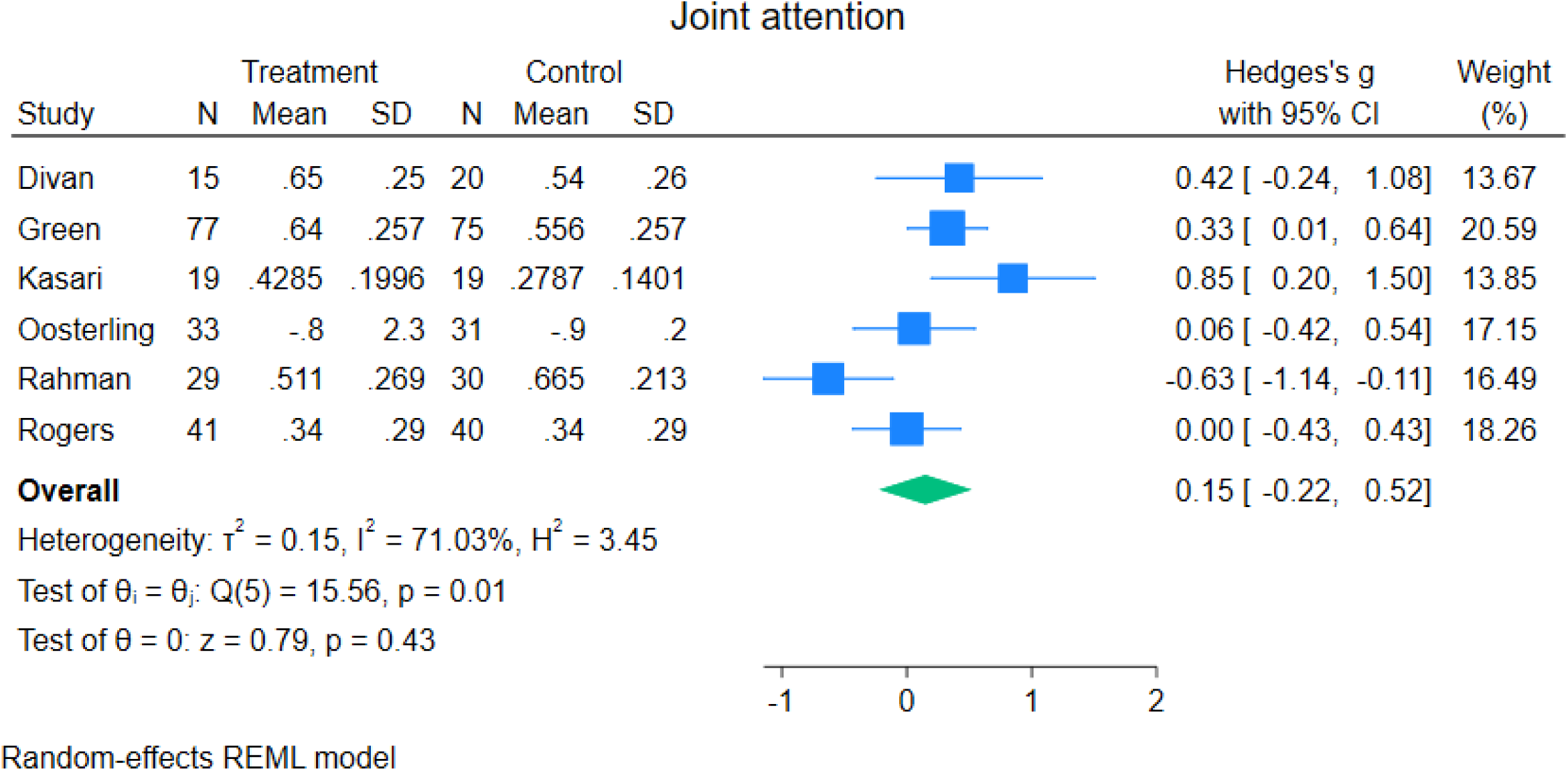

Test of interaction comparing the different control groups, i.e., usual care and waiting list, showed evidence of difference (*p* =0.03). When the trials using usual care as the control intervention were analysed separately, meta-analysis showed no evidence of a difference (SMD 0.04; 95% CI −0.30 to 0.38; 5 trials) (3–5, 7, 8). Only one trial used waiting list as the control (6). Test of interaction comparing the effects between the developmental interventions and NDBIs showed no evidence of a difference (*p* = 0.60). Test of interaction comparing the different age groups showed no evidence of a difference (*p* = 0.43). None of the remaining predefined subgroup analyses could be performed due to lack of relevant data.

### Parent sensitivity/synchronicity

Four of the trials assessed the effects of PACT versus usual care (3–5, 9). Whitehouse et al. (2021) assessed effects of iBASIS-VIPP versus usual care. The trial by Poslawsky et al. (2015) assessed the effects of Video-feedback Intervention to promote Positive Parenting adapted to Autism (VIPP-AUTI) versus usual care. Siller et al. (2013) assessed effects of Focused Playtime Intervention versus usual care. Gong et al. (2025) assessed effects of the Heart-Mind-Behavior Training Program versus waiting list.

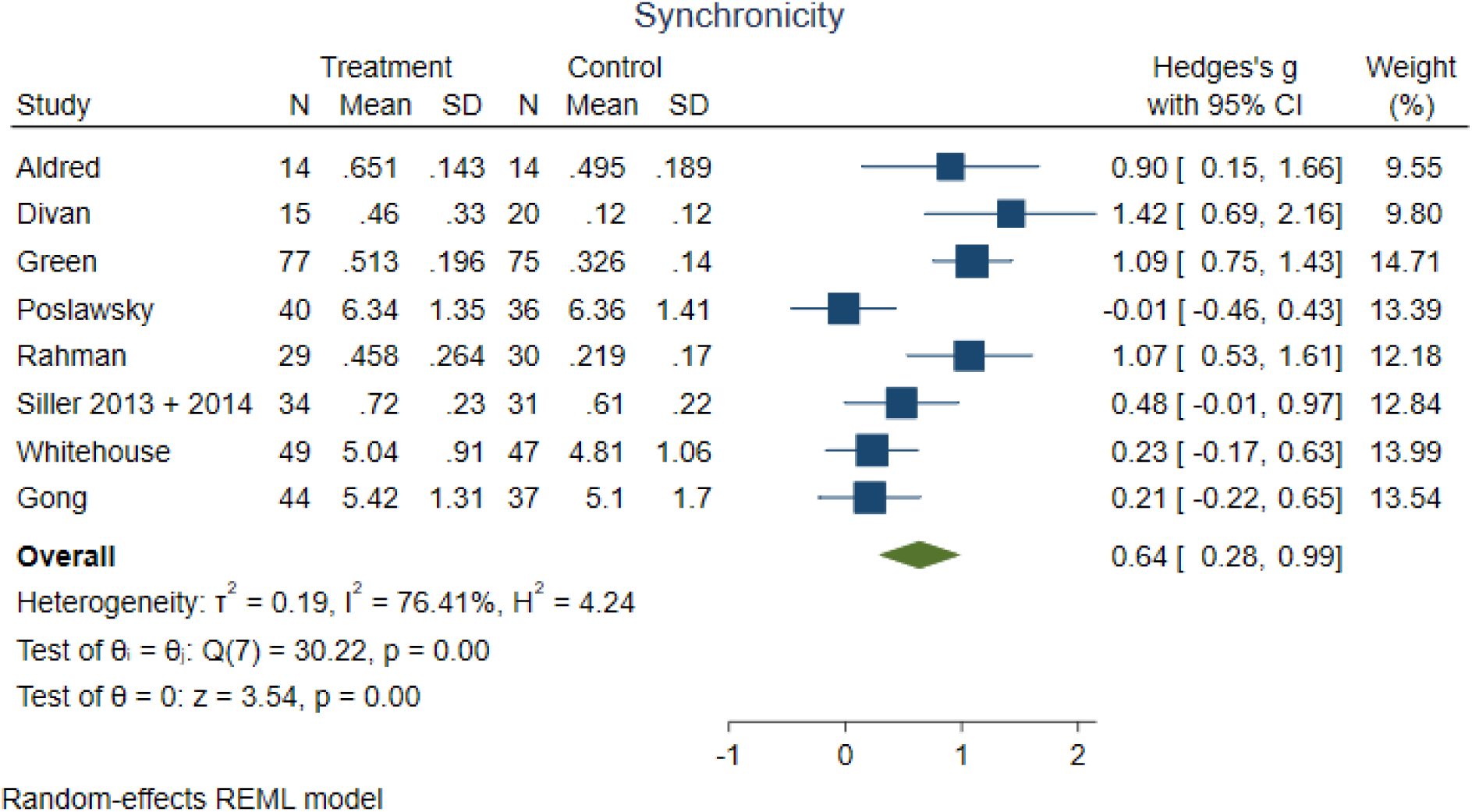

Test of interaction comparing the different control groups, i.e., usual care and waiting list, showed no evidence of difference (*p* = 0.10). Test of interaction comparing the different age groups showed no evidence of a difference (*p* = 0.06). None of the remaining predefined subgroup analyses could be performed due to lack of relevant data.

### Parent Fidelity

The trial by Brian et al. (2017) assessed the effect of the intervention Social ABC’s versus waiting list, Reitzel et al. (2013) assessed effects of Functional Behavior Skills Training versus usual care, and Rogers et al. (2012) assessed effects of parent delivery of Early Start Denver Model (P-ESDM) versus usual care.

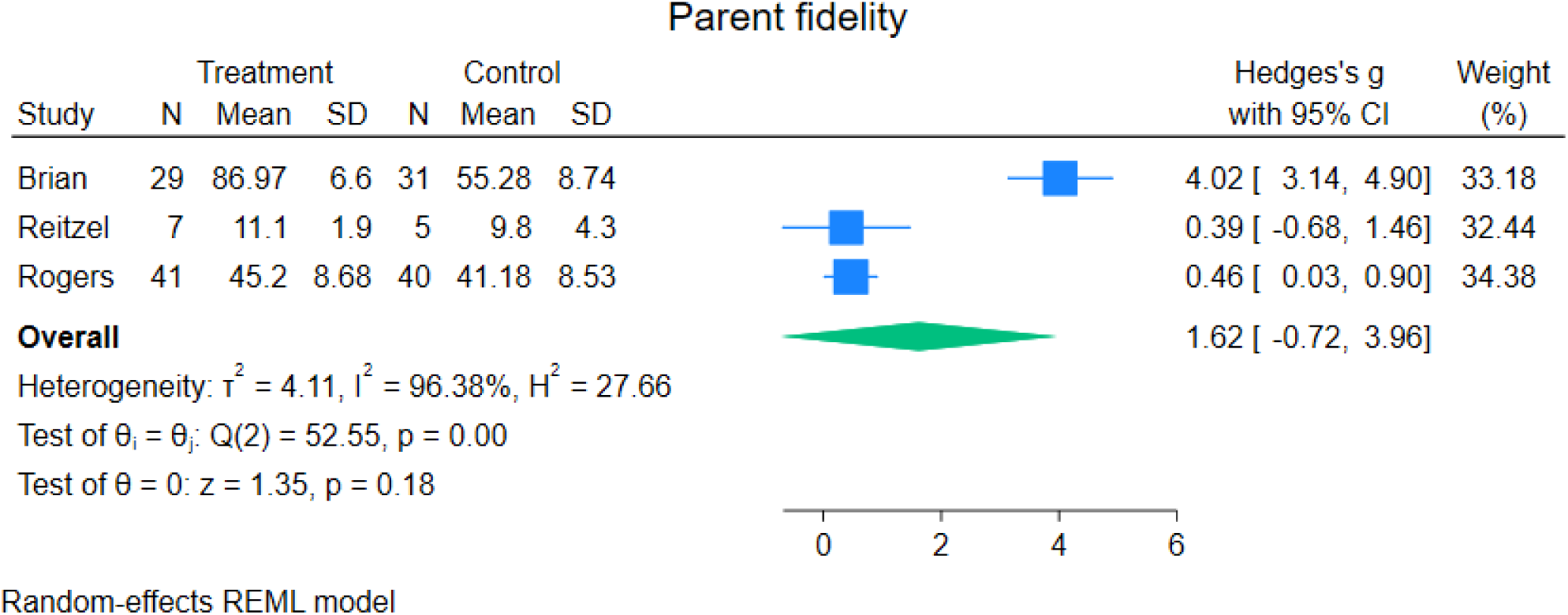

Test of interaction comparing the different control groups, i.e., usual care and waiting list, showed evidence of difference (*p* =0.00). When the trials using usual care as the control intervention were analysed separately, meta-analysis showed evidence of a beneficial effect (SMD 0.45; 95% CI 0.05 to 0.86; 2 trials) (8, 10). Test of interaction comparing the effects between the behavioural interventions and NDBIs showed no evidence of a difference (*p* = 0.32). None of the remaining predefined subgroup analyses could be performed due to lack of relevant data.

### Autism Characteristics

Three trials assessed effects of NDBIs: Brian et al. (2017) assessed the effect of Social ABC’s versus waiting list, Ginn et al. (2017) assessed effects of CDIT versus waiting list, de Korte et al. (2021) assessed effects of Pivotal Response Training versus usual care. The remaining four trials assessed effects of developmental interventions: Pajareya & Nopmaneejumruslers (2011) assessed effects of DIR-Floortime versus waiting list, Solomon & Mahoney (11, 12) assessed effects of PLAY versus usual care, Whitehouse et al. (2021) assessed effects of I-BASIS-VIPP versus usual care, and Gong et al. (2025) assessed effects of the Heart-Mind-Behavior Training Program versus waiting list.

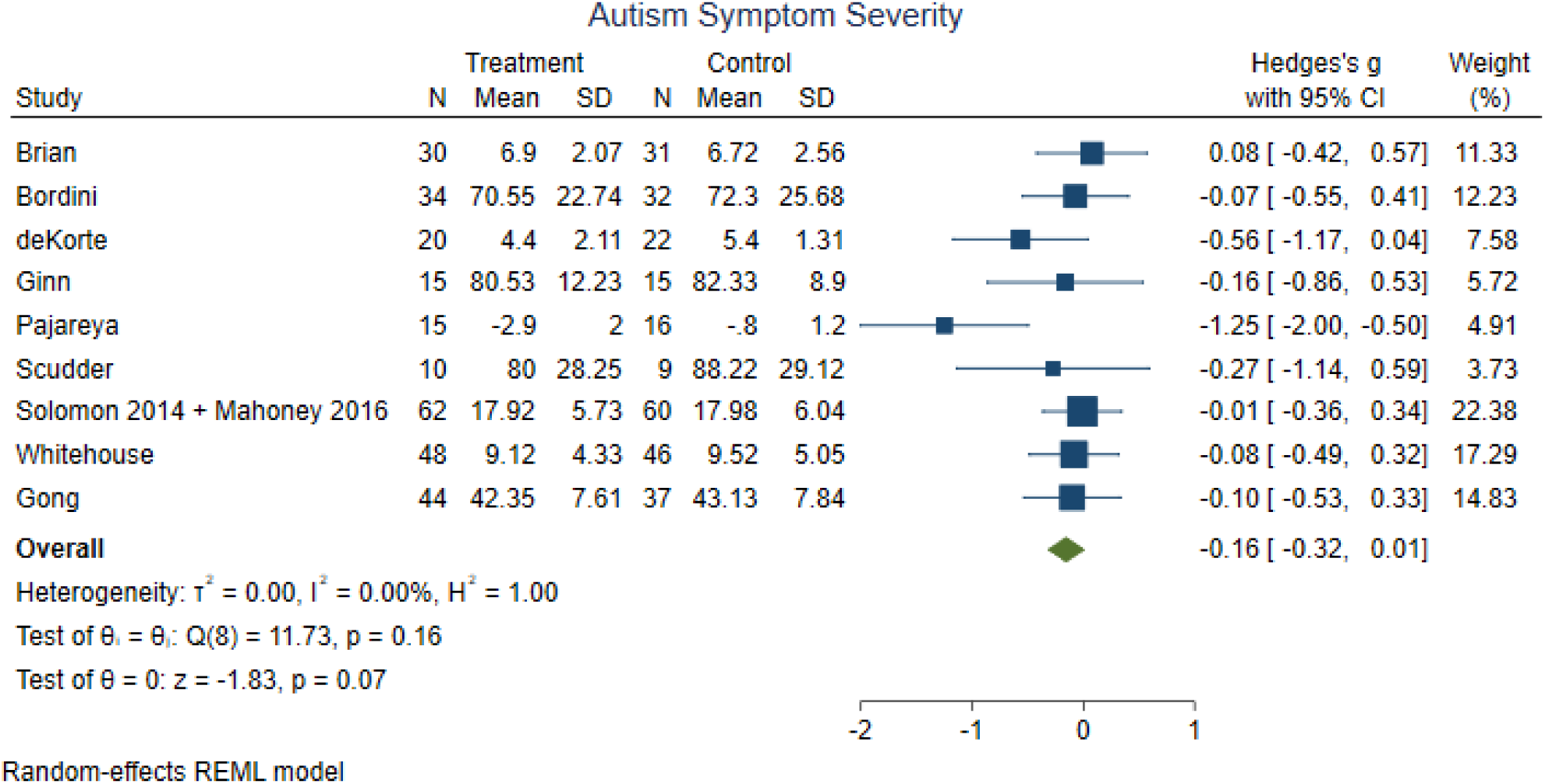

Test of interaction comparing the different control groups, i.e., usual care and waiting list, showed no evidence of a difference (*p* = 0.48). Test of interaction comparing the effects between the developmental interventions and NDBI showed no evidence of a difference (*p* = 0.88). Test of interaction comparing the different age-groups showed no evidence of a difference (*p* = 0.28). Test of interaction comparing the different groups of autism severity showed no evidence of a difference (*p* = 0.63). None of the remaining predefined subgroup analyses could be performed due to a lack of relevant data.

### Child Social Communication

Four trials assessed effects of NDBIs; Rogers et al. (2012) assessed effects of P-ESDM versus usual care, Brian et al. (2017) assessed the effects of Social ABC’s versus waiting list, both the Drew et al. (2002), and the trial by Oosterling et al. (2010) assessed effects of Focus Parent Training versus usual care. Five trials assessed the effects of developmental interventions: Carter et al. (2011) assessed effects of Hanen’s More than Words versus no treatment, and Mahoney & Solomon (2016) assessed effects of PLAY versus usual care, Gong et al. (2025) assessed effects of the Heart-Mind-Behavior Training Program versus waiting list, while Aldred et al. (2004) and Divan et al. (2019) assessed effects of PACT versus usual care.

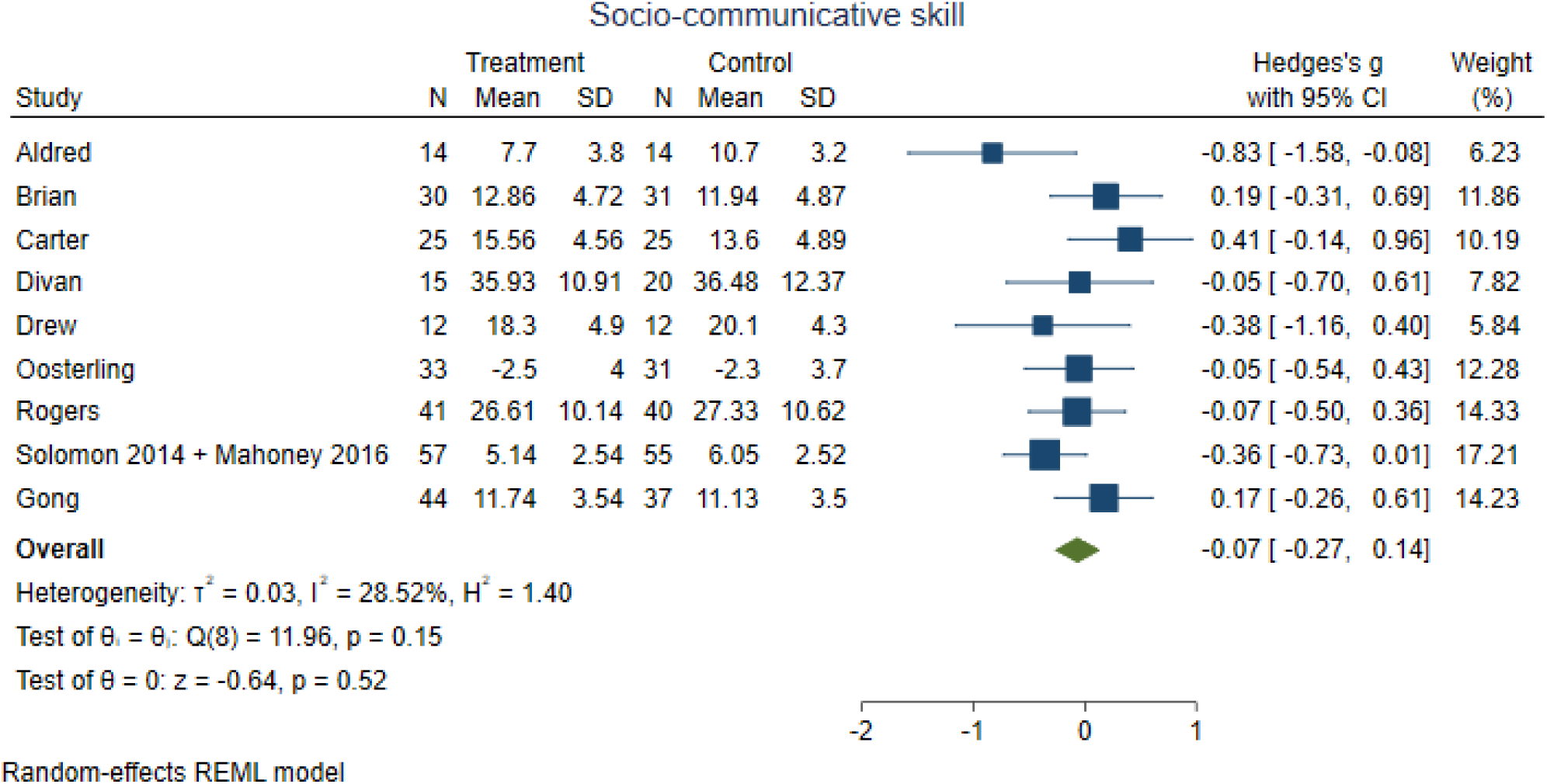

Test of interaction comparing the different control groups, i.e., usual care and waiting list, showed evidence of difference (*p* =0.02). When the trials using usual care as the control intervention were analysed separately, meta-analysis showed evidence of a beneficial effect (SMD −0.24; 95% CI −0.45 to −0.03; 6 trials) (3, 7–9, 11–13). Test of interaction comparing the effects between the developmental interventions and NDBIs showed no evidence of a difference (*p* = 0.76). Test of interaction comparing the different age groups showed no evidence of a difference (*p* = 0.82). None of the remaining predefined subgroup analyses could be performed due to lack of relevant data.

### Child Repetitive Behavior

Three studies assessed effects of NDBIs: Rogers et al. (2012) assessed effects of P-ESDM versus usual care, Brian et al. (2017) assessed the effects of Social ABC’s versus waiting list, Drew et al. (2002) assessed effects of Focus Parent Training versus usual care. Two trials assessed the effects of PACT versus usual care (5, 9). Mahoney & Solomon assessed effects of PLAY versus usual care (11, 12). Gong et al. (2025) assessed effects of the Heart-Mind-Behavior Training Program versus waiting list. Grahame et al. (2015) assessed effects of Managing Repetitive Behavior Programme versus waiting list.

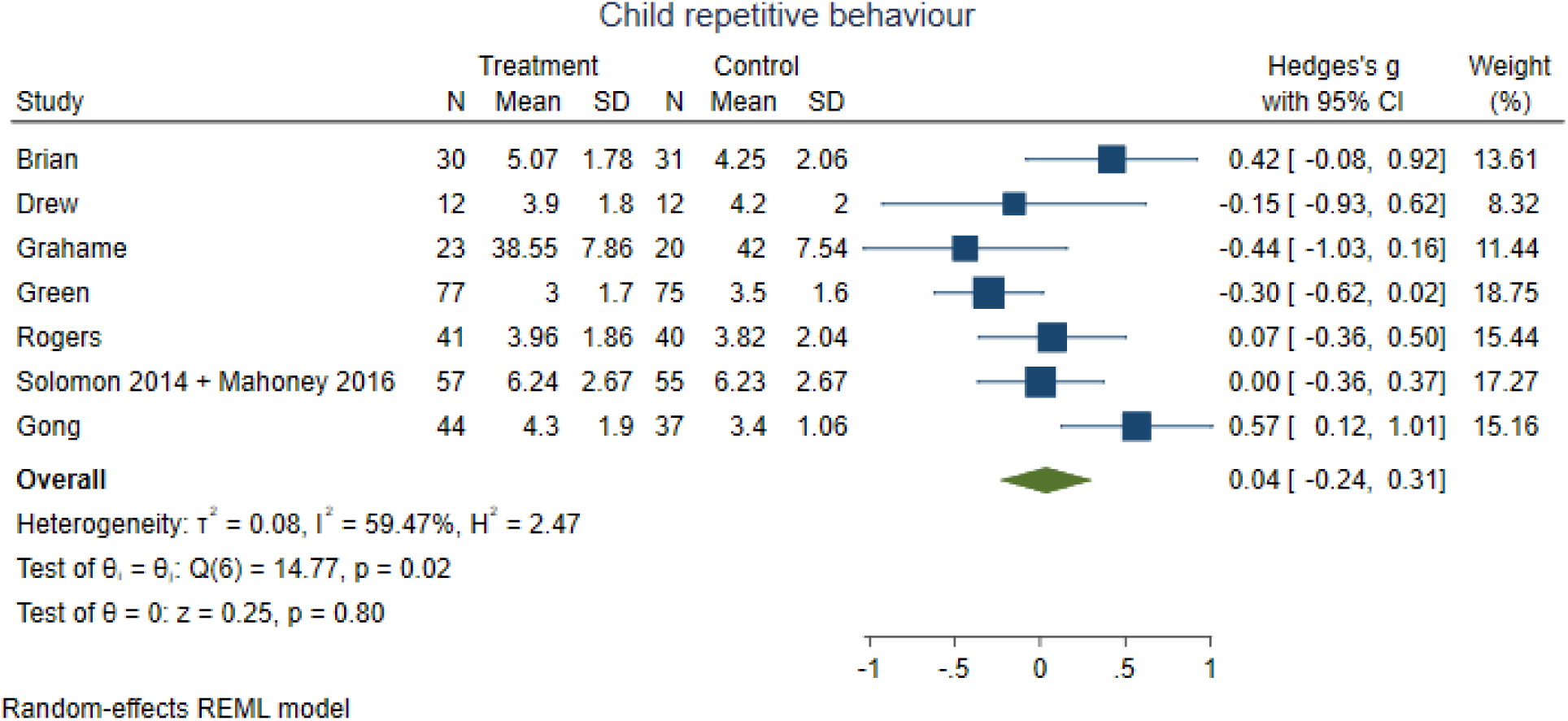

Test of interaction comparing the different control groups, i.e., usual care and waiting list, showed no evidence of a difference (*p* = 0.32). Test of interaction comparing the effects between the behavioral and developmental interventions and NDBIs showed no evidence of a difference (*p* = 0.21). Test of interaction comparing the different age groups showed no evidence of a difference (*p* = 0.72). None of the remaining predefined subgroup analyses could be performed due to a lack of relevant data.

